# Common- and rare-variant genetic architecture of heart failure across the allele frequency spectrum

**DOI:** 10.1101/2023.07.16.23292724

**Authors:** David S.M. Lee, Kathleen M. Cardone, David Y. Zhang, Noah L. Tsao, Sarah Abramowitz, Pranav Sharma, John S. DePaolo, Mitchell Conery, Krishna G. Aragam, Kiran Biddinger, Ozan Dilitikas, Lily Hoffman-Andrews, Renae L. Judy, Atlas Khan, Iftikhar Kulo, Megan J. Puckelwartz, Nosheen Reza, Benjamin A. Satterfield, Pankhuri Singhal, Regeneron Genetics Center, Zoltan P. Arany, Thomas P. Cappola, Eric Carruth, Sharlene M. Day, Ron Do, Christopher M. Haggarty, Jacob Joseph, Elizabeth M. McNally, Girish Nadkarni, Anjali T. Owens, Daniel J. Rader, Marylyn D. Ritchie, Yan V. Sun, Benjamin F. Voight, Michael G. Levin, Scott M. Damrauer

**Author notes:** These authors contributed equally to this work (Michael G. Levin, Scott M. Damrauer).

## Abstract

Heart failure (HF) is a complex trait, influenced by environmental and genetic factors, which affects over 30 million individuals worldwide. Historically, the genetics of HF have been studied in Mendelian forms of disease, where rare genetic variants have been linked to familial cardiomyopathies. More recently, genome-wide association studies (GWAS) have successfully identified common genetic variants associated with risk of HF. However, the relative importance of genetic variants across the allele-frequency spectrum remains incompletely characterized. Here, we report the results of common- and rare-variant association studies of all-cause heart failure, applying recently developed methods to quantify the heritability of HF attributable to different classes of genetic variation. We combine GWAS data across multiple populations including 207,346 individuals with HF and 2,151,210 without, identifying 176 risk loci at genome-wide significance (P-value < 5×10^−8^). Signals at newly identified common-variant loci include coding variants in Mendelian cardiomyopathy genes (*MYBPC3*, *BAG3*) and in regulators of lipoprotein (*LPL*) and glucose metabolism (*GIPR*, *GLP1R*). These signals are enriched in myocyte and adipocyte cell types and can be clustered into 5 broad modules based on pleiotropic associations with anthropomorphic traits/obesity, blood pressure/renal function, atherosclerosis/lipids, immune activity, and arrhythmias. Gene burden studies across three biobanks (PMBB, UKB, AOU), including 27,208 individuals with HF and 349,126 without, uncover exome-wide significant (P-value < 1.57×10^−6^) associations for HF and rare predicted loss-of-function (pLoF) variants in *TTN*, *MYBPC3*, *FLNC, and BAG3.* Total burden heritability of rare coding variants (2.2%, 95% CI 0.99-3.5%) is highly concentrated in a small set of Mendelian cardiomyopathy genes, while common variant heritability (4.3%, 95% CI 3.9-4.7%) is more diffusely spread throughout the genome. Finally, we show that common-variant background, in the form of a polygenic risk score (PRS), significantly modifies the risk of HF among carriers of pathogenic truncating variants in the Mendelian cardiomyopathy gene TTN. Together, these findings provide a genetic link between dysregulated metabolism and HF, and suggest a significant polygenic component to HF exists that is not captured by current clinical genetic testing.

## Introduction

Heart failure (HF) is a heterogeneous clinical syndrome affecting >30 million individuals worldwide and is the leading cause of unplanned hospital admissions in the United States for individuals >65 years old. HF arises from diverse etiologic causes, including chronic ischemic heart disease, myocardial infarction, diabetes, hypertension, obesity, arrhythmias, infiltrative and inflammatory disorders, and exposure to drugs or environmental toxins^1^. Although there has been progress in mapping genes involved in familial/Mendelian forms of cardiomyopathy, the genetic architecture of common HF remains less well understood. Unlike for other common cardiometabolic traits, the number of genetic loci associated with HF has remained comparatively limited despite increasing sample sizes^2–6^. In a subset of affected individuals, pathogenic variants affecting a core set of cardiomyopathy genes can be identified as the putative cause of HF using clinical panel-based genetic testing^7,8^. As with other complex disease traits, common less-penetrant variants influencing these core genes might also contribute to HF risk^4,6,9^; yet the catalog of such variants and their cumulative contribution to HF has not been fully elaborated. More broadly, few studies have integrated effects across common and rare variants to quantify the extent they implicate similar genes and pathways in HF.

Here, we conduct a comprehensive analysis of genetic variants across the allele frequency spectrum to examine shared and distinct pathways implicated by common and rare genetic variants in HF. Specifically, we perform a multi-population GWAS meta-analysis to identify loci where common genetic variants are associated with all-cause HF, uncover 5 genetic modules of HF through clustering of HF loci, and in parallel conduct rare variant gene burden studies across 3 combined cohorts to identify genes where rare predicted damaging coding variants contribute to HF risk. We leverage this data to estimate the heritability of HF and quantify enrichment of both common and rare variants in a core set of known cardiomyopathy genes. Finally, we construct a multi-population PRS to evaluate how common variants act collectively to modify the penetrance of loss-of-function truncating variants in *TTN*, the most common Mendelian risk factor for cardiomyopathy. Overall, our findings highlight the significant polygenic component of HF risk within the general population that is currently not captured by clinical genetic testing, and suggest that polygenic background may be important even among carriers of highly penetrant loss-of-function variants in known monogenic cardiomyopathy genes.

## Results

### GWAS of All-Cause Heart Failure

We conducted a multi-population GWAS meta-analysis of all-cause HF using an inverse-variance weighted fixed effects model, assembled from participants of large consortia and medical biobanks (Figure 1). Comprised of 207,346 individuals with and 2,151,210 individuals without HF, this analysis represents an approximate doubling in the effective sample size of the previously largest published HF GWAS (Figure 1)^3^. We combined individuals across four population groups with genetic similarity to 1000 Genomes Project reference superpopulations^12^: a European group (EUR, 81.1% of effective sample size), an East Asian group (EAS, 6.5%), an African group including admixed African Americans (AFR, 9.7%), and an Admixed American group (AMR, 2.6%). The analysis identified 9,990 variants at 176 loci where genetic associations reached genome-wide significance (P-value < 5×10^−8^ - Figure 2, Suppl. Fig. 1 and Suppl. Table 1), representing an approximately 3.7-fold increase in the number of genome-wide significant loci compared to prior analyses^3^. Given the diverse nature of the contributing cohorts we additionally performed meta-regression using MR-MEGA, which partitions heterogeneity into ancestry-correlated and residual effects. Out of 176 genome-wide significant loci from our primary analysis, 97 remained genome-wide significant in this sensitivity analysis, and all loci identified by METAL were replicated at the Bonferroni-corrected threshold with P-value < 2.8×10^−4^ (Suppl. Fig. 2 and Suppl. Table 2) using MR-MEGA. MR-MEGA identified an additional 10 loci at the genome-wide significance threshold that were not identified by METAL. Genes implicated by these loci by proximity include XKR6 and PTPRG which have both been associated with coronary artery disease and stroke^13,14^, and ZPR1 a gene strongly associated with circulating lipid levels^15^. Because MR-MEGA does not provide effect sizes other than for each ancestry component, we focused further downstream analysis using results from METAL.

**Figure 1:**
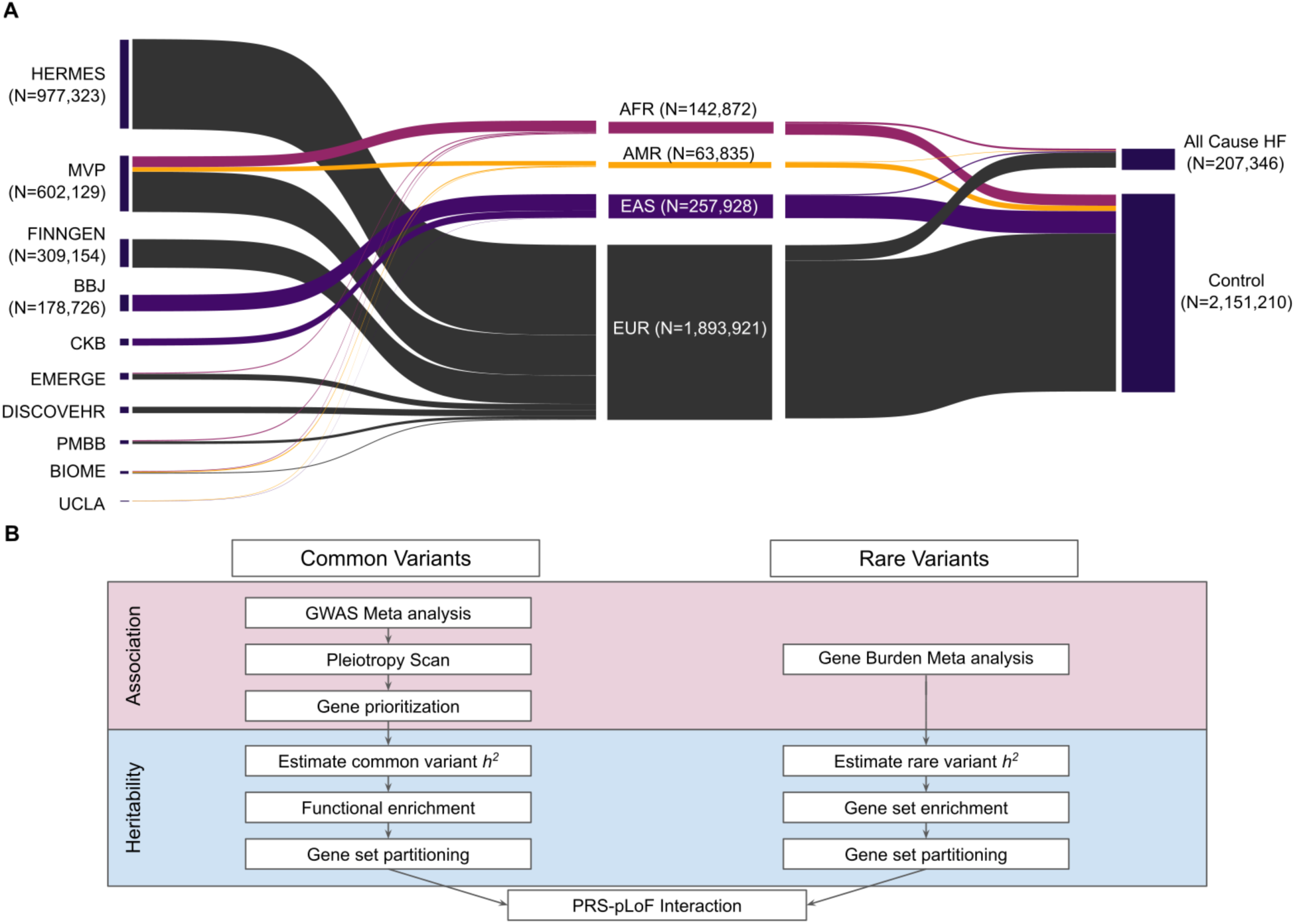
(A) Study population used in GWAS meta-analysis. We conducted a meta-analysis of heart failure across 10 cohorts comprising a combined count of N=207,346 participants with HF and 2,151,210 participants without HF. Summary statistics were collected across 10 cohorts and meta-analyzed using METAL. (B) Overview of study design, which encompasses GWAS meta-analysis and estimate of common variant h^2^ in parallel with an analysis of rare coding variant associations with heart failure and rare variant h^2^ across 3 cohorts.

**Figure 2:**
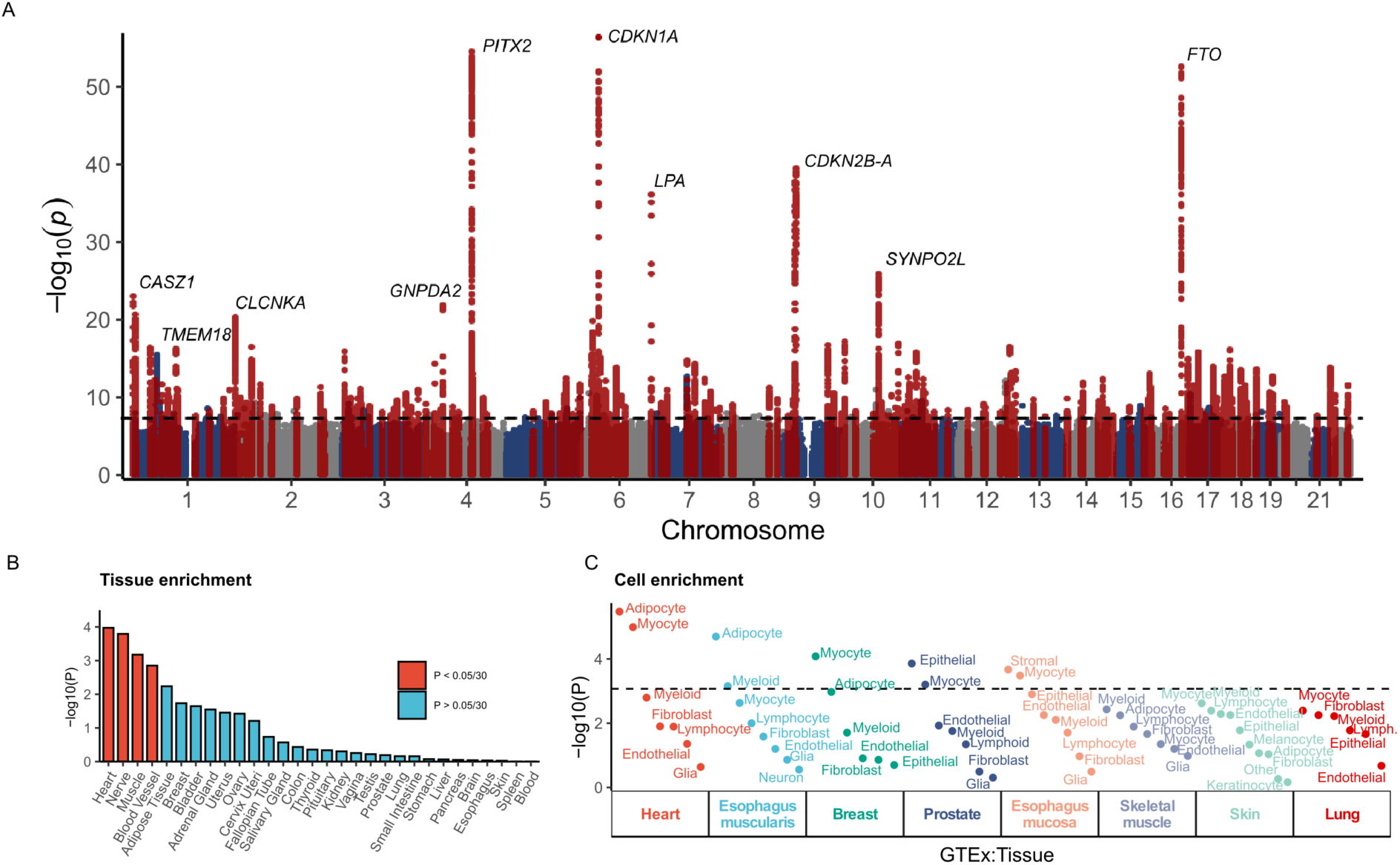
(A) Manhattan plot for common variant association in multi population meta-analysis (207,346 HF, 2,151,210 non-HF). Each point represents a variant, organized by chromosomal position along the x-axis. Lead variants are annotated with the closest mapped gene and the surrounding +/-500kb is highlighted in red. (B) Enrichment for tissue-specific differentially increased gene expression of GWAS prioritized genes. P-values determined by hypergeometric test. Enrichment P-values above the Bonferroni-corrected significance threshold (0.05/30) are shaded red. (C) Enrichment for expression of GWAS prioritized genes in GTEx multi-tissue single cell sequencing data. P-values determined by hypergeometric test (see Methods). Dashed black line represents the Bonferroni-corrected significance threshold (0.05/59).

Of the 47 lead variants reported in the previously largest published HF GWAS^3^, 46 variants (or perfect proxies) were associated with HF with concordant direction of effects (two-tailed binomial P = 6.82 × 10^−13^) in the current analysis. Forty one of 46 (89%) were significant at the genome-wide threshold, while 5 of 46 (11%) (rs1712355, rs12325072, rs138178446, rs150966198, rs6944634) remained significant at a conservative Bonferroni-corrected replication threshold (P-value < 0.05/46) with concordant directions of effect. Among 61 additional lead variants previously reported across 3 studies^2,16,17^ 54 (89%; two-tailed binomial P = 4.32 × 10^−10^) were replicated at the genome-wide threshold with concordant directions of effect, while the remaining 7 variants remained significant at a conservative Bonferroni-corrected replication threshold (P-value < 0.05/7) with concordant directions of effect (Suppl. Table 3). Of the 176 loci identified in our study (Fig. 2A), 105 were located >500 kb from lead variants identified in previous HF GWAS (where multiple signals were present at some loci)^3^. Two association signals were genome-wide significant in population-restricted analyses but not significant in the combined meta-analysis – these included the locus tagged by rs373205748 which was significant in the EAS population group, maps to the NEURL1 gene, and has previously been associated with arrhythmias in the Japanese population^18^, and an intronic T insertion in the CELF gene (chr11: 4748724) which was significant in the AFR population group.

#### Pleiotropic genetic associations

Many lead variants in our study show pleiotropic genetic associations with known HF risk factors, including BMI, atrial fibrillation, LVEF, systolic and diastolic blood pressure, coronary artery disease, lipid metabolism, diabetes, alcohol consumption, smoking, and kidney function among previously-published GWAS available in the NHGRI/EBI GWAS Catalog (Suppl. Fig. 5, Suppl. Table 3)^19–23^. To understand the full spectrum of pleiotropic associations of HF risk loci, we performed phenome-wide colocalization using summary results from genetic association studies of >2000 traits assessed in the UK Biobank, including phecodes, biomarkers, and physiologic assessments^24^. At the 176 HF risk loci, we identified 536 traits with a genome-wide significant association within +/-500kb of the lead HF signal (Suppl. Table 5). To determine whether these pleiotropic associations were driven by the same genetic signal as HF, we performed multi-trait colocalization at each locus using HyPrColoc^25^, finding putative evidence of shared associations (posterior probability >= 0.25) at 125/176 (71%) loci. Loci sharing the most pleiotropic associations with HF included *FTO* (124 traits), *SH2B3* (106 traits), *ANAPC4* (77 traits), *ABO* (77 traits), and *MC4R* (71 traits). These loci included shared associations with anthropometric/blood pressure traits (*FTO*, *ANAPC4*, *MC4R*) and metabolic/blood cell traits (*SH2B3*, *ABO*).

We next focused on loci with high-confidence evidence of colocalization (posterior probability > 0.7), defining a bipartite network linking HF risk loci with their pleiotropic trait/disease associations. Using a projection of this network, we used Louvain community detection to identify modules of traits/diseases that are connected based on evidence of colocalization at multiple HF loci. This analysis revealed 5 disease modules: an anthropometric/obesity module, including measures of body size (waist/hip circumference, height) and composition (body fat, body water, impedance); a blood pressure/renal module; an atherosclerosis/lipids module containing lipoprotein(a) and other major circulating lipids/lipoproteins, as well as manifestations of atherosclerosis including coronary/peripheral/valvular heart disease; an immunologic module containing type 1 diabetes, hypothyroidism, rheumatoid arthritis and blood cell traits; and an arrhythmia module including atrial fibrillation (Suppl. Table 6).

#### Fine-mapping of risk loci

We performed fine-mapping of the 176 loci reaching genome-wide significance in our multi-population meta-analysis to identify candidate causal variants. Compared to fine-mapping of the EUR population alone, the multi-population analysis improved fine-mapping resolution, decreasing the median size of the 99% credible sets by 35.1%, from 55.5 to 36 fine-mapped variants per locus (Suppl. Fig. 6). At 45 loci (25%), the fine-mapped credible set contained 10 or fewer variants. We explored protein coding consequences for fine-mapped variants within 99% credible sets across all 176 lead loci (n=14,628) using Ensembl VEP (Suppl. Table 7). We additionally used CARMA to identify 30 secondary signals of association at lead loci (Suppl. Table 8). Approximately one-third of lead loci identified by METAL (61/176) contained a fine-mapped variant affecting a protein-coding sequence, with 42 loci harboring variants predicted to be either missense or protein truncating variants. Among these variants, four were predicted to be high-confidence protein truncations and were located in newly implicated HF risk loci: *SLC23A1* (rs11242462), *LPL* (rs328), *OR4P4* (rs76160133), and *MAPT* (rs754512). *SLC23A1* encodes a sodium-dependent L-ascorbic acid transporter associated with circulating plasma vitamin C levels in humans^26^ and variants at this locus have recently been found to associate with right ventricular ejection fraction^27,28^; *LPL* encodes the lipoprotein lipase enzyme which plays a central role in the regulation of lipid metabolism and transport, and has been linked previously to several cardiometabolic phenotypes, including coronary artery disease^29^, Type 2 Diabetes^30^, and BMI^31^; *OR4P4* encodes an olfactory receptor gene at the 11q11 locus which has previously been linked to obesity^32^; and *MAPT* encodes the *Tau* proteins involved in maintaining microtubule stability which lies in the MAP3K14 locus that is associated with maximum left ventricular wall thickness^33^. Other variants uncovered by our approach include a missense variant in the cardiomyopathy-associated gene *MYBPC3* (rs11570076, tagged by the novel HF locus at rs2904129) located within a key domain for regulating the cMyBP-C interaction with S2 domain of myosin, and the previously described *BAG3* missense variant rs2234962 (tagged by rs17617337) that is enriched among patients with dilated cardiomyopathy^34^.

#### Gene-prioritization

To nominate genes implicated by GWAS loci, we used an ensemble approach combining physical mapping to the nearest-gene, the coding consequence of fine-mapped variants, and results from TWAS and MAGMA (see Methods) to prioritize 577 genes, of which 73 genes were prioritized by at least 3 methods (Suppl. Table 9). Among this subset of highly prioritized genes at novel loci were the glucose-dependent insulinotropic polypeptide receptor (*GIPR*) and the glucagon-like peptide-1 receptor (*GLP1R*). Of note, neither of these genes have previously been associated with HF in humans, although cardioselective deletion of *GIPR* has previously been shown to improve survival and reduce adverse remodeling following myocardial ischemic injury in mice^35^. Given the role of these genes in glucose homeostasis and obesity, we performed multi-trait colocalization using GWAS summary statistics to evaluate the genetic evidence supporting shared or independent signals for HF, diabetes^5^, hemoglobin A1c^36^, and body mass index^37^. At the *GIPR* locus there was 99.4% posterior probability of a shared genetic variant influencing both HF and BMI. In contrast, at the *GLP1R* locus we did not find evidence of colocalization, indicating the possibility that the HF, diabetes, and obesity signals at this locus may be due to separate mechanisms.

#### Tissue, cell, and pathway-enrichment

Using the full set of genes nominated at GWAS loci (n = 577), we queried public data to identify relevant tissues and biological pathways implicated by loci from our GWAS meta-analysis. As expected, heart and blood vessels were among the tissues containing significantly up-regulated expression of GWAS-prioritized genes (Fig. 2B) while hallmark gene sets for myogenesis and hypoxia were also enriched (Suppl. Fig. 7). GWAS-prioritized genes were also 1.7-fold enriched in known drug targets recorded in DrugBank (95% CI 1.2-2.4, P-value 0.0035, Fisher’s Exact Test). Among phenotype-associated gene sets published in the NHGRI/EBI GWAS Catalog, GWAS-prioritized genes overlapped most significantly with gene sets associated with coronary artery disease, systolic blood pressure, body mass index, and atrial fibrillation, reflecting the multifactorial nature of HF (Suppl. Fig. 8). Consistent with an emerging role for metabolic processes in HF, when matched against single cell expression data from 13 cell types across 8 different tissues published by GTEx^38^, GWAS-prioritized genes were most significantly enriched in myocyte and adipocyte cells in the heart (Fig. 2C). Cardiac adipose tissue is thought to contribute to cardiac pathology through myriad mechanisms^39^, and this enrichment in cardiac adipocytes suggests part of their contribution to HF is mediated by gene expression changes resulting from germline genetic variation. Notably, prior GWAS had only reported enrichment of GWAS genes in cardiac myocytes^3^.

### Rare Variant Genetic Architecture of All-cause Heart Failure

To assess the rare variant architecture of HF, we conducted an exome-collapsing gene-based association test using linked electronic health record data and whole exome or whole genome sequencing for 376,334 participants from the Penn Medicine Biobank (PMBB, n = 39,544), UK Biobank (UKB, n = 150,442), and All of Us (AOU, n = 186,348) research program. We meta-analyzed results from gene-based group tests for rare predicted loss-of-function (pLoF) variants using SAIGE-GENE^40^ (Figure 3) for 20,675 participants with and 287,123 participants without HF from the EUR population and 6,533 participants with and 62,003 participants without HF from the AFR population. Because missense variants may produce distinct consequences compared to pLoF variants^41^, we repeated the gene-based analysis, testing associations with carrier status for a predicted damaging missense variant and all-cause HF.

**Figure 3:**
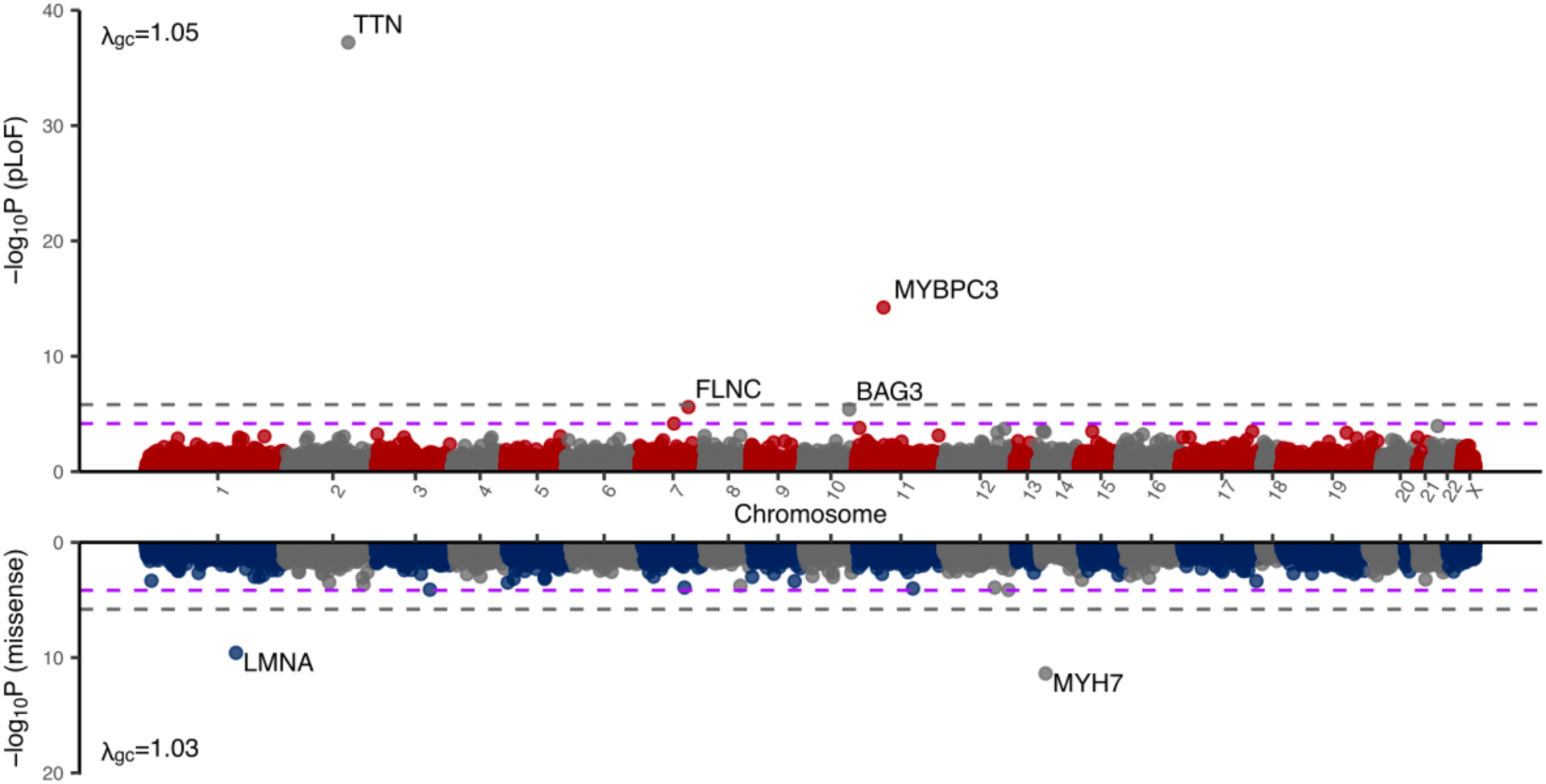
Gene burden results for multi-population meta-analysis across PMBB, AOU, and UKB. pLoF variants and damaging missense variants were aggregated separately and the burden of pLoF and damaging missense variants was tested for an association with heart failure. Dashed grey line represents the Bonferroni-corrected significance threshold while purple line represents the FDR < 0.05 threshold. *P-*values derived from fixed-effects meta-analysis of burden testing statistics implemented in SAIGE-GENE(+).

Under this framework, we replicated the previously-reported^7^ association between rare pLoF variants in *TTN* and HF (P-value 6.12×10^−38^, Suppl. Table 10) at a Bonferroni-adjusted exome-wide significance (P-value < 1.57×10^−6^ [0.05/31856]). We also uncovered Bonferroni adjusted exome-wide significant associations for rare variants in *MYBPC3* (P-value 5.93×10^−15^), *MYH7* (P-value 4.26×10^−12^) and *LMNA* (P-value 2.62×10^−10^). Additionally, rare pLoF burden in two genes, *FLNC* (P-value 2.56×10^−6^), and BAG3 (P-value 4.02×10^−6^) reached exome-wide significance at FDR < 0.05. Finally, pLoF variants in *TET2* had been associated with HF in an exome-collapsing study of HF containing participants from two large clinical trials^7^, however we found no association between *TET2* and all-cause HF in our meta-analysis (P-value 0.49).

#### Rare-variant associations in Mendelian cardiomyopathy genes

Taking a targeted approach, we next assessed contributions of pLoF variants in a set of 71 expert-curated (NIH ClinGen) genes associated with Mendelian forms of dilated, hypertrophic, or arrhythmogenic cardiomyopathy (representing the most prevalent forms of inherited cardiomyopathy^42^) to all-cause HF risk. We grouped these genes by levels of evidence supporting their role as a monogenic cause of cardiomyopathy as determined by expert panels^43–45^. This resulted in 24 definitive evidence genes, 6 strong or moderate evidence genes, 33 limited or disputed evidence genes, and 8 genes without evidence for involvement in human cardiomyopathy despite experimental evidence in the literature (Suppl. Table 13).

We next asked whether pLoF burden in each cardiomyopathy gene was associated with increased odds of all-cause HF at an exploratory significance threshold of P-value < 0.05. Among the 24 genes classified as definitive cardiomyopathy genes, pLoF carrier status in 9 conferred increased odds of all-cause HF (BAG3, MYBPC3, LMNA, FLNC, RBM20, TTN, TNNT2, DSP, PKP2 in our combined cohort - Suppl. Fig. 9, Suppl. Table 10). No genes with moderate to strong evidence supporting their association with cardiomyopathy demonstrated associations with all-cause HF, while 2 of 33 genes with limited or disputed evidence were associated with increased odds of all-cause HF (*PSEN1, CALR3*) in this exploratory analysis. Finally, no genes deemed to have no evidence of an association with any cardiomyopathy showed even nominal associations with a diagnosis of all-cause HF. Compared to carrier status for pLoF variants, predicted damaging missense variants did not increase risk of HF in most genes. *MYH7* was the clear exception to this pattern, where increased risk of HF was limited to damaging missense carriers; pLoF carrier status conferred no additional risk of HF. This supports previous observations that missense variants in *MYH7* tend to cause HCM through gain-of-function mechanisms, and highlights the comparatively limited evidence supporting a direct relationship between most loss-of-function truncating variants in *MYH7* and cardiomyopathy^46–48^.

### Heritability of Heart Failure based on Common and Rare Variants

To investigate the contribution of common genetic variants to the heritability of all-cause HF, we used LD-score regression with corresponding population-specific LD scores from Pan-UKB^49^. Common variant heritability (AF 5-50%) for all-cause HF ranged from 3-10% across the 4 populations included in our analysis (AFR 8.6%, 95% CI 6.1-11%, AMR 10%, 95% CI 6.0-14%, EAS 3.4%, 95% CI 1.9-4.9%, EUR 4.3%, 95% CI 3.9-4.7%), using population specific prevalence data for heart failure from NHANES based on self-reported race/ethnicity^50^ (Suppl. Fig. 10). Overall the range of these estimates was consistent with previous GWAS meta analyses of all-cause HF, which have estimated common variant heritability around 8.8%^2^.

We assessed the contribution of rare coding genetic variation to the heritability of all-cause HF using whole exome or whole genome sequencing from the PMBB, UKB, and AOU with Burden Heritability Regression (BHR)^11^. We estimate the total burden heritability mediated by rare (AF < 0.001) predicted damaging missense and pLoF variants is 2.24% (95% CI 0.99-3.48%, Suppl. Table 14). As expected, burden heritability was concentrated in pLoF variants, where biological impacts are the most severe, and in the rarest variants (Figure 4A), consistent with the impact of negative selection. In contrast, heritability for missense and synonymous variants was near zero.

**Figure 4:**
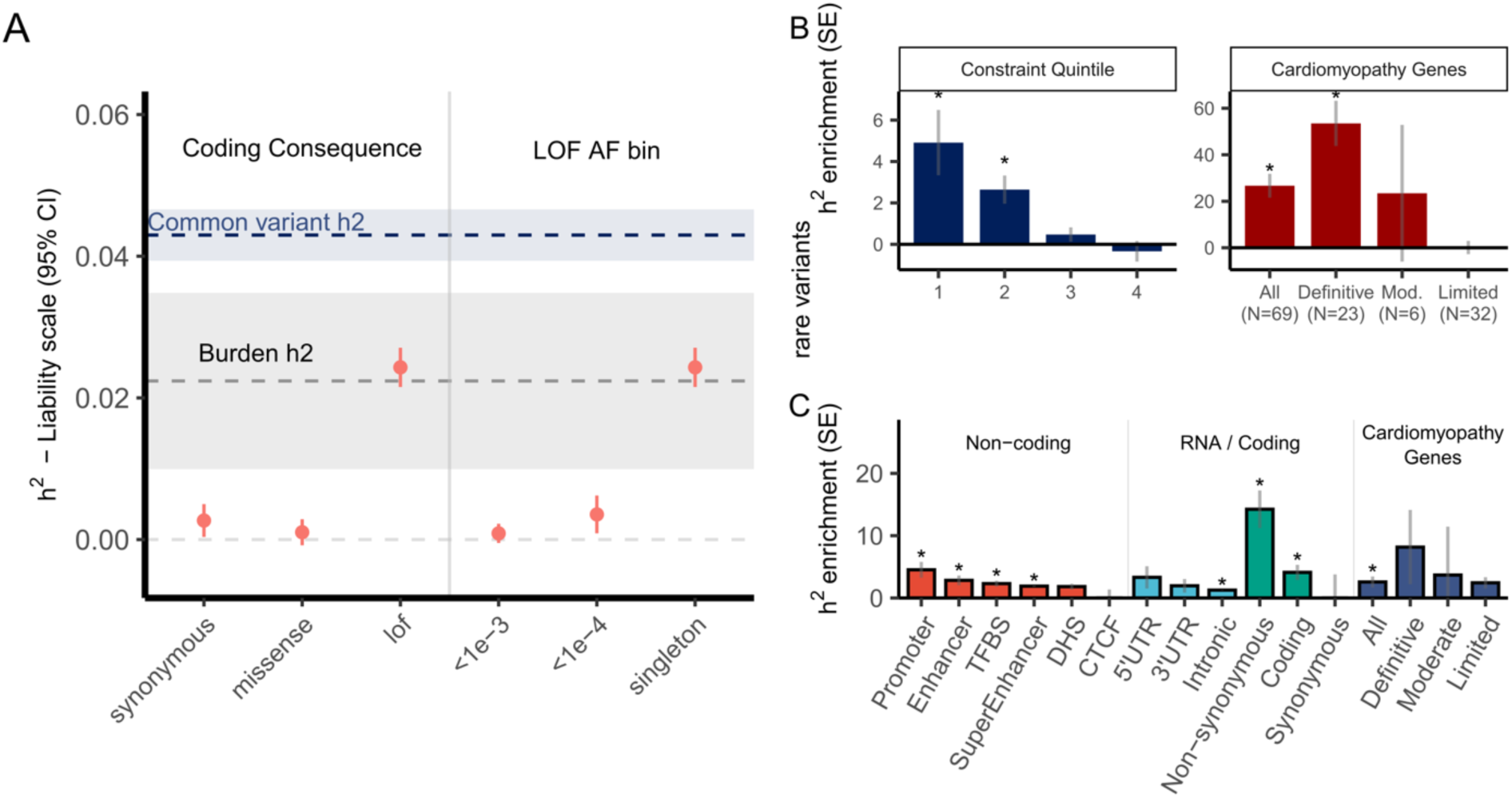
Estimates of rare coding variant burden heritability and comparison of heritability enrichment across several categories for common and rare variants. (A) BHR heritability estimate for rare coding variants split by (i) coding consequence and (ii) AF bins in the EUR population. For coding consequence, burden heritability estimates were determined using singletons. The common variant heritability estimate from the EUR GWAS population is represented by the blue dashed line with 95% confidence intervals represented by the blue shaded area. The total burden heritability of pLoF and damaging missense variants is represented by the grey dashed line, with 95% confidence intervals represented by the grey shaded area. Burden heritability is significantly concentrated in predicted loss-of-function (LOF) variants and increases with decreasing allele frequency bin. (B) Enrichment of rare variant heritability across (i) constraint quintiles or (ii) expert-reviewed genes involved in cardiomyopathies. (C) Enrichment in common variant heritability across genomic annotations and in genes implicated in cardiomyopathies as determined by stratified LDSC for Non-coding and RNA/Coding annotations and AMM for cardiomyopathy genes.

Additionally, as seen in other common complex disease traits^11^, burden heritability in all-cause HF was enriched in more constrained genes (as defined by the gnomAD LOEUF quintile – Figure 4B). These findings support a polygenic model of HF whereby common variants affecting many genes explain a comparable proportion of heritability to rare, highly penetrant coding variants across the genome.

As highlighted by results from our exome-wide association study, the burden of rare variant class (pLoF versus missense mutations) on HF risk can differ significantly for the same gene. However, whether this differential impact extends across the genome remains unclear. To systematically investigate the impact of variant class (pLoF versus damaging missense) on HF risk for each gene, we calculated the burden heritability correlation between all pLoF and predicted damaging missense singleton variants. We found that there was no significant correlation in burden heritability between pLoF and predicted damaging missense variants (r_g_ = 0.001, P-value = 0.95), suggesting that indeed the association between burden of rare variants and HF risk differs significantly between pLoF and predicted damaging missense variants on a genome-wide scale.

Next, we asked what proportion of heritability could be attributed to the set of 71 genes implicated in monogenic cardiomyopathies. While rare pLoF burden heritability was 27-fold enriched across all cardiomyopathy genes, this gene set was enriched only 2.5-fold for common variants (Figure 4B, Suppl. Table 15). This difference was most striking for definitive monogenic cardiomyopathy genes, where rare burden heritability was 60-fold enriched compared to 8.2-fold enriched for common variants. The smaller enrichment of heritability across known cardiomyopathy genes for common variants, coupled with the comparable common- and burden heritability estimates for HF, is consistent with common variant HF heritability being significantly more polygenic than rare coding variant HF heritability. Indeed, the fraction of common variant heritability mediated through the 509 protein-coding genes linked to GWAS-significant loci in our study accounted for 37.5% of the total common variant heritability estimate (Fig. 5A). In contrast, the 24 definitive cardiomyopathy genes explained an approximately similar 36.2% of pLoF burden heritability in the EUR population, with TTN and other definitive cardiomyopathy genes accounting for a majority of this fraction (Figure 5B, Suppl. Tables 16, 17). We found similar patterns of enrichment in rare variant heritability within the AFR population, although this analysis was comparatively limited due to reduced sample size and power (Suppl. Fig. 11, Suppl. Fig. 12).

**Figure 5:**
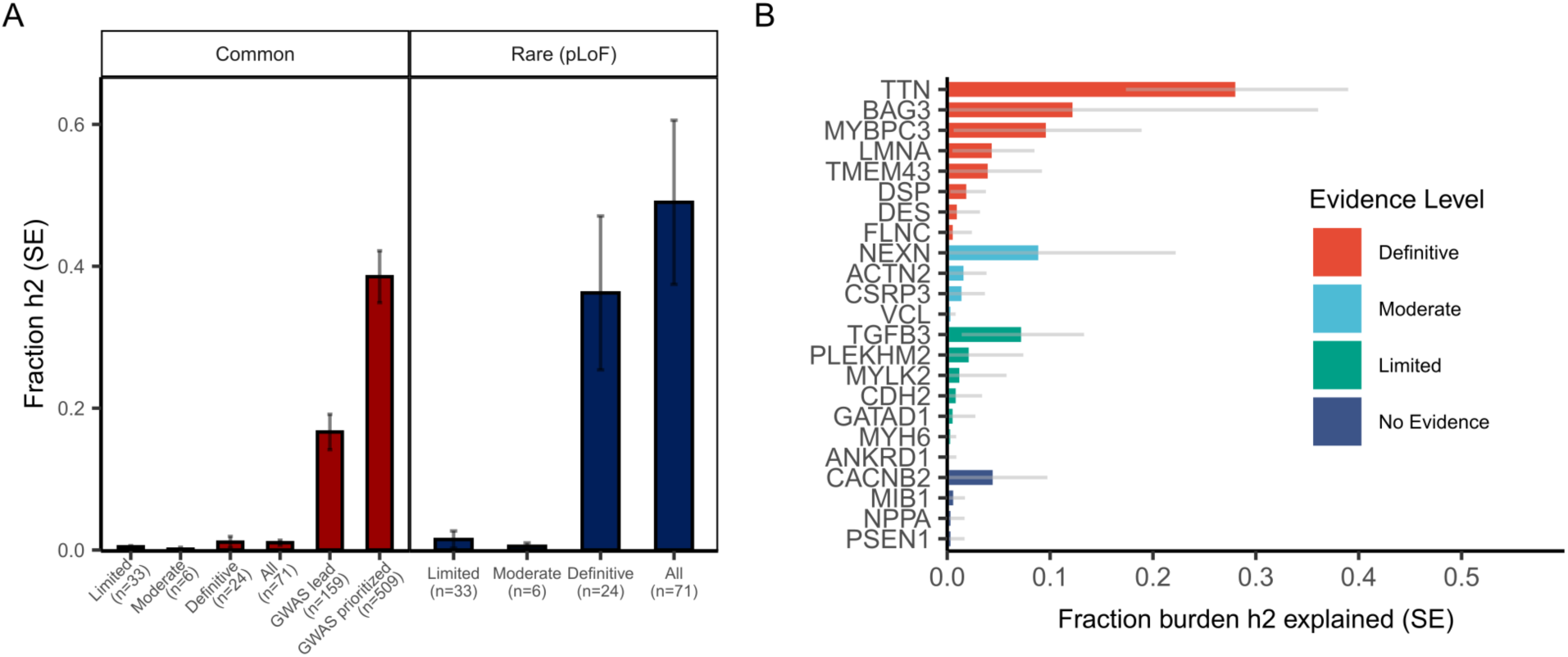
(A) Fraction of common (red) or rare (blue) burden heritability explained by known cardiomyopathy genes in HF. Fraction of common variant heritability was estimated using AMM for all cardiomyopathy gene sets, all protein coding genes nearest to GWAS significant loci (n=159), and all protein coding genes implicated by GWAS significant loci (n=509). (B) Fraction of pLoF burden heritability explained by pLoF variants for selected cardiomyopathy gene estimated to contribute >0.1% to pLoF burden heritability. See Suppl. Table 17 for full enrichment statistics.

To further characterize the relationship between GWAS-nominated and known cardiomyopathy genes, we examined interactions between these two gene sets using the STRING protein-protein interaction network^51^. We found that GWAS-prioritized genes were 1.7 and 3-fold enriched (permutation P-value < 0.0001) for protein interactions with known cardiomyopathy genes using either a relaxed or stringent threshold respectively (Suppl. Fig. 13). Within this network of interactions, we found that a broader set of sparsely connected genes implicated by common variants appear to interact with a core cardiomyopathy gene set that is much more densely connected. Indeed, the median number of interactions for GWAS-prioritized genes in this network was 3, compared to 45 for cardiomyopathy genes. Together this implies a shared genetic architecture exists between common HF and rare mendelian HF where many common variants influence HF risk both directly by affecting a set of core cardiomyopathy genes and indirectly through affecting a myriad of peripheral genes that influence and interact with this core set of cardiomyopathy genes. Among the processes implicated by the network of common and rare variant HF genes using STRING’s gene set enrichment analysis includes pathways involved in intestinal lipid absorption, cholesterol metabolism, LV systolic function, and LV diastolic function.

### Comparison of common- and rare-variant effects across populations

The prevalence of HF varies across population subgroups, including self-reported race/ethnicity^50^. Whether some of this difference is attributable to genetic variation with population-enriched effects remains controversial, and has been challenging to address due to the limited representation of non-European populations in registries and genomic studies of HF/cardiomyopathy^52,53^. Recently, population genetics studies have suggested that the effects of genetic variants are largely conserved across populations^54^; whether genetic effects are conserved across populations for HF remains unknown. To address this question, we compared the effects of common and rare HF-associated genetic variants in the AFR and EUR population groups. For common variants, we found that GWAS effect sizes are highly conserved across the two groups (Deming regression slope = 0.96, 95% CI 0.87 to 1.1, P-value = 0.5; Figure 6A). For rare variants, we similarly found that gene-collapsing effect sizes were conserved across the two groups (Deming regression slope = 0.51, 95% CI -0.12 to 1.1, P-value = 0.1; Figure 6B), however the wide confidence intervals do not exclude potentially large effects.

**Figure 6:**
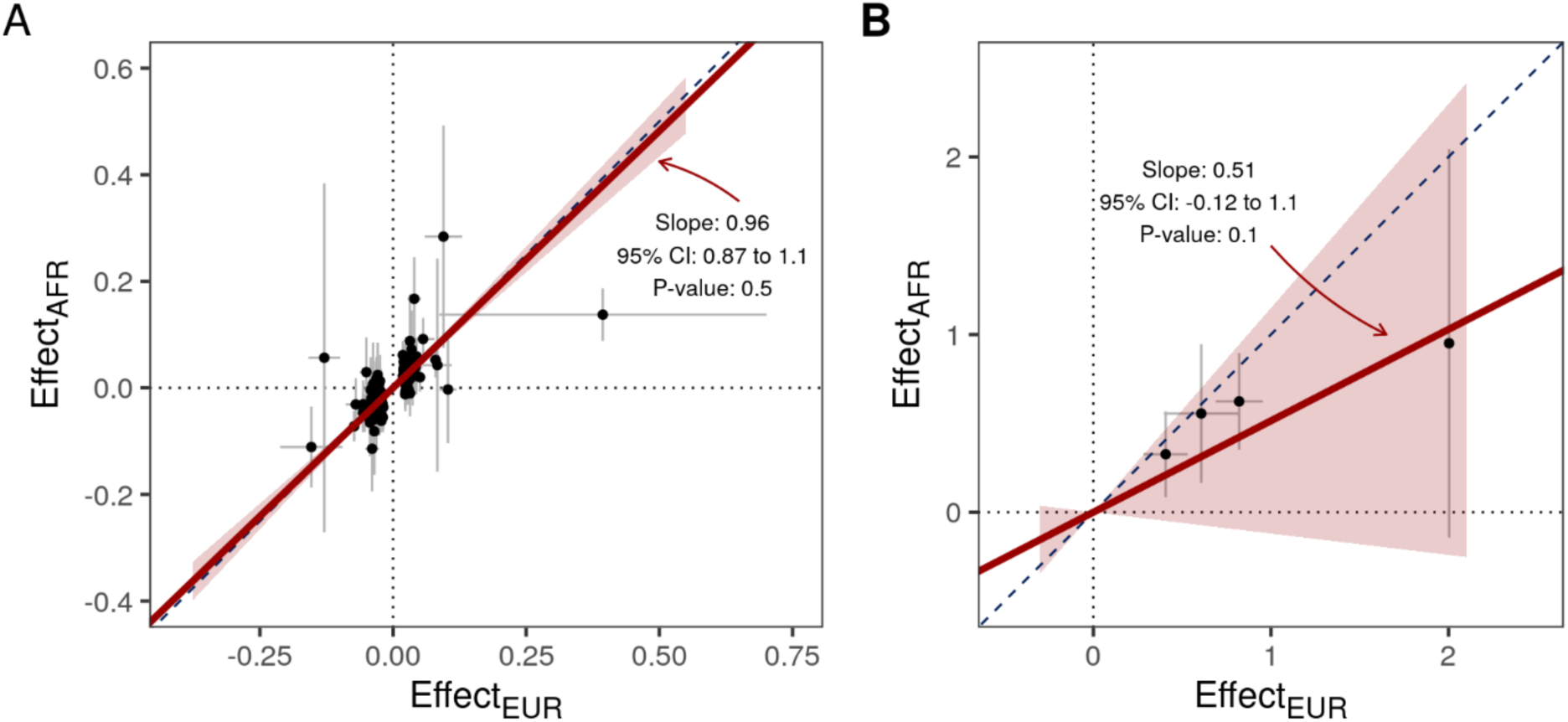
Deming regression was performed to compare the effects of common (A) and rare (B) genetic variation across European and African populations. (A) GWAS effect estimates for lead variants at the 176 genome-wide significant GWAS loci are plotted in the EUR and AFR populations (points), with 95% confidence intervals represented by black bars. The Deming regression slope (red line) and 95% confidence interval (red shaded area) demonstrate strong concordance of effects across the two populations. (B) Gene-collapsing burden effect estimates from the rare-variant exome-wide association study are plotted for the exome-wide significant pLOF and missense associations. The Deming regression slope (red line) and 95% confidence interval (red shaded area) demonstrate concordance of effects across the two populations, but with large confidence intervals.

In a complementary analysis, we leveraged the MR-MEGA meta-regression model to partition heterogeneity into population group-correlated and residual components. We observed significant population-correlated heterogeneity (adjusted P < 0.05 after Bonferroni adjustment for 107 genome-wide significant MR-MEGA signals) at 12 (11%) HF-associated loci with (Suppl. Table 2). Fewer than one locus would be expected to meet this threshold, and our findings therefore suggest enrichment of population group-correlated heterogeneity at these HF loci (one-sided binomial test P < 2.2×10^−16^). No variants demonstrated significant residual heterogeneity that may reflect differences in study designs, phenotyping, or exposures. Collectively, these results suggest that on average, common variants and rare pLoF and damaging missense variants are likely to have similar effects on risk of HF across populations, although the prevalence of disease-associated variation may vary across populations. These results are consistent with those from recent GWAS of polygenic traits^54,55^, and reinforce the need to study diverse populations to fully uncover the spectrum of HF-associated genetic variation.

### Modification of HF risk for loss-of-function *TTN* carriers by polygenic risk factors

In some cases, the etiology of HF is assessed clinically using panel-based testing for rare pathogenic variants in known cardiomyopathy genes. However, the role of testing for common variants is unclear. While individually common variants tend to have smaller effects than rare deleterious variants, in aggregate they explain an important portion of HF heritability, suggesting they may play a role in predicting HF risk. To evaluate how common variants may clinically contribute to HF risk, we generated a multi-population polygenic risk score (PRS) for HF using summary statistics from the GWAS meta-analysis (excluding PMBB participants), using PRS-CSx^56^. The PRS was normally distributed in the held-out PMBB population, and the mean PRS was significantly greater among individuals with HF than those without (Suppl. Fig. 14A). The PRS was predictive of prevalent HF in PMBB (AUC 0.72, 95% CI 0.72-0.73, including covariates – see Methods), and significantly improved upon a naïve baseline model (Brier skill score 0.065, 95% CI 0.063-0.068). Prevalence of HF ranged from 7.7% (95% CI 6.9-8.6) among the lowest PRS decile to 16% (95% CI 14.9-17.1) among the highest PRS decile (Suppl. Fig. 14B).

Finally, we asked whether polygenic risk might modify the penetrance of high PSI (>90% proportion spliced into cardiac-expressed transcripts) truncating variants in *TTN* (TTNtv), a “definitive”-evidence cardiomyopathy gene which accounts for the majority of the rare variant burden heritability of HF (Fig. 5B)^42^. The HF PRS was associated with prevalent HF among both non-carriers (OR 1.29, 95% CI 1.26-1.33) and carriers (OR 1.41, 95% CI 1.14-1.75) in the held-out PMBB population. The prevalence of HF among TTNtv carriers ranged from 8.3% (95% CI 2.2-23.6%) in the lowest PRS decile to 28.1% (95% CI 17.4% to 41.7%) among the highest decile (Suppl. Fig 15). Collectively, these results suggest that while TTNtv carriers have a higher baseline prevalence of HF, overall risk of HF depends on both carrier status and polygenic risk.

## Discussion

We present a comprehensive analysis of genetic contributions to heart failure across the allele frequency spectrum. Our study highlights the shared and unique genetic architectures of common and rare variants in HF, finding significant common and rare variant heritability, which is concentrated in different functional annotations. We characterize the contribution of known cardiomyopathy genes to HF risk across both common and rare variants, and explore the interaction between common and rare genetic variation. Overall, these findings have implications for future genetic association studies of HF, and for HF risk-stratification.

In our multi-population genome-wide meta-analysis, we identified 176 loci associated with HF, substantially increasing the number of HF risk loci compared to prior studies^2,3,57^. We find evidence of pleiotropy at many HF risk loci, revealing 5 modules of traits/diseases with shared genetic susceptibility including obesity, blood pressure, ischemic heart disease, immunologic diseases, and arrhythmia. Metabolism and nutrition are also broad processes implicated by our novel HF loci, with fine-mapped variants implicating the obesity-related genes *OR4P4* and LPL, new HF associations with loci near the insulin signaling and glucose homeostasis genes *GIPR* and *GLP1R*, and the enrichment of expression of GWAS-prioritized genes in cardiac adipocytes. Mendelian randomization has previously implicated *GLP1R* signaling in HF^58^, and clinical trials for the dual GIP/GLP-1 receptor agonist tirzepatide as a potential therapy for select patients with heart failure are ongoing^59^. Although medications targeting these mechanisms have potent effects on diabetes, glycemic traits, and obesity, our colocalization findings indicate that the HF effects at these loci may be separate from glycemic traits, raising the possibility of direct HF-specific benefits. Together, these findings lend direct genetic evidence supporting the observed association between dysregulated glucose metabolism with cardiac dysfunction in pre-clinical and animal models^60^. Although we report a significant increase in the number of HF-associated loci in our study, genes prioritized from our ensemble approach explain only roughly 17-38% of common-variant heritability, indicating that more common-variant HF loci remain undiscovered.

In parallel, we investigated rare coding variant contributions to HF. Our exome-wide association study identified significant associations for the known cardiomyopathy genes *TTN*, *MYBPC3*, *FLNC*, *BAG3*, *LMNA* and *MYH7*. Using an unbiased approach, we show that disease-causing mechanisms in cardiomyopathy genes varies depending on variant class, and that the correlation between the burden heritability of pLoF variants and predicted damaging missense variants on HF risk genome-wide is likely minimal. Rare variants in known cardiomyopathy genes explain a significant portion of burden heritability in HF, yet these genes account for less than half of overall burden heritability. We expect that larger cohorts will reveal more associated genes, but we anticipate they will contribute less to the rare variant burden heritability of HF compared to known cardiomyopathy genes.

Across both our common- and rare-variant analyses, we find that heritability of HF is enriched within a core set of cardiomyopathy genes. This finding provides empirical support for the current expert pathogenicity categorization of these genes, and supports their inclusion in cardiomyopathy testing panels^43–45^. Importantly, although current clinical cardiovascular testing panels focus on damaging coding variants in these core genes, we find that common variants at these same loci also contribute to risk of HF. In contrast to rare variants, for which most of the heritability is accounted for by these core genes, we find that common variant heritability is substantially more polygenic, and explains a significant fraction of HF heritability. In combination, these results are consistent with a polygenic model of inheritance,^61^ where a set of core genes and pathways are influenced by a much broader set of interconnected regulatory networks. Supporting this model for HF, we find significant common-variant heritability in non-coding functional annotations, while rare-variant heritability is concentrated in low-frequency, loss-of-function variation in constrained genes. These results suggest that at a population level, common genetic variation plays a significant role in risk of HF.

Finally, we observe that common and rare genetic variation act together to influence HF risk, which may have implications for HF risk stratification. Previously it had been shown that an individual’s polygenic background can modify the penetrance of pathogenic variants across several monogenic conditions^62,63^. Here, we observe that polygenic background modifies TTNtv carrier status, suggesting that risk stratification for rare pLoF variant carriers in monogenic cardiomyopathies may be improved if additional information is available regarding polygenic background. A similar relationship has previously been reported for TTNtv and a PRS for atrial fibrillation^64^, highlighting the diverse manifestations of cardiovascular disease among TTNtv carriers. Future studies are warranted to investigate the use of polygenic background to adjudicate cardiomyopathy genes with less certain levels of evidence and variants of unknown significance, as well as the role of PRS testing more broadly when assessing HF etiology and risk.

### Limitations

The results of our study should be interpreted within the context of their limitations. First, the current analysis focused on an all-cause heart failure phenotype, which may limit discovery of genetic variants that discordantly influence different forms of heart failure (eg. heart failure with reduced vs. preserved ejection fraction, dilated vs. hypertrophic vs. arrhythmogenic cardiomyopathy, ischemic vs. nonischemic cardiomyopathy). Indeed, we and others have previously observed HF risk loci with opposite directions of effect on HF endophenotypes and subtypes^3,63^. By studying an all-cause HF phenotype, the effects of these variants may be biased toward the null. Future large multi-population studies of HF subtypes and endophenotypes are necessary to identify these associations. Second, known cardiomyopathy genes increase HF risk by diverse mechanisms beyond pLoF. While we considered both pLoF and predicted damaging missense variants, domain and gene-specific expertise is likely necessary for accurate pathogenicity evaluation. Moreover, although total burden heritability includes both pLoF and predicted damaging missense variants, BHR tends to underestimate the contribution from missense variants unless all variants for a given gene have the same direction of effect and magnitude, or are perfectly weighted to account for differences. As such our total burden heritability estimate likely represents a lower-bound of rare coding variant heritability. Third, although the prevalence of HF is similar across sexes, whether specific loci affect HF through sex-specific mechanisms requires future study^65^. Finally, despite our attempts to include participants of diverse genetic backgrounds, the study was still predominantly comprised of individuals genetically similar to European reference populations (Fig. 1), although the inclusion of participants from the VA Million Veteran Program (MVP) and All of Us significantly expanded the genetic diversity of our common and rare variant analyses compared to previous publications. We hope that future studies continue to expand on the diversity of participants included from historically underrepresented populations.

In summary, these analyses highlight similarities and differences in the genetic architecture of HF across the allele-frequency spectrum, which is characterized by a highly concentrated component of heritability in rare deleterious variants in known cardiomyopathy genes, and a large polygenic component of heritability that is more diffusely spread across GWAS loci and enriched for non– coding variation. Our analysis of common variants in HF uncovers 176 HF loci of which 105 have not previously been published at genome-wide significance, and our analysis of pleiotropy mediated by these loci reveals five distinct modules of traits/diseases with shared genetic susceptibility with HF. Finally, we find that both common and rare variants contribute significantly to HF risk and that even for carriers of rare, highly penetrant TTN truncating mutations there may be additional utility in assessing polygenic HF risk based on common variants.

## Methods

### Genome-wide meta-analysis of heart failure

GWAS summary statistics for HF were obtained from non-overlapping analyses of 8 separate cohorts/consortia (HERMES, VA MVP, FinnGen, Mount Sinai BioMe, Penn Medicine Biobank, eMERGE, Geisinger DiscovEHR, and the Global Biobank Meta-analysis Initiative). All-cause HF was defined using cohort-specific definitions (pheCodes or ICD9/10 codes documented within the electronic health record for all studies except HERMES, which additionally included expert adjudication among some cohorts).

Analyses were performed separately by cohort and population, adjusted for age, sex, and population structure. Fixed-effects meta-analysis was performed using METAL^66^ using the inverse-variance weighted (standard error) method within and across populations to generate population-specific and multi-population meta-analysis summary statistics. Fine-grained population estimation from summary statistics was performed using *bigsnpr*^67^. Additional association statistics were generated using MR-MEGA, accounting for 4 genetic principal components^68^. Independent significant genomic risk loci were defined by grouping genome-wide significant variants (P-value < 5×10^−8^) into 1MB clusters using the *genome_cluster* function of the *tidygenomics* package in R^69^. To minimize spurious associations, loci were required to have at least two variants achieving the genome-wide significance threshold. Lead variants at each locus were defined by the variant with the lowest P-value, present in at least two studies.

### Gene Prioritization and Identification of variants within the 99% credible set

Credible sets were generated surrounding the index variant at each 1MB locus identified in the multi-population meta-analysis, centered on the index variant (+/-500kb). Credible sets were constructed by first estimating approximate Bayes factors as described in Graham et. al, which assumes a single causal variant per locus^70^. Posterior probabilities were then generated by dividing the Bayes factor for each variant at a locus by the sum of Bayes factors at each locus^68^. Finally, variants at each locus were ranked in descending order of posterior probability and included until the cumulative posterior probability at each locus attained 99%. We performed additional analysis with CARMA to identify secondary signals (Suppl. Table 8). Possible protein-coding consequences were annotated for each variant in this 99% credible set using Ensembl VEP^71^. TWAS was performed using S-MultiXcan^72^ using GTEx tissue expression data while additional gene candidates were identified from GWAS loci using MAGMA^73^ following the standard protocol, with statistical significance determined using a Bonferroni correction for multiple testing (0.05/# of genes in each TWAS or MAGMA analysis). Gene set enrichment analysis was performed using FUMA^74^ for tissue-specific enrichments. To identify cell-type specific enrichment, processed single-cell expression data was downloaded from GTEx v8^38^. Cell-type specific expression profiles were constructed by taking median expression values across samples for each annotated cell-type, and then enrichment for cell-type specific expression was determined by assessing the number of prioritized GWAS genes that were expressed by each cell-type using the hypergeometric test. Prioritized genes were assessed for enrichment in DrugBank targets by matching DrugBank target IDs to Ensembl gene IDs and testing for overrepresentation by Fisher’s Exact Test.

### Pleiotropy and Multi-trait colocalization

Assessment of shared loci in the NHGRI-EBI catalogue using the February 11, 2024 release. Traits were clustered based on the number of shared rsIDs with lead HF loci (Suppl. Fig. 5). Multi-trait colocalization was performed using HyPrColoc to evaluate the presence of a shared causal variant influencing HF traits/diseases in the UK Biobank^25^. This Bayesian algorithm is designed to evaluate for the presence of a causal variant shared among several traits, under the single causal variant assumption. Summary genetic associations for >2000 traits/diseases (phecodes, biomarkers, and continuous measurements) were obtained from the Pan-UK Biobank project. At each of the 176 heart failure risk loci, multi-trait colocalization was performed simultaneously for heart failure and all other traits with a genome-wide significant association within +/-500kb of the lead HF risk variant. In an additional focused analysis, GWAS summary statistics for diabetes^5^, hemoglobin A1c^36^, and body mass index^37^ were queried at the *GLP1R* and *GIPR* loci to identify variants +/-500kb of the lead variant for HF at each locus. Because the diabetes, A1c, and BMI GWAS were performed among European populations, the European population GWAS of heart failure was used to minimize differences in colocalization due to linkage disequilibrium. Evidence for colocalization was determined based on the default variant specific regional and alignment priors, with colocalization determined with posterior probability >= 0.25, determined by the authors of HyPrColoc based on simulations maximizing true positive rate and minimizing false positive rate^25^.

### Pleiotropy Network

Among HF loci with high-confidence evidence of colocalization (posterior probability > 0.7), a bipartite network was created linking each locus with each colocalized trait/disease using the *tidygraph*^75^ package in R. A projection of this network was then created to generate a disease-disease network including traits/diseases that were connected by colocalization at multiple HF risk loci. Among this disease-disease network, Louvain community detection was performed to identify modules of distinct communities.

### Curation of Cardiomyopathy Genes

Cardiomyopathy genes for dilated and hypertrophic cardiomyopathies were selected from two previous publications from the NIH ClinGen working group and classified according to the highest level of evidence supporting each gene across DCM, HCM, or ARVC collectively. We focused this analysis on genes from Ingles et al.^43^ for hypertrophic cardiomyopathy, Jordan et al. for dilated cardiomyopathy^44^, and James et al. for arrhythmogenic cardiomyopathy^45^. Except for *FHL1*, *ALPK3*, and *PLN*, genes involved in syndromic HCM were not included in our set of 71 cardiomyopathy genes.

### Rare variant association analysis

Sample preparation, exome sequencing, and variant QC of participants in the Penn Medicine Biobank have been described previously^76^. We further processed variants to remove variants flagged during joint variant calling based on GATK’s hard-filtering thresholds^77^. When >10% of variant alternate calls across PMBB participants at a particular site were flagged for QC these variant sites were removed from the analysis. Otherwise, flagged variants composing less than 10% of all alternate calls at a variant site were changed to missing. Variants from the UKB and AOU cohorts were processed as previously published^78,79^.

Variant annotation from PMBB, UKB, and AOU was performed using Ensembl VEP^71^. Damaging missense variants for each gene were identified using publicly available REVEL scores using a threshold of > 0.5^80^. For predicted loss-of-function (pLoF) variants (essential splice site disrupting, stop gain, or frameshift variants) annotated by Ensembl VEP, we further restricted analysis to the subset of variants passing all LOFTEE filters to produce a filtered high quality set of pLoF variants^81^. Rare variant association studies were performed using SAIGE-GENE^40^ for individuals of European or African populations separately, adjusting for age, sex, and the first 10 genetic principal components. Variants were considered rare if MAF < 0.1% in gnomAD exomes (without consideration for super-population maximum allele frequencies) and in each respective cohort. P-values were derived from a fixed-effects meta-analysis of EUR and AFR population effect size estimates for PMBB, UKB, and AOU for each gene after adjustment for case-control imbalance in each respective cohort using Firth’s logistic regression. pLoF and damaging missense variants were analyzed separately, amounting to 31,857 tests, corresponding to Bonferroni-adjusted significance thresholds of 1.57×10^−6^. Unless otherwise noted, all other rare variant studies in the present study relied on pLoF variant annotations produced from this analysis. Odds ratios for HF among pLoF or damaging missense carriers in the subset of 71 curated cardiomyopathy genes were derived from meta-analysis effect size estimates.

### Estimation of common variant heritability and enrichment

Common variant heritability was estimated using LD score regression (LDSC)^82,83^ for each population separately in the HF GWAS meta-analysis using reference panels published by the Pan-UK Biobank.

To determine heritability enrichment and fraction of common variant heritability distributed across different functional categories we used stratified LD-score regression^84^ and precomputed baseline 2.2 model LD scores available <u>online here</u>. Since pre-computed LD scores are not available for African populations, we generated custom LD scores using bed files corresponding to the baseline 2.2 model (<u>online here</u>) to estimate heritability enrichment across functional categories using summary statistics derived from the African population group GWAS fraction.

To estimate the enrichment and fraction of common variant heritability mediated through specific gene sets, we used the Abstract Mediation Model with default parameters^10^. Briefly, for analysis of cardiomyopathy genes, we separated cardiomyopathy genes into sets based on their level of supporting evidence as determined by expert review, and assessed each set separating for common variant heritability enrichment and the fraction of overall common variant heritability explained. To determine the enrichment of common variant heritability explained by all genome-wide significant loci, we used the set of nearest mapped protein coding genes for each loci. Of 176 independent genome-wide significant loci, we were able to identify a nearest protein-coding gene (as annotated by Ensembl VEP) for 516 genes prioritized through our ensemble approach from 159 loci that could be used in our estimate of common variant heritability mediated through various GWAS-significant gene sets. To calculate the fraction of common variant heritability mediated through these genes we multiplied their estimated enrichment in heritability as determined by AMM by the fraction of genes in each gene set respectively divided by the total number of genes included in the AMM model.

### Estimation of rare variant heritability and enrichment

To estimate rare variant burden heritability, we used Burden Heritability Regression (BHR)^11^. BHR was used to estimate heritability statistics for EUR and AFR population participants from AOU, PMBB, and UKB separately. These estimates were combined using a fixed-effects meta-analysis to produce a combined estimate for total burden heritability, and other associated heritability statistics presented in the study. Only variants annotated within protein coding exons on autosomes were used for this analysis. Rare (AF <0.001%) pLoF and damaging missense variants were combined to estimate total burden heritability. To estimate rare variant heritability enrichment across cardiomyopathy genes, we supplied BHR with separate annotation lists of cardiomyopathy gene IDs in addition to the baseline model. Estimates of fraction burden heritability attributable to specific genes were included in the meta-analysis only if those genes contained at least five qualifying rare variants in each respective study.

Carriers of predicted loss-of-function variants in TTN affecting high-percent spliced in (PSI) exons were identified in the PMBB, and an interaction effect between hi-PSI TTNtv carrier status and HF polygenic risk was tested for among EUR and AFR participants of PMBB. The multi-population PRS was constructed using the “auto” setting of PRS-CSx^56^, using population-specific LD reference panels from the UK Biobank. Classification performance for prevalent HF was assessed in PMBB using repeated cross validation (3 repeats of 10 folds), using the area under the receiver operator characteristic curve as a measure of discrimination, and using the Brier skill score as an integrated measure of discrimination and calibration in comparison to a naïve/no-skill baseline model^85^. TTNtv carrier status was included in a logistic model with the multi-population PRS derived from the multi-population GWAS using HF (defined as 2 instances of phecode 428) as an outcome, including an interaction term between the PRS and TTNtv carrier status. All PRS models were adjusted for age, sex, and 5 genetic principal components, with PRS scaled/centered within each target population (EUR or AFR) prior to modeling.

## Data Availability

Summary statistics from the genome-wide association study meta-analysis and rare-variant gene-burden association study of all-cause heart failure will be made available upon publication. Polygenic risk scores for heart failure will be deposited in the Polygenic Score Catalog.

## Code Availability

Publicly available software was used to perform the analyses. Code is available from the corresponding author upon reasonable request.

## Acknowledgements

We thank the participants of the Penn Medicine Biobank and other cohorts/consortia contributing to this analysis. The PMBB is supported by Perelman School of Medicine at University of Pennsylvania, a gift from the Smilow family, and the National Center for Advancing Translational Sciences of the National Institutes of Health under CTSA award number UL1TR001878. D.S.M.L. was supported by NIH Medical Scientist Training Program T32GM007170. M.G.L. was supported by the Institute for Translational Medicine and Therapeutics of the Perelman School of Medicine at the University of Pennsylvania, the NIH/NHLBI National Research Service Award postdoctoral fellowship (T32HL007843), the Measey Foundation, and Doris Duke Foundation (Award 2023-2024). S.M.D. was supported by the US Department of Veterans Affairs Clinical Research and Development Award IK2-CX001780. This publication does not represent the views of the Department of Veterans Affairs or the United States Government.

This research has been conducted using the UK Biobank Resource under Application Number 32133.

We acknowledge the VA Million Veteran Program (MVP) and the VA-DOE genome-wide PheWAS core analytic team for generating the MVP summary statistics that were used in this manuscript^86^.

The *All of Us* Research Program is supported by the National Institutes of Health, Office of the Director: Regional Medical Centers: 1 OT2 OD026549; 1 OT2 OD026554; 1 OT2 OD026557; 1 OT2 OD026556; 1 OT2 OD026550; 1 OT2 OD 026552; 1 OT2 OD026553; 1 OT2 OD026548; 1 OT2 OD026551; 1 OT2 OD026555; IAA #: AOD 16037; Federally Qualified Health Centers: HHSN 263201600085U; Data and Research Center: 5 U2C OD023196; Biobank: 1 U24 OD023121; The Participant Center: U24 OD023176; Participant Technology Systems Center: 1 U24 OD023163; Communications and Engagement: 3 OT2 OD023205; 3 OT2 OD023206; and Community Partners: 1 OT2 OD025277; 3 OT2 OD025315; 1 OT2 OD025337; 1 OT2 OD025276. In addition, the All of Us Research Program would not be possible without the partnership of its participants.

