## Supplemental Text and Figures for "Common- and rare-variant genetic architecture of heart failure across the allele frequency spectrum"

Study Populations, Phenotyping, Genotyping, and Quality Control

HERMES Consortium: Details of the HERMES Consortium have been previously described[1]. Briefly, the HERMES Consortium included participants of European ancestry from 26 separate cohorts or population-based studies including the UK Biobank. Heart failure was defined based on study-specific criteria, including diagnosis codes, discharge codes, death certificate codes, and expert clinician evaluation. Genotyping was performed using study-specific high density genotyping arrays, and quality controls included sample/variant call rate, Hardy-Weinberg equilibrium, and minor allele frequency, with additional study-specific quality controls. Imputation was performed to a variety of imputation panels including 1000 Genomes and Haplotype Reference Consortium. Genome wide association studies were adjusted for covariates including age, sex, and genetic principal components where available. Summary level quality control was performed using EasyQC, prior to fixed-effects meta-analysis using METAL[2].

Penn Medicine BioBank: The Penn Medicine Biobank is a longitudinal genomics and precision medicine study in which participants consent to linkage of genomic information and biospecimens to the electronic health record. All patients receiving care at Penn Medicine (Philadelphia, PA) are eligible for enrollment, and more than 65,000 participants are currently enrolled. Heart failure was defined based on diagnosis codes documented within the electronic health record. International Classification of Diseases version 9/10 codes were mapped to pheCodes[3], and individuals with codes 425 or 428 (including subcodes) were considered heart failure cases. Individuals without these codes were considered controls. Genotyping was performed using the Infinium Global Screening Array (GSA) (Illumina, Inc. San Diego, CA), and the current analysis included 43,623 participants with available genotype data. Pre-imputation quality control was performed using PLINK[4,5] to exclude low marker call rate (<95%), low sample call rate (<90%), or sex discordance between genotype and reported sex. Imputation to the TOPMed reference panel (97,256 samples and 308,107,085 variants) was performed using the TOPMed Imputation Server, with phasing performed using EAGLE and imputation performed using MINIMAC4[6]. Identity-by-descent with a Pi-hat threshold of 0.25 to account for relatives up-to first cousins was performed on the imputed genotype data, and one sample per related pair was removed. Ancestry-specific GWAS (European and African Ancestry) were performed using PLINK[4,5] among variants meeting the following quality thresholds: $r2 > 0.3$; minor allele frequency > 0.001 or minor allele count > 20, adjusted for age, sex, and 5 ancestry-specific genetic principal components. LiftOver was used to map genome positions from hg38 to hg19/GRCh37[7].

eMERGE: The genome-wide association study (GWAS) was conducted using the electronic Medical Records and Genomics (eMERGE) Network dataset. The case/control cohort was comprised of individuals 18 years or older from the following non-pediatric eMERGE sites:

Marshfield Clinic, Vanderbilt University Medical Center, Kaiser Permanente/University of Washington Medical Center, Columbia University, Mayo Clinic, Northwestern University, Geisinger Health System, Harvard (Partners Health Care), Icahn School of Medicine at Mount Sinai, and Meharry Medical College. Cases were defined as having an occurrence of one or more of the following International Classification of Diseases (ICD-9 or ICD-10) codes: I11.0, I13.0, I13.2, I25.5, I42.0, I42.5, I42.8, I42.9, I50.0, I50.1, I50.9, 425.4, 428.0, 428.1, 428.9, 402.01, 402.11, 402.91, 404.01, 404.11, 404.91, 404.03, 404.13, 404.93, 425.4, I50.0, I50.1, I50.2, I50.4, I50.9, or/and I50.82. Controls were defined as any individuals not having any of the aforementioned ICD codes. We conducted ancestry-specific analyses, creating separate case/control cohorts for individuals self-identifying as "Black" and "White" in the "Race" variable in the eMERGE dataset. There were a total of 7,208 controls and 2,607 cases in the African cohort (total = 9,815 individuals), and 48,714 controls and 14,065 cases in the European cohort (total = 62,779 individuals). Genotyping and quality control of eMERGE has been previously reported[8]. Imputed genotype data with a minor allele frequency threshold of 0.01 to conduct the GWAS. Plink-v2.04,5 was used for all analysis. For the quality control of the genetic data we checked for robust sex concordance, a marker call rate of 0.01, and a sample call rate of 0.01. Related individuals were dropped using a pihat > 0.25 threshold. A Hardy-Weinberg equilibrium p-value threshold of 1e-9 was used. This produced a total of 13,275,706 SNPs tested in the African cohort and 7,654,263 SNPs tested in the European cohort. Logistic regression was run using covariates sex, age and the first 5 principal components (PCs). PCs were calculated from the ancestry-specific cohorts.

Mount Sinai BioMe: BioMe is an electronic health record-linked clinical-care biobank which includes more than 45,000 participants of diverse ancestry. Participants are recruited from the Mount Sinai healthcare system (New York, NY). The current analysis included individuals of European or African self-reported race/ancestry to avoid overlapping the Global Biobank Meta-analysis Initiative dataset which included Hispanic individuals from BioMe. Individuals with heart failure were identified using electronic health record diagnosis codes. Individuals were considered heart failure cases if they had evidence of the following codes: ICD10: I11.0, I13.0, I13.2, I25.5, I42.0, I42.5, I42.8, I42.9, I50.0, I50.1, I50.9; ICD9: 4254, 4280, 4281, 4289. Genotyping was performed using the Global Screening Array (Illumina, Inc. San Diego, CA). Genotype quality controls included checks for sex discordance, sample duplicates, low call rate (<95%), Hardy-Weinberg Equilibrium (p < 1x10-5Individuals with closer than second-degree relatedness were removed using KING9. Genotypes were imputed using the TOPMed imputation server (https://imputation.biodatacatalyst.nhlbi.nih.gov). Ancestry-specific GWAS were performed using PLINK version 2 among well-imputed variants with minor allele frequency > 0.01, adjusting for age, sex and 10 genetic principal components[4,5]. LiftOver was used to map genome positions from hg38 to hg19[7].

Geisinger DiscovEHR: DiscovEHR is a collaboration between Geisinger and the Regeneron

Genetics Center. The population is derived from patients who have previously consented to participate in the Geisinger MyCode Community Health Initiative. MyCode is an IRB-approved research study, and all participants have provided informed consent for broad use of samples for research. Participants are broadly recruited to MyCode from both primary and specialty care clinics across the Geisinger system. Exome sequencing and genome-wide genotyping of samples are performed by Regeneron, and genomic data are linked with Geisinger's long-standing electronic health record, comprising both inpatient and outpatient records. This study included data from 82,608 patients who were genotyped on the Illumina Global Screening Array chip out of 144,204 total patients in the cohort. Heart failure status was assigned based on ICD-10 codes. Details of genotyping and quality control have been previously reported[9]. The DiscovEHR participants included in the current analysis were distinct from participants included in the previously-published HERMES GWAS.

Global Biobank Meta-analysis Initiative: The Global Biobank Meta-analysis Initiative (GBMI) is a network of 19 biobanks representing >2 million consenting participants, with linkage of electronic health record and genotype data. Details of phenotyping, genotyping, quality control, and GWAS in GBMI have been reported previously[10]. To avoid overlap with other datasets included elsewhere in the current meta-analysis, we included two GWAS from GBMI (admixed-American and East Asian ancestry studies). Individuals were considered heart failure cases based on pheCode or ICD codes recorded within electronic health records, using study-specific definitions adapted from a GBMI-recommended definition. Genotyping was performed individually by biobank using genotype arrays. Following standard sample- and variant-level quality control genotypes were imputed into reference panels including 1000 Genomes, Haplotype Reference Consortium, or TOPMed. Ancestry-specific GWAS were performed by study, with suggested covariates including age, age2, sex, age*sex, 20 first principal components, and biobank specific covariates including genotyping batches and recruiting centers. GWAS were recommended to be performed using SAIGE or REGENIE[11,12]. LiftOver was used to map genome positions from hg38 to hg19[7].

FinnGen: FinnGen is a public-private partnership aiming to collect genome and health data on 500,000 Finnish biobank participants. The study consists of ~200,000 legacy samples primarily collected by the National Institute for Health and Welfare, and an additional ~300,000 samples to be prospectively collected from hospital biobanks. Participating individuals consent to linkage of genome-wide genotyping with nationwide registers of longitudinal health data. Individuals with heart failure were identified using inpatient, outpatient, insurance reimbursement, and medication records, based on the "I9_HEARTFAIL_ALLCAUSE" phenotype (https://risteys.finngen.fi/phenocode/I9_HEARTFAIL_ALLCAUSE). Details of genotyping and quality control are available from https://finngen.gitbook.io/documentation/. Briefly, individuals underwent genotyping using Illumina or Affymetrix chip arrays. Individuals with ambiguous gender, high genotype missingness (>5%), excess heterozygosity (+/- 4 standard deviation), and non-Finnish ancestry were excluded. Variants with high-missingness, low Hardy-Weinberg Equilibrium p-value ) <1 x 10-6), and minor allele count <3 were excluded. Samples were pre-phased using EAGLE[13], and imputed to the SISu v3 imputation reference panel using BEAGLE 4.115. LiftOver was used to map genome positions from hg38 to hg19[7]. FinnGen participants provided informed consent for biobank research, and the Coordinating Ethics Committee of the Hospital District of Helsinki and Uusimaa (HUS) approved the FinnGen Study protocol No. HUS/990/2017.

VA Million Veteran Program (replication): Details of the VA Million Veteran Program (MVP) have been previously described[14,15]. Briefly MVP recruits participants from the Department of Veterans Affairs Healthcare System, who consent to linkage of electronic health records with biospecimens, surveys, and genomic information. More than 850,000 participants have enrolled, with genomic and electronic health record data currently available for approximately 650,000. HF phenotyping has been previously described, and was based on a combination of structured (ICD codes), and unstructured (ejection fraction) data extracted from the electronic health record[15]. Participants underwent genotyping using a custom Affymetrix Axiom Biobank Array[14]. Genotyping quality control has been previously described, and excluded duplicate samples, samples with more heterozygosity than expected, missing genotype calls (>2.5%), or discordance between genetically inferred sex and phenotypic gender[16]. One individual from each pair of related individuals (more than second degree relatedness as determined by KING[17]) was excluded. Variants were imputed to the 1000 Genomes Phase 3 version 5 reference panel using MINIMAC46. Following imputation, low-quality (r2<0.3) were excluded from further analysis. Ancestry was assigned using HARE as previously described19. PLINK2[4,5] was used to test for associations between each common (minor allele frequency >0.01) directly measured or imputed variant and all-cause heart failure, adjusted for age, sex, and ten genetic principal components.

Regeneron Genetics Center Banner Author List and Contribution Statements

**RGC Management and Leadership Team**
Goncalo Abecasis, PhD , Aris Baras, M.D. , Michael Cantor, M.D. , Giovanni Coppola, M.D. , Andrew Deubler , Aris Economides, Ph.D. , Luca A. Lotta, M.D., Ph.D. , John D. Overton, Ph.D. , Jeffrey G. Reid, Ph.D. , Katherine Siminovitch, M.D. , Alan Shuldiner, M.D.

**Sequencing and Lab Operations**
Christina Beechert , Caitlin Forsythe, M.S. , Erin D. Fuller , Zhenhua Gu, M.S. , Michael Lattari , Alexander Lopez, M.S. , John D. Overton, Ph.D. , Maria Sotiropoulos Padilla, M.S. , Manasi Pradhan, M.S. , Kia Manoochehri, B.S. , Thomas D. Schleicher, M.S. , Louis Widom , Sarah E. Wolf, M.S. , Ricardo H. Ulloa, B.S.

**Clinical Informatics**
Amelia Averitt, Ph.D. , Nilanjana Banerjee, Ph.D. , Michael Cantor, M.D. , Dadong Li, Ph.D. , Sameer Malhotra, M.D. , Deepika Sharma, MHI , Jeffrey Staples , Ph.D.

**Genome Informatics**
Xiaodong Bai, Ph.D. , Suganthi Balasubramanian, Ph.D. , Suying Bao, Ph.D. , Boris Boutkov, Ph.D. , Siying Chen, Ph.D. , Gisu Eom, B.S. , Lukas Habegger, Ph.D. , Alicia Hawes, B.S. , Shareef Khalid , Olga Krasheninina, M.S. , Rouel Lanche, B.S. , Adam J. Mansfield, B.A. , Evan K. Maxwell, Ph.D. , George Mitra, B.A. , Mona Nafde, M.S. , Sean O'Keeffe, Ph.D. , Max Orelus, B.B.A. , Razvan Panea, Ph.D. , Tommy Polanco, B.A. , Ayesha Rasool, M.S. , Jeffrey G. Reid, Ph.D. , William Salerno, Ph.D. , Jeffrey C. Staples, Ph.D. , Kathie Sun, Ph.D.

**Analytical Genomics and Data Science**
Goncalo Abecasis, D.Phil. , Joshua Backman, Ph.D. , Amy Damask, Ph.D. , Lee Dobbyn, Ph.D. , Manuel Allen Revez Ferreira, Ph.D. , Arkopravo Ghosh, M.S. , Christopher Gillies, Ph.D. , Lauren Gurski, B.S. , Eric Jorgenson, Ph.D. , Hyun Min Kang, Ph.D. , Michael Kessler, Ph.D. , Jack Kosmicki, Ph.D. , Alexander Li , Ph.D. , Nan Lin, Ph.D. , Daren Liu, M.S. , Adam Locke, Ph.D. , Jonathan Marchini, Ph.D. , Anthony Marcketta, M.S. , Joelle Mbatchou, Ph.D. , Arden Moscati, Ph.D. , Charles Paulding, Ph.D. , Carlo Sidore, Ph.D. , Eli Stahl, Ph.D. , Kyoko Watanabe, Ph.D. , Bin Ye, Ph.D. , Blair Zhang, Ph.D. , Andrey Ziyatdinov, Ph.D.

**Therapeutic Area Genetics**
Ariane Ayer, B.S. , Aysegul Guvenek, Ph.D. , George Hindy, Ph.D. , Giovanni Coppola, M.D. , Jan Freudenberg, M.D. , Jonas Bovijn M.D. , Katherine Siminovitch, M.D. , Kavita Praveen, Ph.D. , Luca A. Lotta, M.D. , Manav Kapoor, Ph.D. , Mary Haas, Ph.D. , Moeen Riaz , Ph.D. , Niek Verweij, Ph.D. , Olukayode Sosina, Ph.D. , Parsa Akbari, Ph.D. , Priyanka Nakka, Ph.D. , Sahar Gelfman, Ph.D. , Sujit Gokhale, B.E. , Tanima De, Ph.D. , Veera Rajagopal, Ph.D. , Alan Shuldiner, M.D. , Bin Ye, Ph.D. , Gannie Tzoneva, Ph.D. , Juan Rodriguez-Flores, Ph.D.

**Research Program Management & Strategic Initiatives**
Esteban Chen, M.S. , Marcus B. Jones, Ph.D. , Michelle G. LeBlanc, Ph.D. , Jason Mighty, Ph.D. , Lyndon J. Mitnaul, Ph.D. , Nirupama Nishtala, Ph.D. , Nadia Rana, Ph.D. , Jaimee Hernandez

### Supplemental Figures

#### Table of Contents


| Page(s) | Figure # | Title |
|---|---|---|
| 7-38 | 1 | Regional association plots |
| 40 | 2 | Overlap between lead loci idenfied by METAL and MR-MEGA |
| 41 | 3 | Fine-scale inference of ancestry proportions using summary statistics |
| 42 | 4 | Q-Q plot of p-value distribution from multi-population GWAS meta analysis |
| 43 | 5 | Clustering shared rsIDs from NHGRI-EBI GWAS Catalog |
| 44 | 6 | Credible set sizes for multi-population analysis and each ancestry separately |
| 45 | 7 | FUMA: Hallmark gene set enrichment analysis |
| 45 | 8 | FUMA: GWAS catalog enrichment analysis |
| 46 | 9 | Association of predicted damaging missense or high-confidence loss-of-function variants in known cardiomyopathy genes with all-cause HF |
| 47 | 10 | Common variant h2 estimates from HF GWAS across individuals of African, Admixed American, East Asian, and European populations. |
| 48 | 11 | Burden h2 (liability scale) for PMBB AFR population across coding consequence and AF bin categories |
| 49 | 12 | Fraction burden heritability explained by known cardiomyopathy genes in AFR HF participants |
| 50 | 13 | StringDB GWAS-Cardiomyopathy Interaction Network and Enrichment |
| 51 | 14 | HF PRS distributions among PMBB participants |
| 52 | 15 | Log-odds of HF in the PMBB for TTNtv carriers as a function of multiancestry PRS Z-score scaled separately by ancestry for EUR and AFR populations |

**Suppl. Fig. 1.** Regional association plots for each genome-wide significant locus in the multi-ancestry meta-analysis. Linkage disequilibrium information was obtained from the University of Michigan LocusZoom API via the locusplotr package. For multiancestry analysis a cosmopolitan LD panel ("ALL") was used.

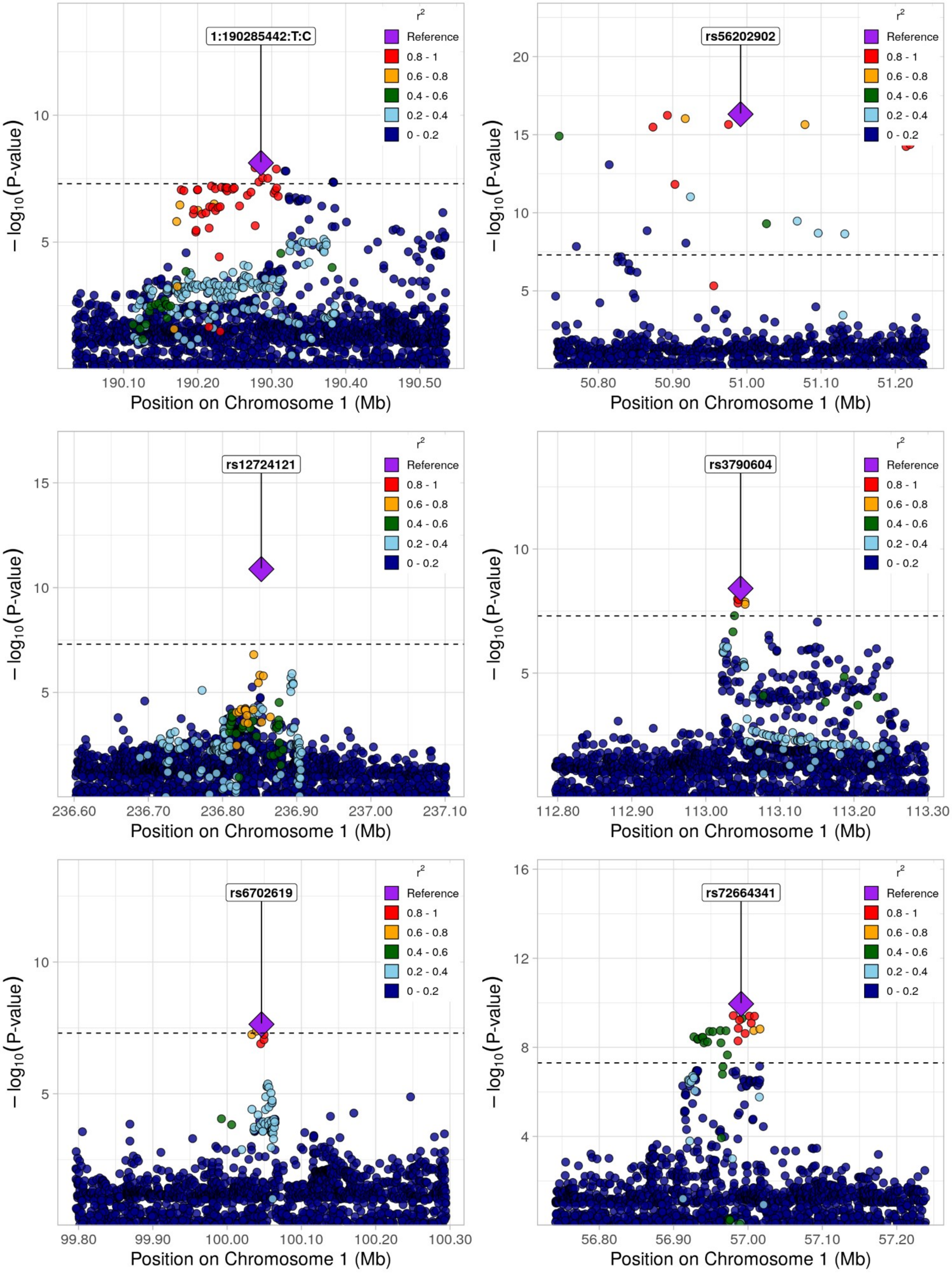

1:190285442:T:C
rs56202902
rs12724121
rs3790604
rs6702619
rs72664341
r²
Reference
0.8 - 1
0.6 - 0.8
0.4 - 0.6
0.2 - 0.4
0 - 0.2
− log10(P-value)
Position on Chromosome 1 (Mb)

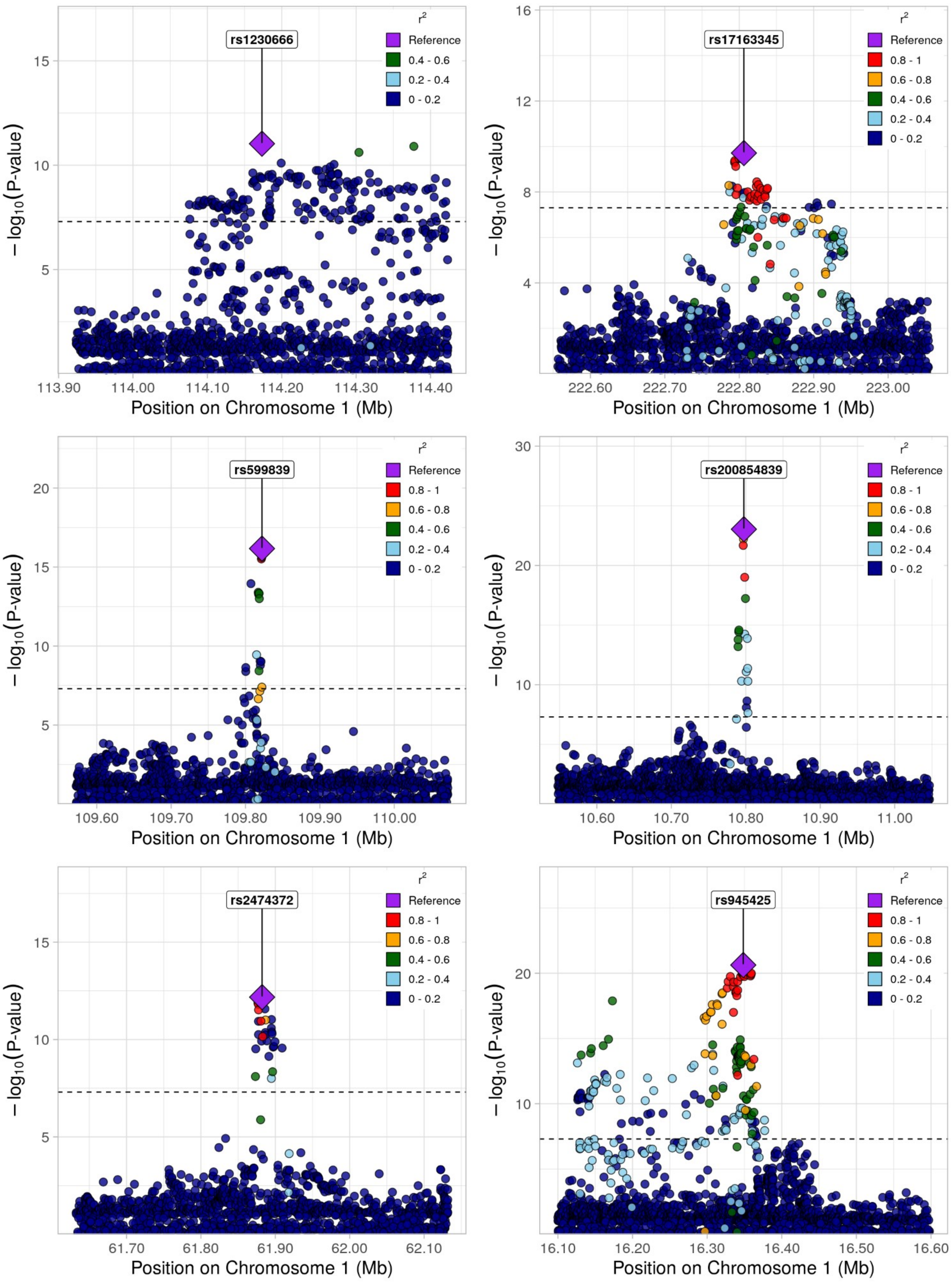

rs1230666
r2
Reference
0.4 - 0.6
0.2 - 0.4
0 - 0.2
15
10
5
113.90
114.00
114.10
114.20
114.30
114.40
Position on Chromosome 1 (Mb)
− log10(P-value)
rs17163345
Reference
0.8 - 1
0.6 - 0.8
0.4 - 0.6
0.2 - 0.4
0 - 0.2
16
12
8
4
222.60
222.70
222.80
222.90
223.00
Position on Chromosome 1 (Mb)
− log10(P-value)
rs599839
20
15
10
5
109.60
109.70
109.80
109.90
110.00
Position on Chromosome 1 (Mb)
− log10(P-value)
rs200854839
30
20
10
10.60
10.70
10.80
10.90
11.00
Position on Chromosome 1 (Mb)
− log10(P-value)
rs2474372
15
10
5
61.70
61.80
61.90
62.00
62.10
Position on Chromosome 1 (Mb)
− log10(P-value)
rs945425
20
10
16.10
16.20
16.30
16.40
16.50
16.60
Position on Chromosome 1 (Mb)
− log10(P-value)

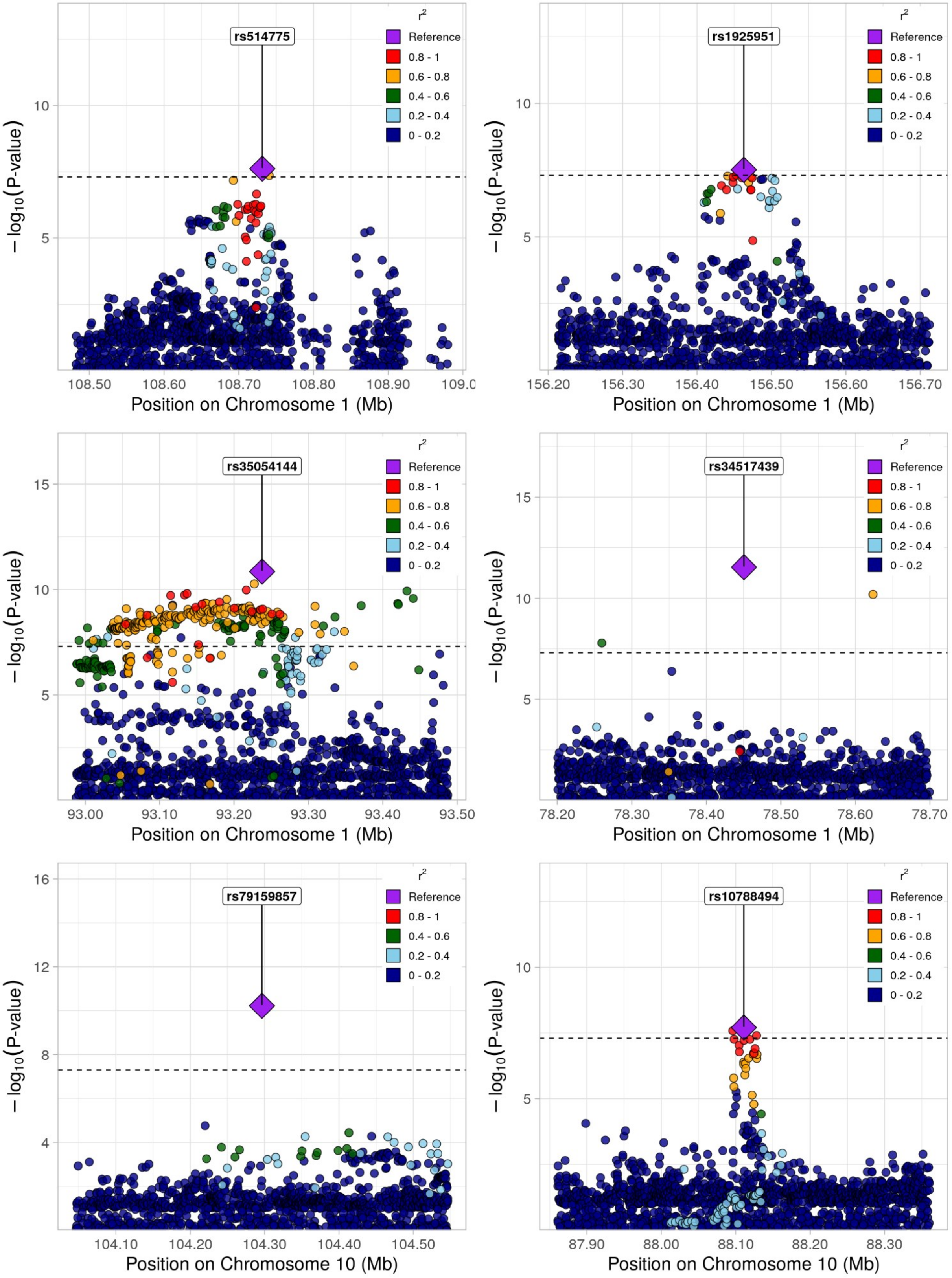
rs514775
Position on Chromosome 1 (Mb)
rs1925951
Position on Chromosome 1 (Mb)
rs35054144
Position on Chromosome 1 (Mb)
rs34517439
Position on Chromosome 1 (Mb)
rs79159857
Position on Chromosome 10 (Mb)
rs10788494
Position on Chromosome 10 (Mb)
$-\log_{10}$(P-value)
$r^2$
Reference
0.8 - 1
0.6 - 0.8
0.4 - 0.6
0.2 - 0.4
0 - 0.2

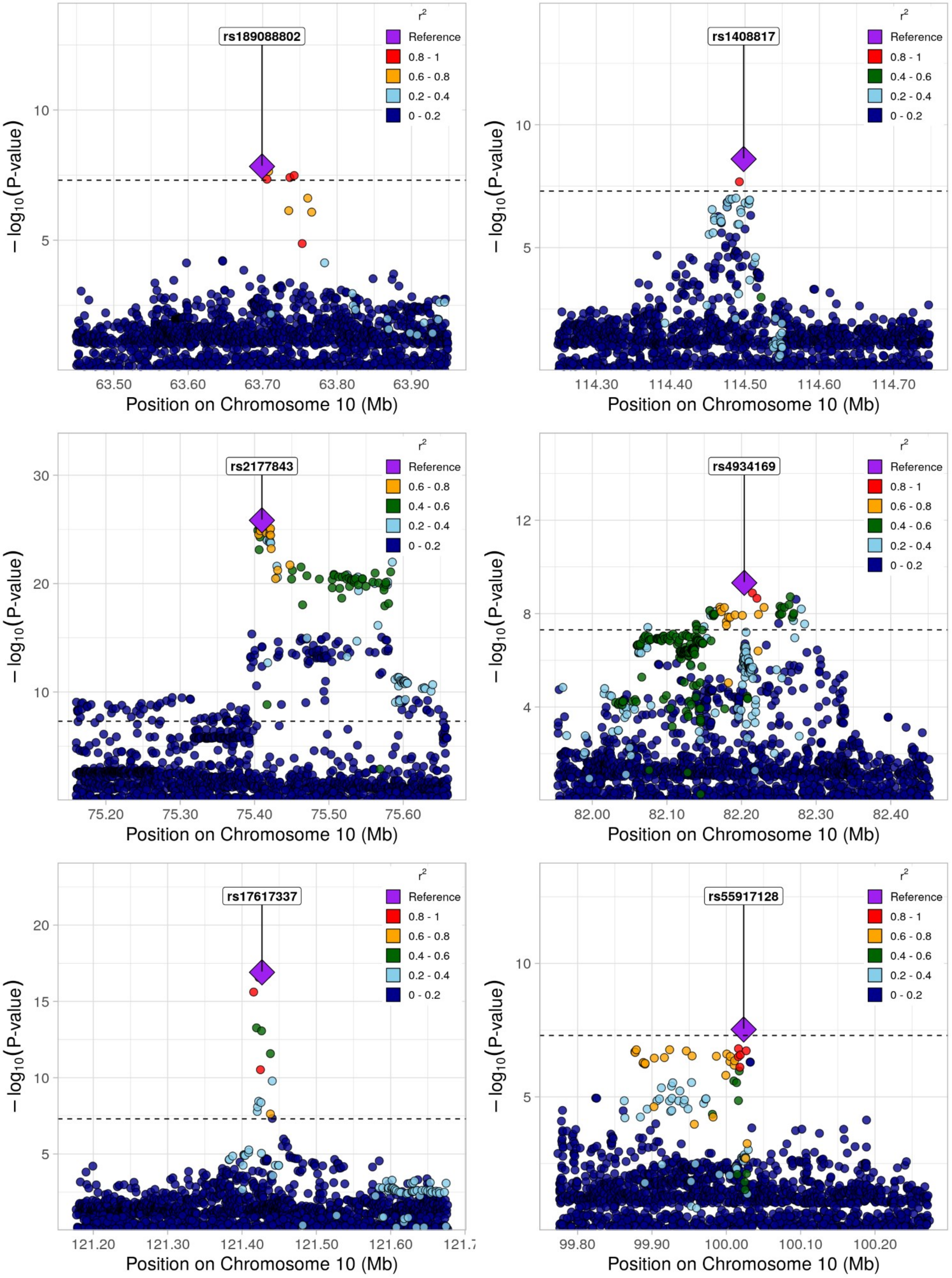

rs189088802
rs1408817
rs2177843
rs4934169
rs17617337
rs55917128
r²
Reference
0.8 - 1
0.6 - 0.8
0.4 - 0.6
0.2 - 0.4
0 - 0.2
− log10(P-value)
Position on Chromosome 10 (Mb)
63.50
63.60
63.70
63.80
63.90
114.30
114.40
114.50
114.60
114.70
75.20
75.30
75.40
75.50
75.60
82.00
82.10
82.20
82.30
82.40
121.20
121.30
121.40
121.50
121.60
99.80
99.90
100.00
100.10
100.20

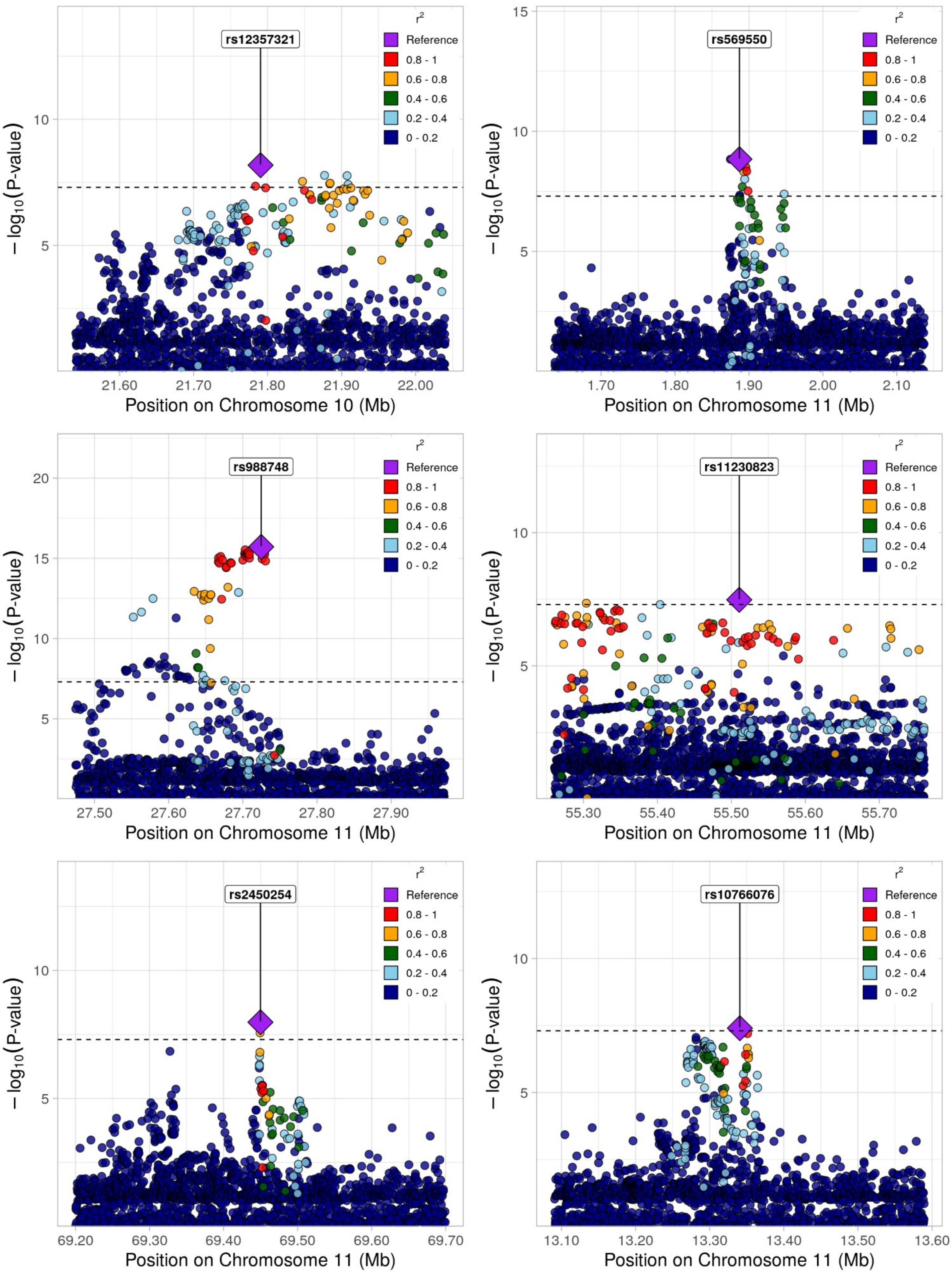
rs12357321
rs569550
rs988748
rs11230823
rs2450254
rs10766076
r²
Reference
0.8 - 1
0.6 - 0.8
0.4 - 0.6
0.2 - 0.4
0 - 0.2
− log₁₀(P-value)
Position on Chromosome 10 (Mb)
Position on Chromosome 11 (Mb)

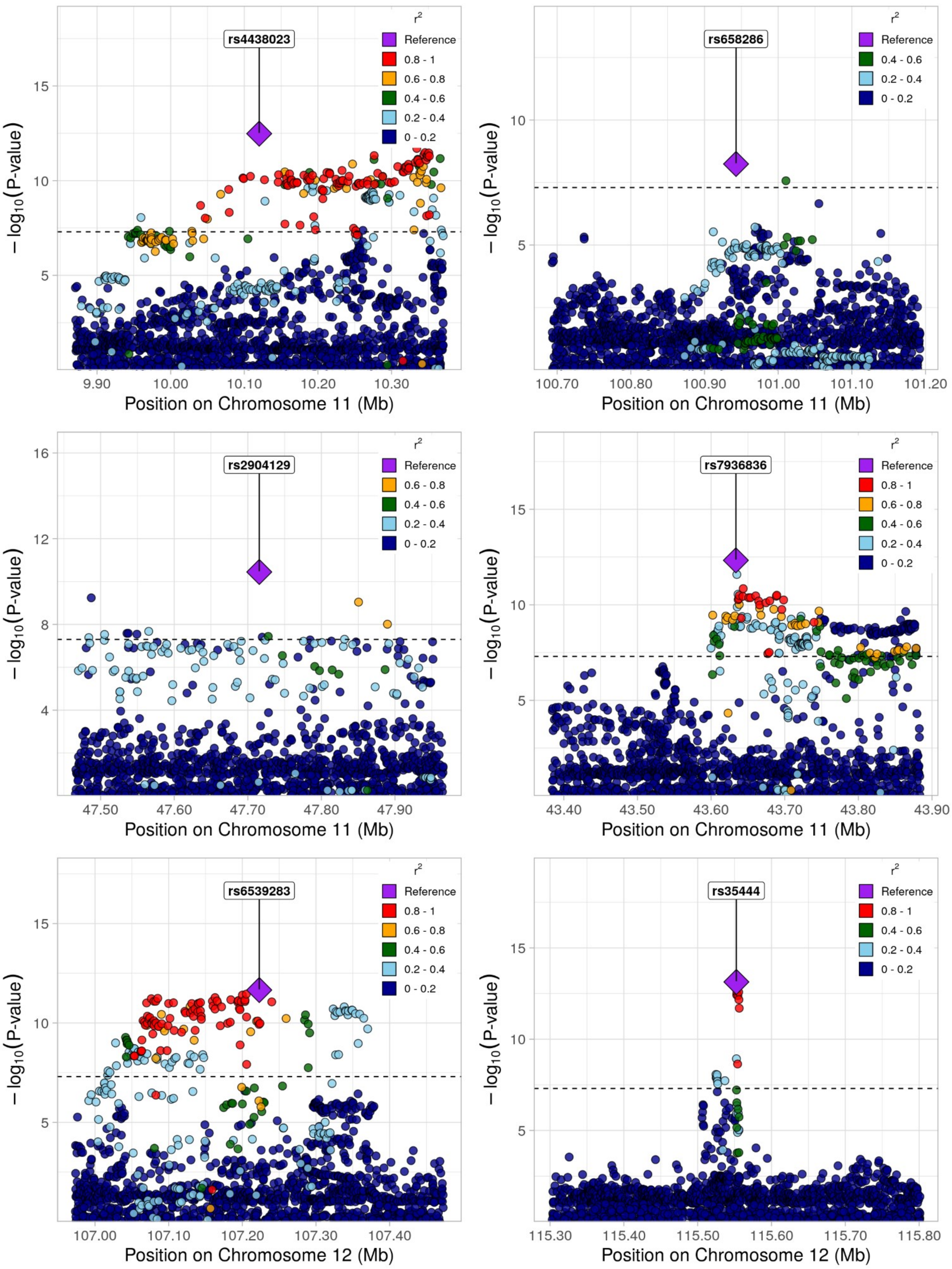
rs4438023
r²
Reference
0.8 - 1
0.6 - 0.8
0.4 - 0.6
0.2 - 0.4
0 - 0.2
−log10(P-value)
15
10
5
9.90
10.00
10.10
10.20
10.30
Position on Chromosome 11 (Mb)
rs658286
r²
Reference
0.4 - 0.6
0.2 - 0.4
0 - 0.2
10
5
100.70
100.80
100.90
101.00
101.10
101.20
Position on Chromosome 11 (Mb)
rs2904129
r²
Reference
0.6 - 0.8
0.4 - 0.6
0.2 - 0.4
0 - 0.2
16
12
8
4
47.50
47.60
47.70
47.80
47.90
Position on Chromosome 11 (Mb)
rs7936836
r²
Reference
0.8 - 1
0.6 - 0.8
0.4 - 0.6
0.2 - 0.4
0 - 0.2
15
10
5
43.40
43.50
43.60
43.70
43.80
43.90
Position on Chromosome 11 (Mb)
rs6539283
r²
Reference
0.8 - 1
0.6 - 0.8
0.4 - 0.6
0.2 - 0.4
0 - 0.2
15
10
5
107.00
107.10
107.20
107.30
107.40
Position on Chromosome 12 (Mb)
rs35444
r²
Reference
0.8 - 1
0.4 - 0.6
0.2 - 0.4
0 - 0.2
15
10
5
115.30
115.40
115.50
115.60
115.70
115.80
Position on Chromosome 12 (Mb)

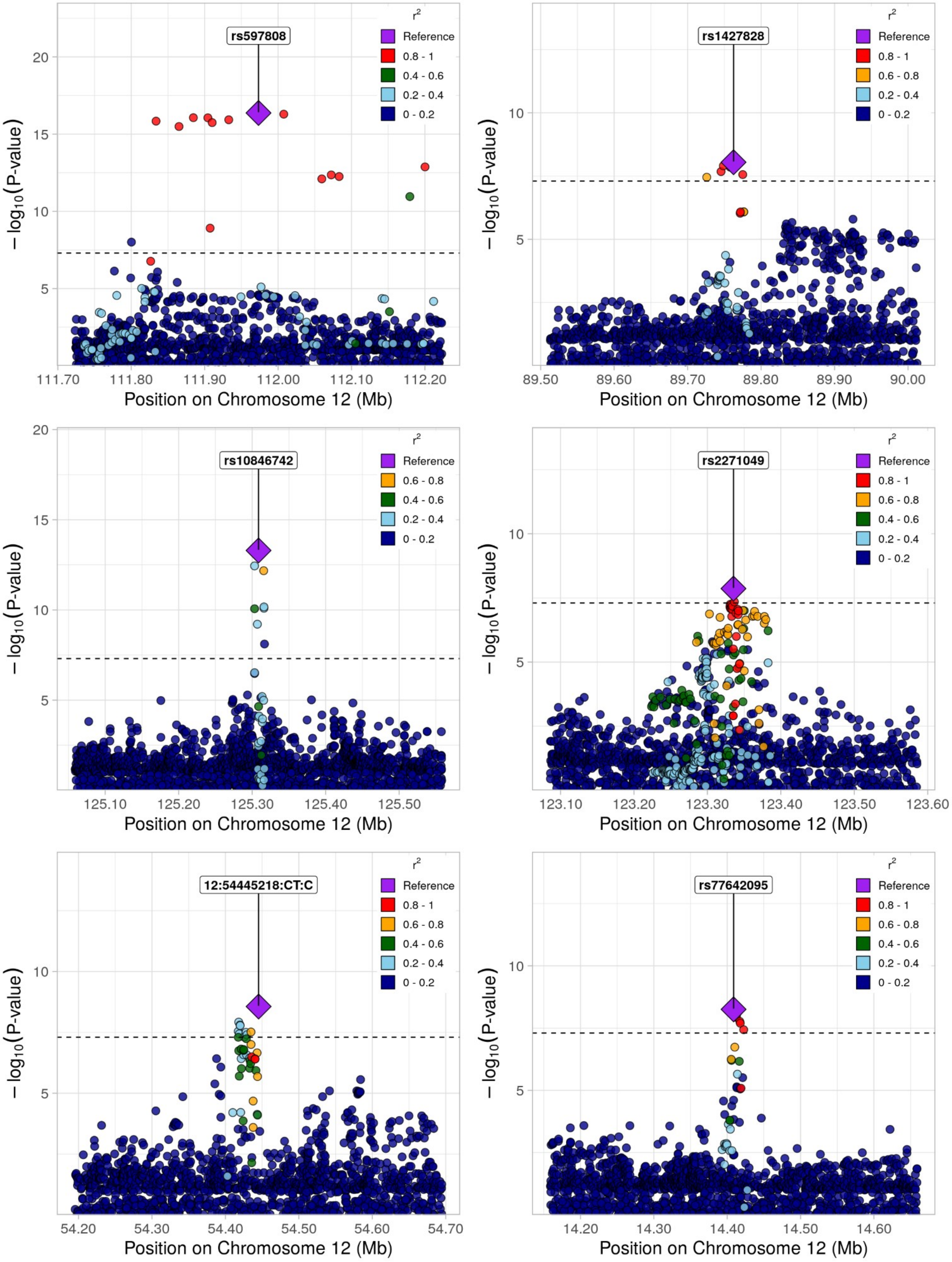
rs597808
r²
Reference
0.8 - 1
0.4 - 0.6
0.2 - 0.4
0 - 0.2
−log10(P-value)
Position on Chromosome 12 (Mb)
rs1427828
Reference
0.8 - 1
0.6 - 0.8
0.2 - 0.4
0 - 0.2
rs10846742
Reference
0.6 - 0.8
0.4 - 0.6
0.2 - 0.4
0 - 0.2
rs2271049
Reference
0.8 - 1
0.6 - 0.8
0.4 - 0.6
0.2 - 0.4
0 - 0.2
12:54445218:CT:C
rs77642095

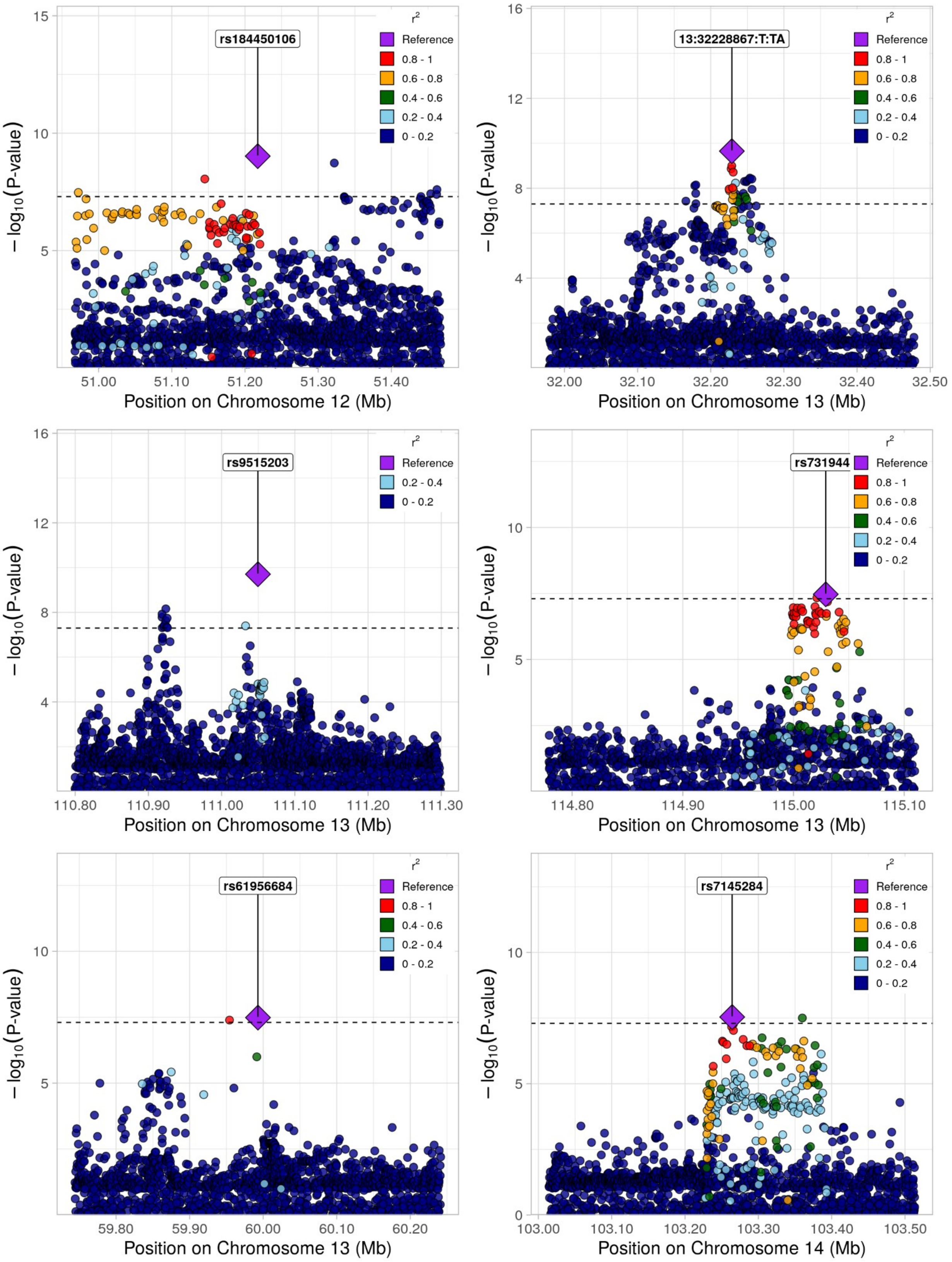

rs184450106
13:32228867:T:TA
rs9515203
rs731944
rs61956684
rs7145284
r²
Reference
0.8 - 1
0.6 - 0.8
0.4 - 0.6
0.2 - 0.4
0 - 0.2
−log10(P-value)
Position on Chromosome 12 (Mb)
Position on Chromosome 13 (Mb)
Position on Chromosome 14 (Mb)

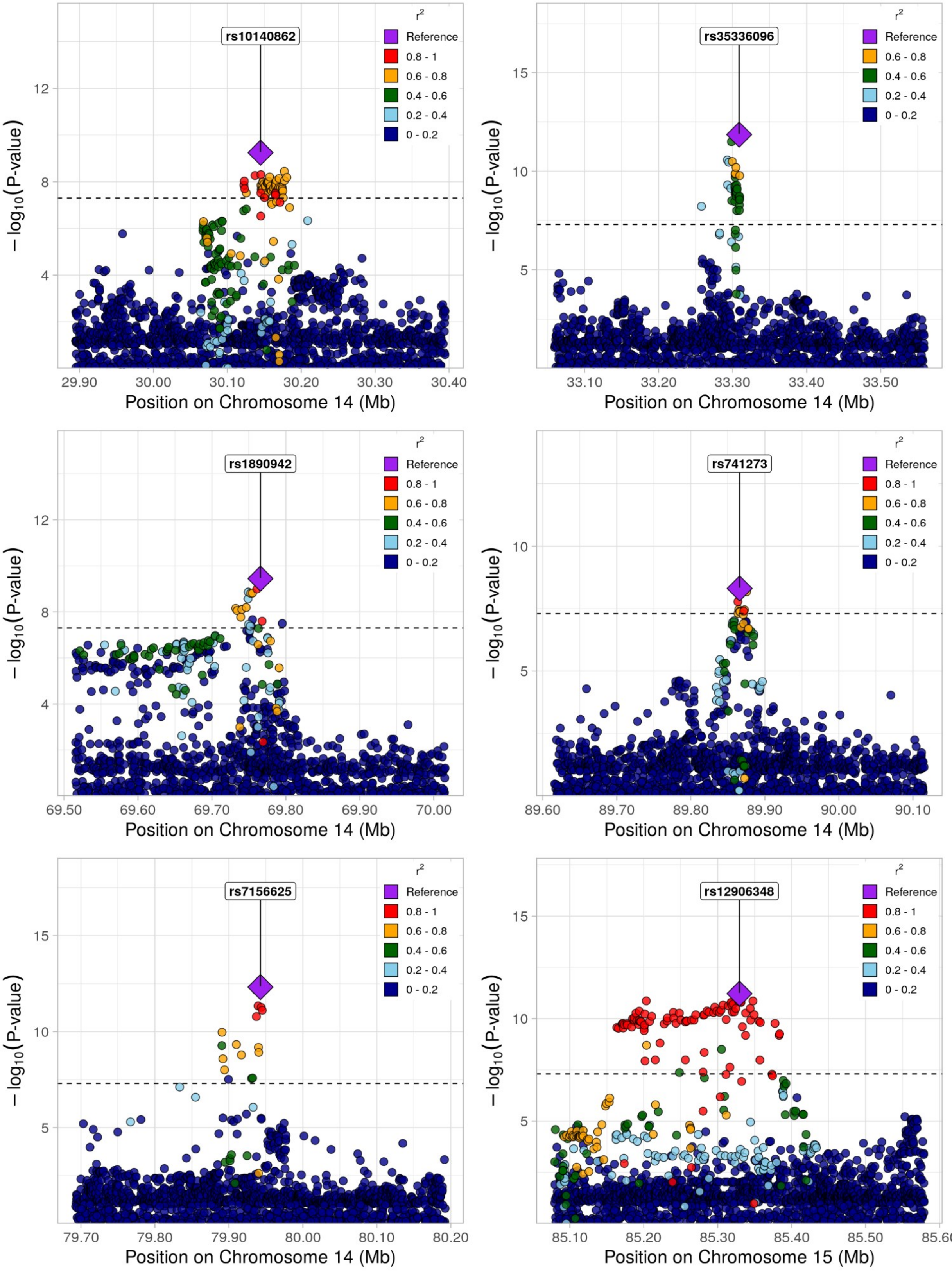
rs10140862
Position on Chromosome 14 (Mb)
$-\log_{10}$(P-value)
r²
Reference
0.8 - 1
0.6 - 0.8
0.4 - 0.6
0.2 - 0.4
0 - 0.2
rs35336096
rs1890942
rs741273
rs7156625
rs12906348
Position on Chromosome 15 (Mb)

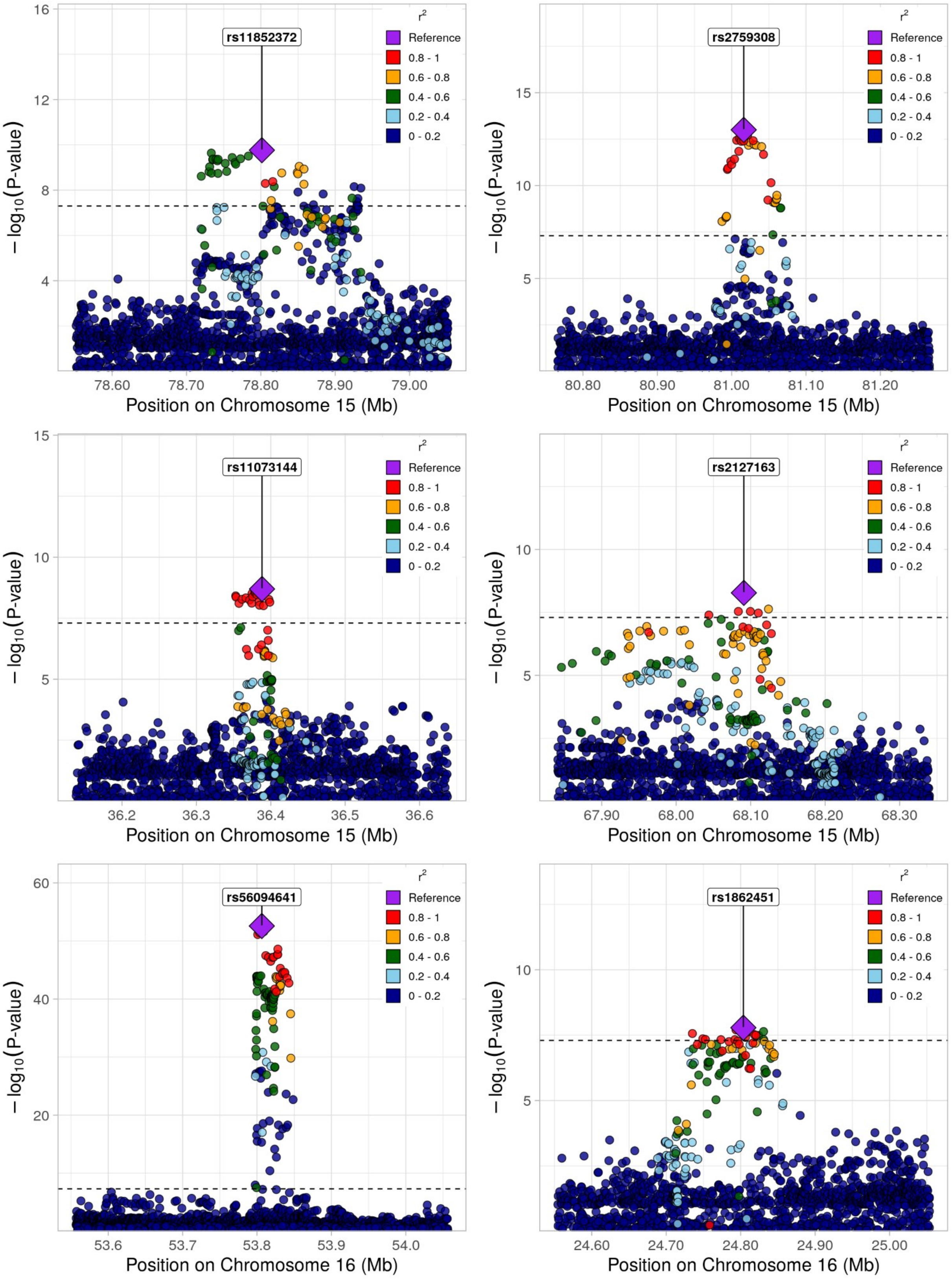
rs11852372
rs2759308
rs11073144
rs2127163
rs56094641
rs1862451
r2
Reference
0.8 - 1
0.6 - 0.8
0.4 - 0.6
0.2 - 0.4
0 - 0.2
−log10(P-value)
Position on Chromosome 15 (Mb)
Position on Chromosome 16 (Mb)

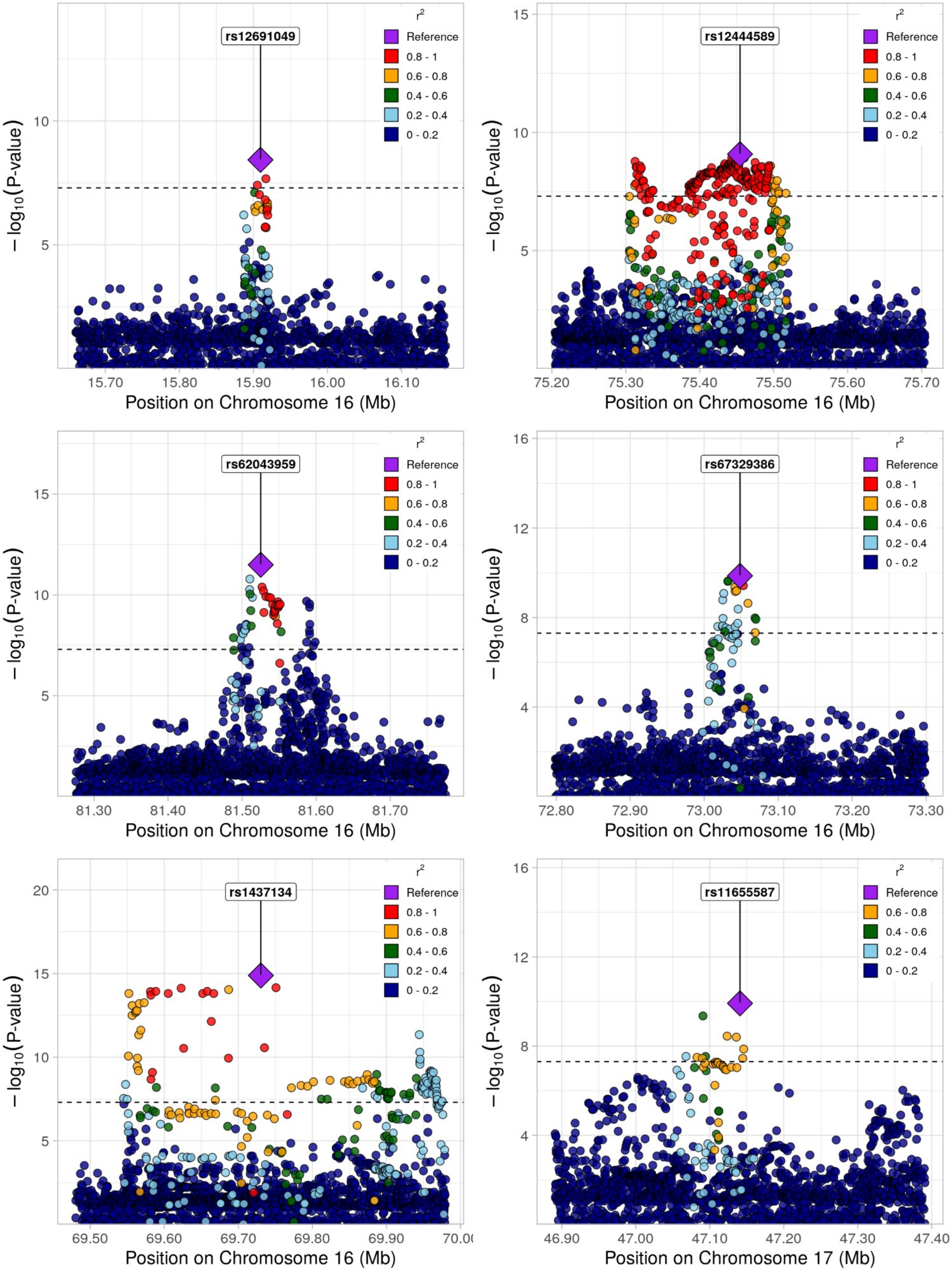
rs12691049
r²
Reference
0.8 - 1
0.6 - 0.8
0.4 - 0.6
0.2 - 0.4
0 - 0.2
− log10(P-value)
15.70
15.80
15.90
16.00
16.10
Position on Chromosome 16 (Mb)
rs12444589
75.20
75.30
75.40
75.50
75.60
75.70
Position on Chromosome 16 (Mb)
rs62043959
81.30
81.40
81.50
81.60
81.70
Position on Chromosome 16 (Mb)
rs67329386
72.80
72.90
73.00
73.10
73.20
73.30
Position on Chromosome 16 (Mb)
rs1437134
69.50
69.60
69.70
69.80
69.90
Position on Chromosome 16 (Mb)
rs11655587
46.90
47.00
47.10
47.20
47.30
47.40
Position on Chromosome 17 (Mb)

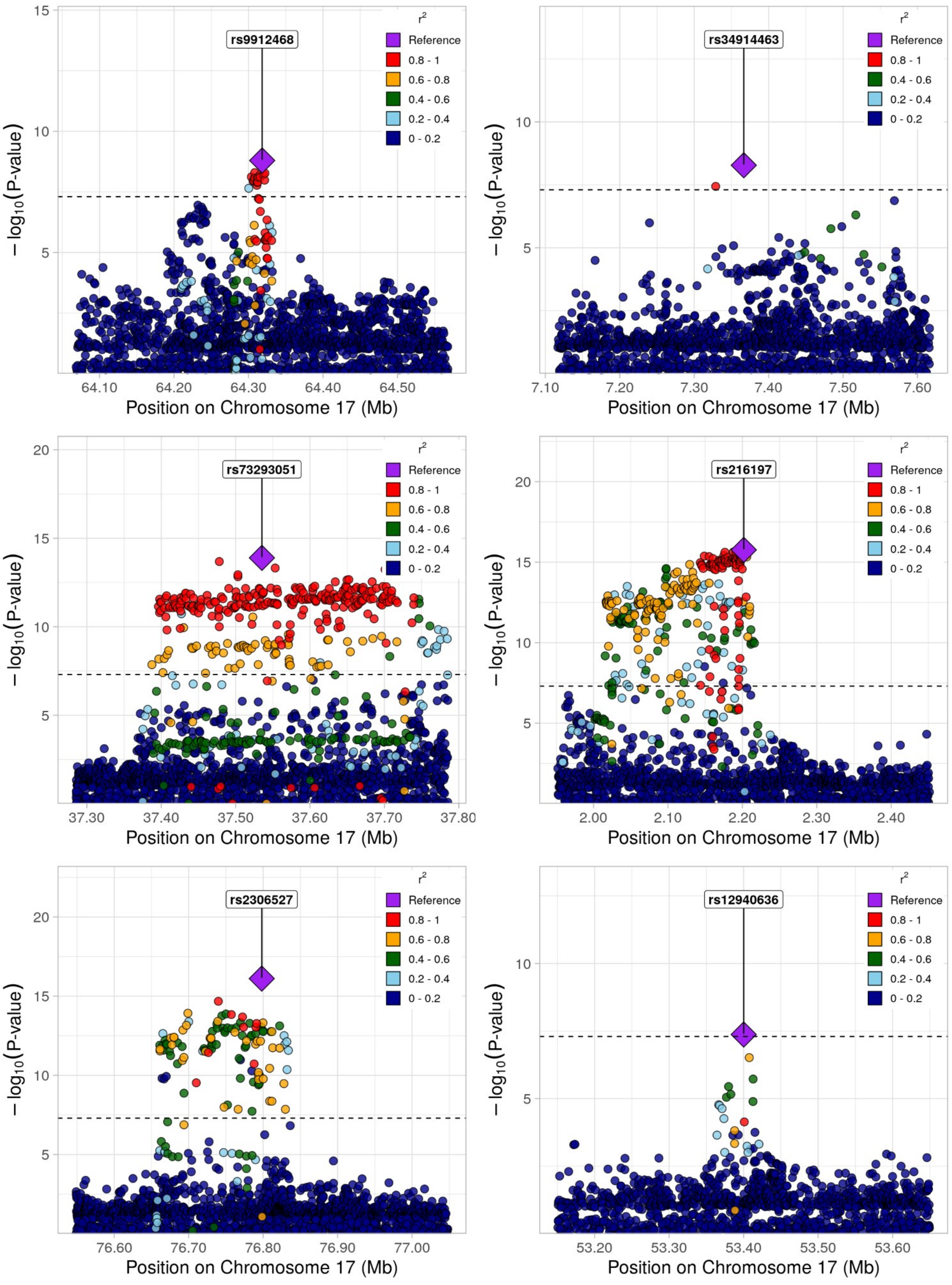
rs9912468
rs34914463
rs73293051
rs216197
rs2306527
rs12940636
r²
Reference
0.8 - 1
0.6 - 0.8
0.4 - 0.6
0.2 - 0.4
0 - 0.2
−log10(P-value)
Position on Chromosome 17 (Mb)
64.10
64.20
64.30
64.40
64.50
7.10
7.20
7.30
7.40
7.50
7.60
37.30
37.40
37.50
37.60
37.70
37.80
2.00
2.10
2.20
2.30
2.40
76.60
76.70
76.80
76.90
77.00
53.20
53.30
53.40
53.50
53.60

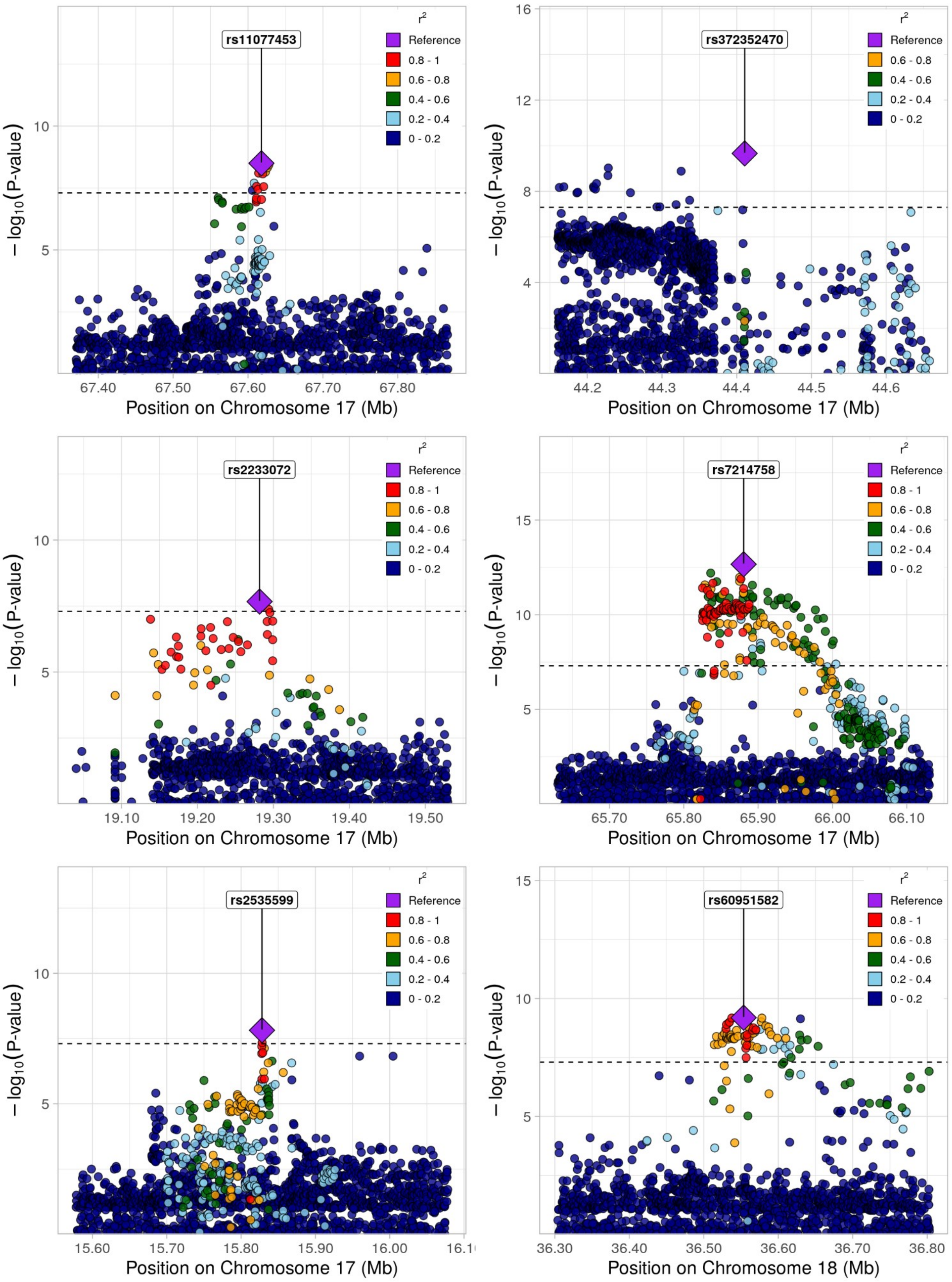
rs11077453
rs372352470
rs2233072
rs7214758
rs2535599
rs60951582
$r^2$
Reference
0.8 - 1
0.6 - 0.8
0.4 - 0.6
0.2 - 0.4
0 - 0.2
$-\log_{10}$(P-value)
Position on Chromosome 17 (Mb)
Position on Chromosome 18 (Mb)

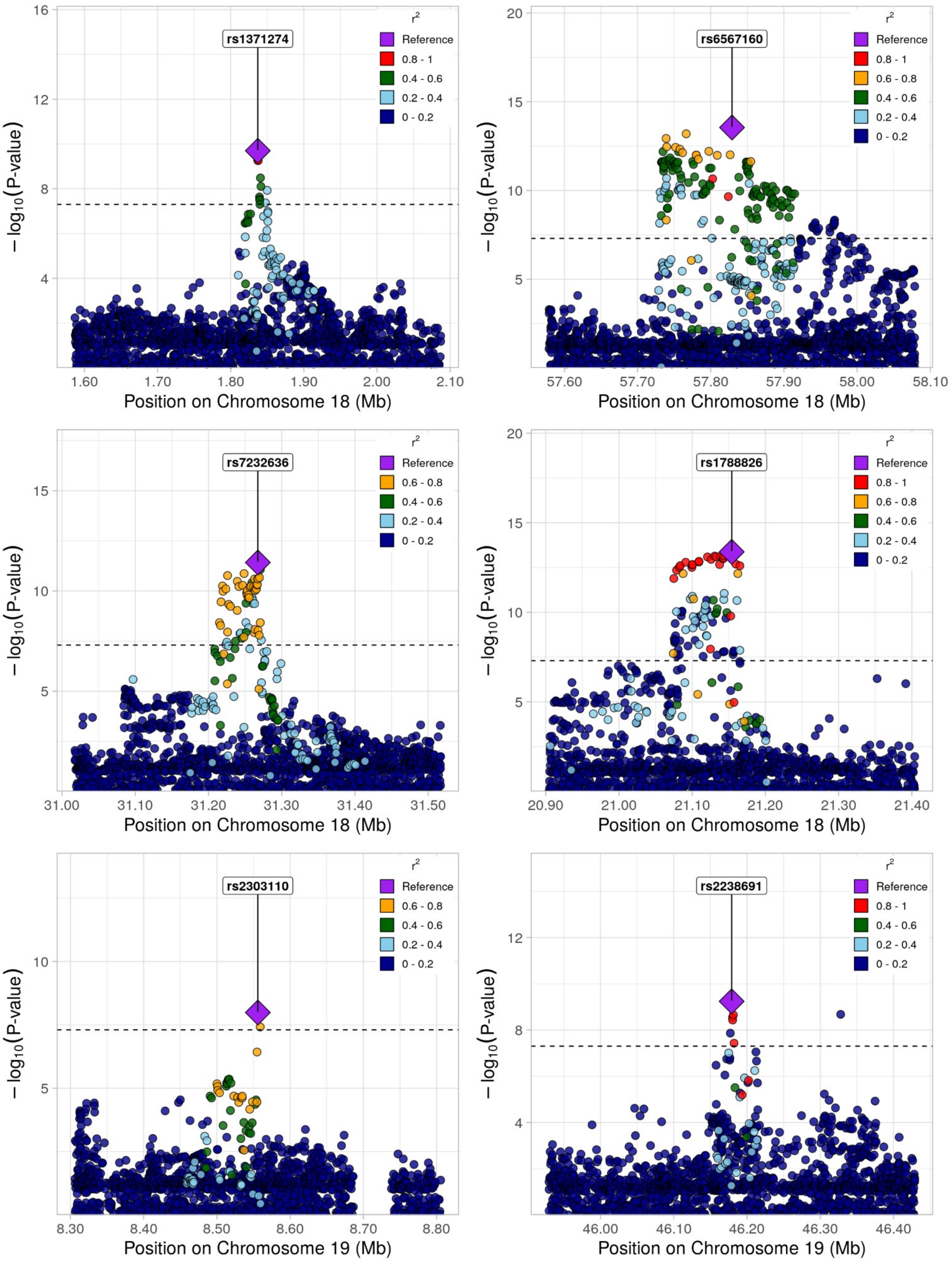
rs1371274
r²
Reference
0.8 - 1
0.4 - 0.6
0.2 - 0.4
0 - 0.2
−log10(P-value)
16
12
8
4
1.60
1.70
1.80
1.90
2.00
2.10
Position on Chromosome 18 (Mb)
rs6567160
r²
Reference
0.8 - 1
0.6 - 0.8
0.4 - 0.6
0.2 - 0.4
0 - 0.2
−log10(P-value)
20
15
10
5
57.60
57.70
57.80
57.90
58.00
58.10
Position on Chromosome 18 (Mb)
rs7232636
r²
Reference
0.6 - 0.8
0.4 - 0.6
0.2 - 0.4
0 - 0.2
−log10(P-value)
15
10
5
31.00
31.10
31.20
31.30
31.40
31.50
Position on Chromosome 18 (Mb)
rs1788826
r²
Reference
0.8 - 1
0.6 - 0.8
0.4 - 0.6
0.2 - 0.4
0 - 0.2
−log10(P-value)
20
15
10
5
20.90
21.00
21.10
21.20
21.30
21.40
Position on Chromosome 18 (Mb)
rs2303110
r²
Reference
0.6 - 0.8
0.4 - 0.6
0.2 - 0.4
0 - 0.2
−log10(P-value)
10
5
8.30
8.40
8.50
8.60
8.70
8.80
Position on Chromosome 19 (Mb)
rs2238691
r²
Reference
0.8 - 1
0.4 - 0.6
0.2 - 0.4
0 - 0.2
−log10(P-value)
12
8
4
46.00
46.10
46.20
46.30
46.40
Position on Chromosome 19 (Mb)

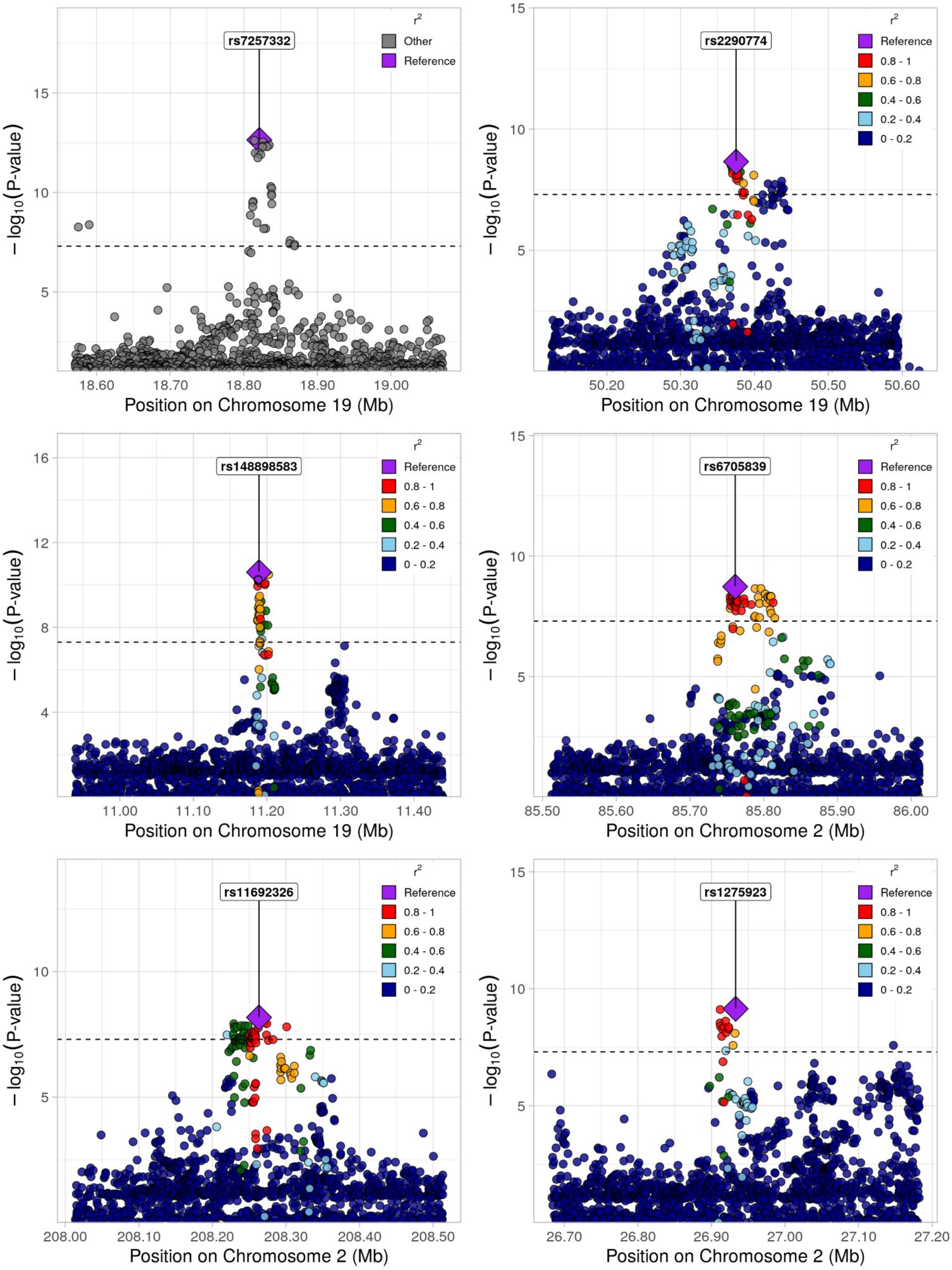
rs7257332
r²
Other
Reference
−log10(P-value)
Position on Chromosome 19 (Mb)
18.60
18.70
18.80
18.90
19.00
rs2290774
Reference
0.8 - 1
0.6 - 0.8
0.4 - 0.6
0.2 - 0.4
0 - 0.2
50.20
50.30
50.40
50.50
50.60
rs148898583
11.00
11.10
11.20
11.30
11.40
rs6705839
Position on Chromosome 2 (Mb)
85.50
85.60
85.70
85.80
85.90
86.00
rs11692326
208.00
208.10
208.20
208.30
208.40
208.50
rs1275923
26.70
26.80
26.90
27.00
27.10
27.20

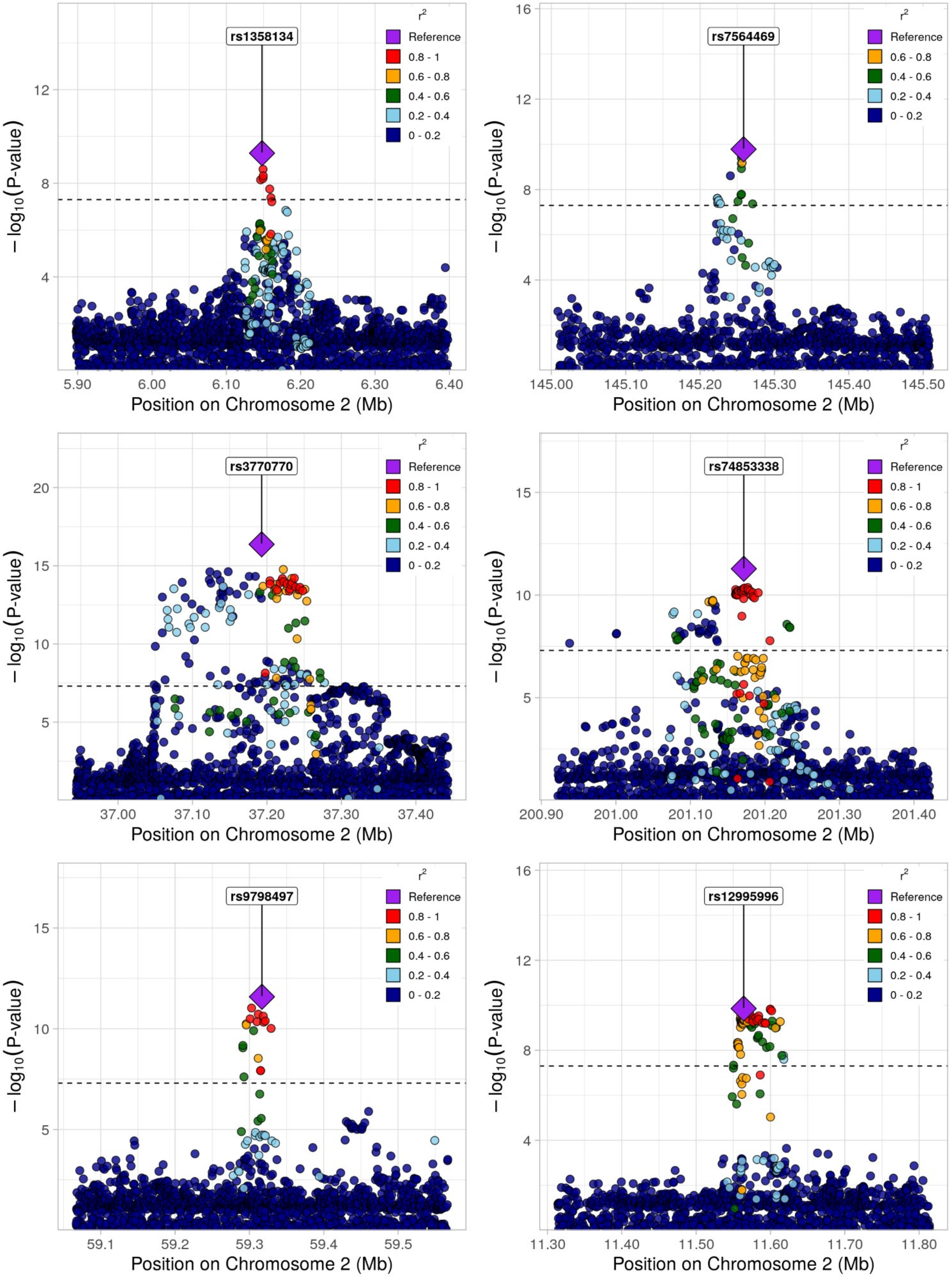
rs1358134
rs7564469
rs3770770
rs74853338
rs9798497
rs12995996
r²
Reference
0.8 - 1
0.6 - 0.8
0.4 - 0.6
0.2 - 0.4
0 - 0.2
−log10(P-value)
Position on Chromosome 2 (Mb)
5.90
6.00
6.10
6.20
6.30
6.40
145.00
145.10
145.20
145.30
145.40
145.50
37.00
37.10
37.20
37.30
37.40
200.90
201.00
201.10
201.20
201.30
201.40
59.1
59.2
59.3
59.4
59.5
11.30
11.40
11.50
11.60
11.70
11.80

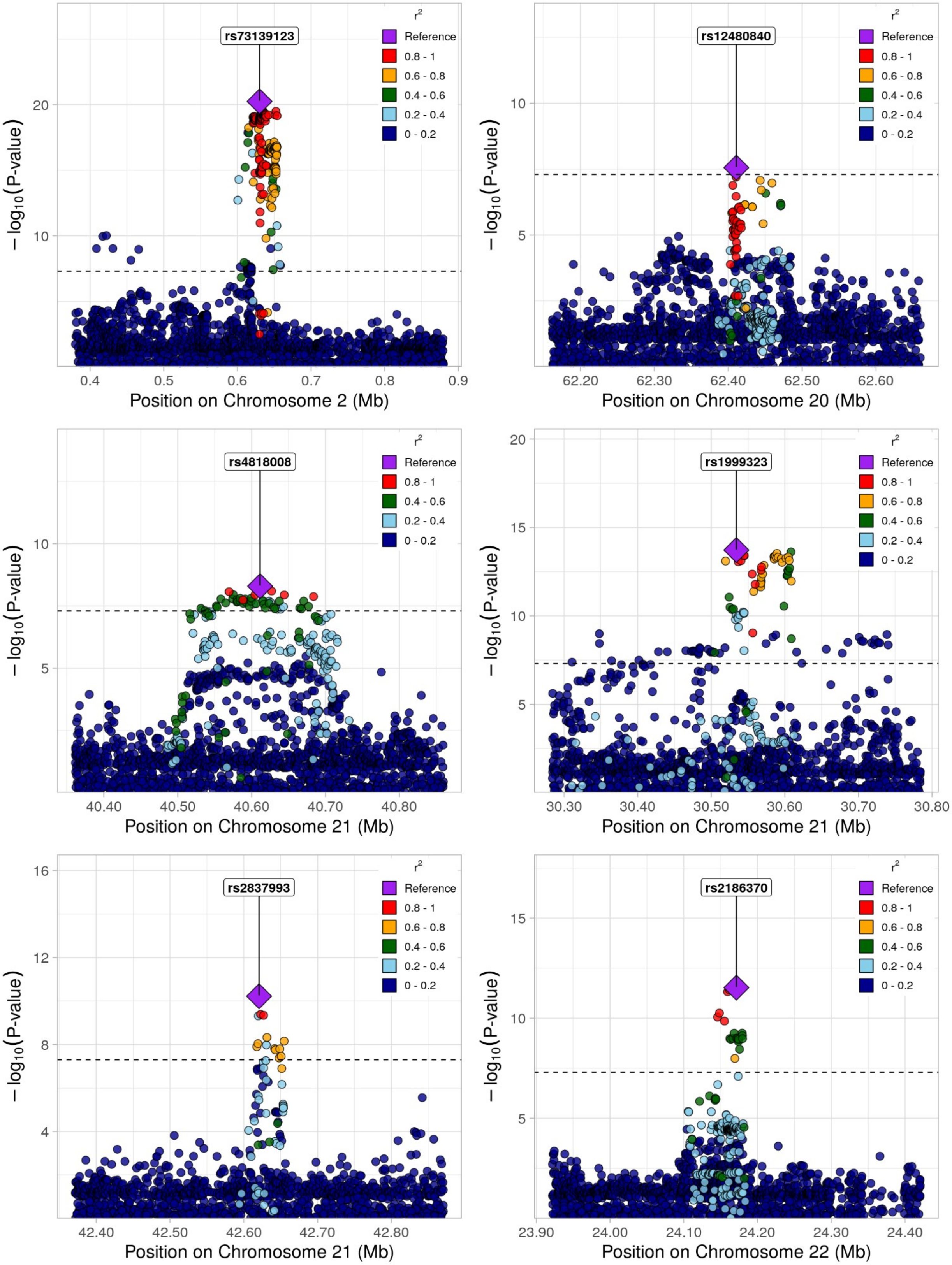

rs73139123
r²
Reference
0.8 - 1
0.6 - 0.8
0.4 - 0.6
0.2 - 0.4
0 - 0.2
−log₁₀(P-value)
Position on Chromosome 2 (Mb)
rs12480840
Position on Chromosome 20 (Mb)
rs4818008
Position on Chromosome 21 (Mb)
rs1999323
Position on Chromosome 21 (Mb)
rs2837993
Position on Chromosome 21 (Mb)
rs2186370
Position on Chromosome 22 (Mb)

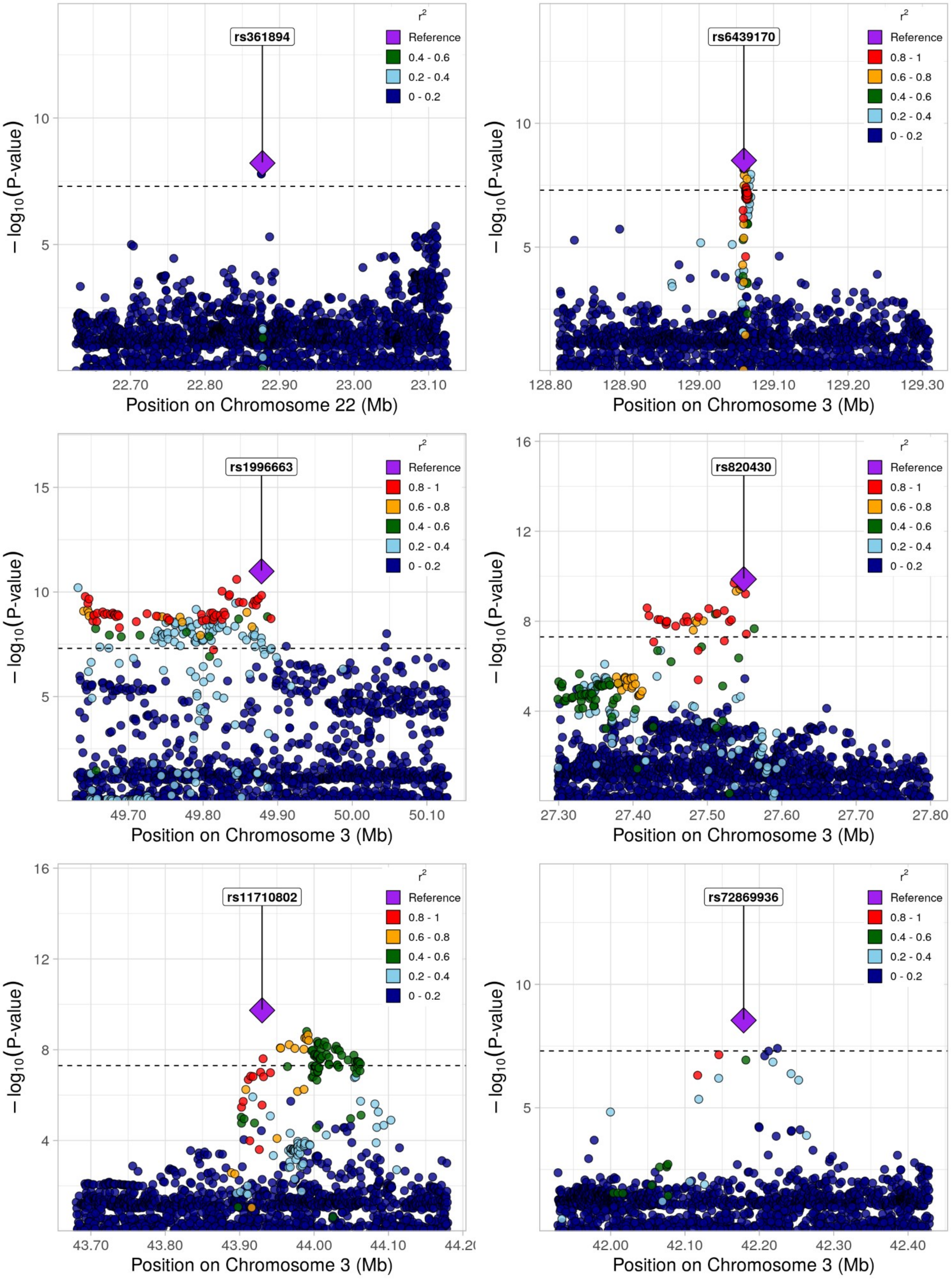
rs361894
r2
Reference
0.4 - 0.6
0.2 - 0.4
0 - 0.2
− log10(P-value)
10
5
22.70
22.80
22.90
23.00
23.10
Position on Chromosome 22 (Mb)
rs6439170
r2
Reference
0.8 - 1
0.6 - 0.8
0.4 - 0.6
0.2 - 0.4
0 - 0.2
− log10(P-value)
10
5
128.80
128.90
129.00
129.10
129.20
129.30
Position on Chromosome 3 (Mb)
rs1996663
r2
Reference
0.8 - 1
0.6 - 0.8
0.4 - 0.6
0.2 - 0.4
0 - 0.2
− log10(P-value)
15
10
5
49.70
49.80
49.90
50.00
50.10
Position on Chromosome 3 (Mb)
rs820430
r2
Reference
0.8 - 1
0.6 - 0.8
0.4 - 0.6
0.2 - 0.4
0 - 0.2
− log10(P-value)
16
12
8
4
27.30
27.40
27.50
27.60
27.70
27.80
Position on Chromosome 3 (Mb)
rs11710802
r2
Reference
0.8 - 1
0.6 - 0.8
0.4 - 0.6
0.2 - 0.4
0 - 0.2
− log10(P-value)
16
12
8
4
43.70
43.80
43.90
44.00
44.10
Position on Chromosome 3 (Mb)
rs72869936
r2
Reference
0.8 - 1
0.4 - 0.6
0.2 - 0.4
0 - 0.2
− log10(P-value)
10
5
42.00
42.10
42.20
42.30
42.40
Position on Chromosome 3 (Mb)

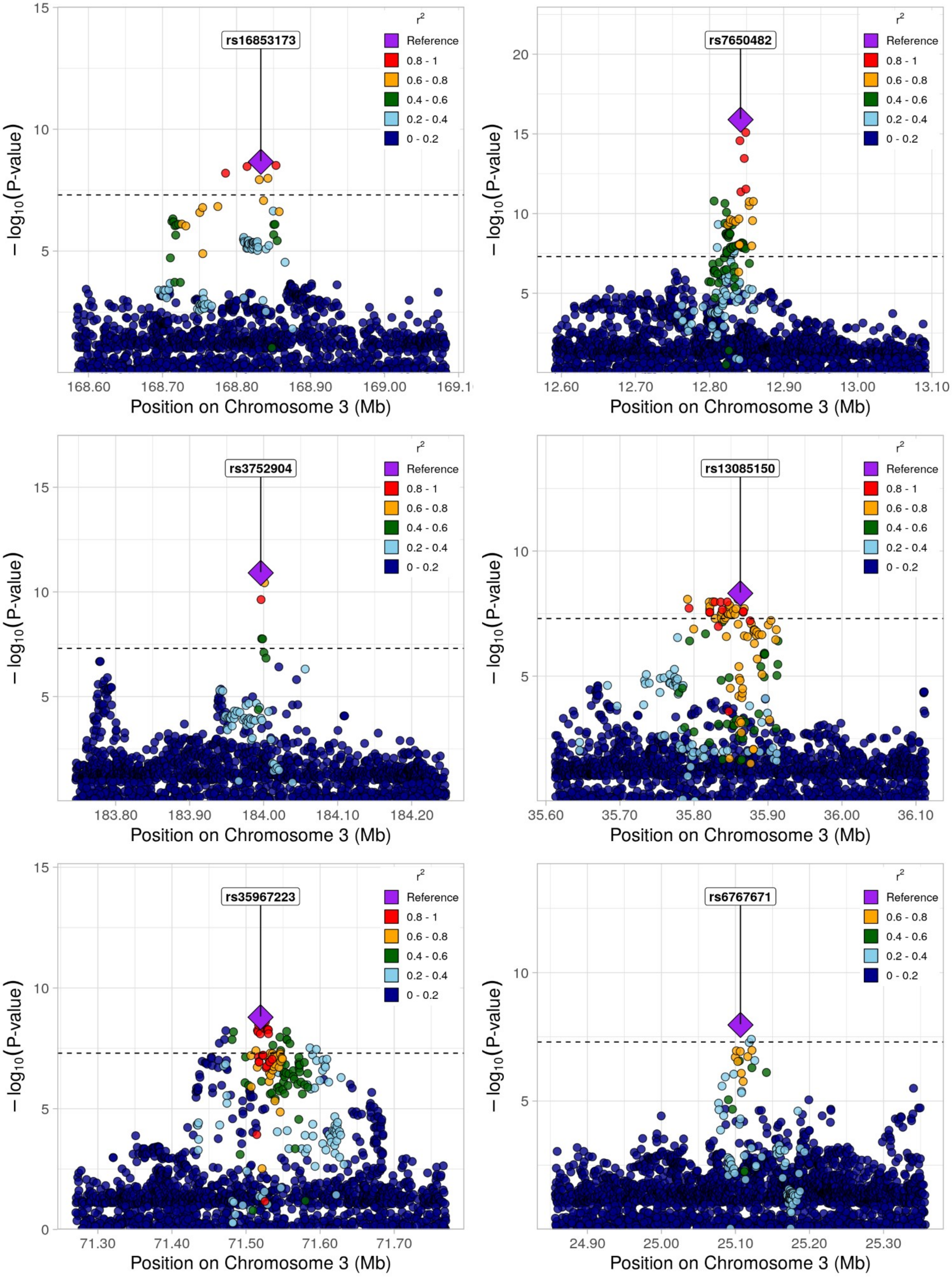
rs16853173
rs7650482
rs3752904
rs13085150
rs35967223
rs6767671
r²
Reference
0.8 - 1
0.6 - 0.8
0.4 - 0.6
0.2 - 0.4
0 - 0.2
− log10(P-value)
Position on Chromosome 3 (Mb)
168.60
168.70
168.80
168.90
169.00
12.60
12.70
12.80
12.90
13.00
13.10
183.80
183.90
184.00
184.10
184.20
35.60
35.70
35.80
35.90
36.00
36.10
71.30
71.40
71.50
71.60
71.70
24.90
25.00
25.10
25.20
25.30

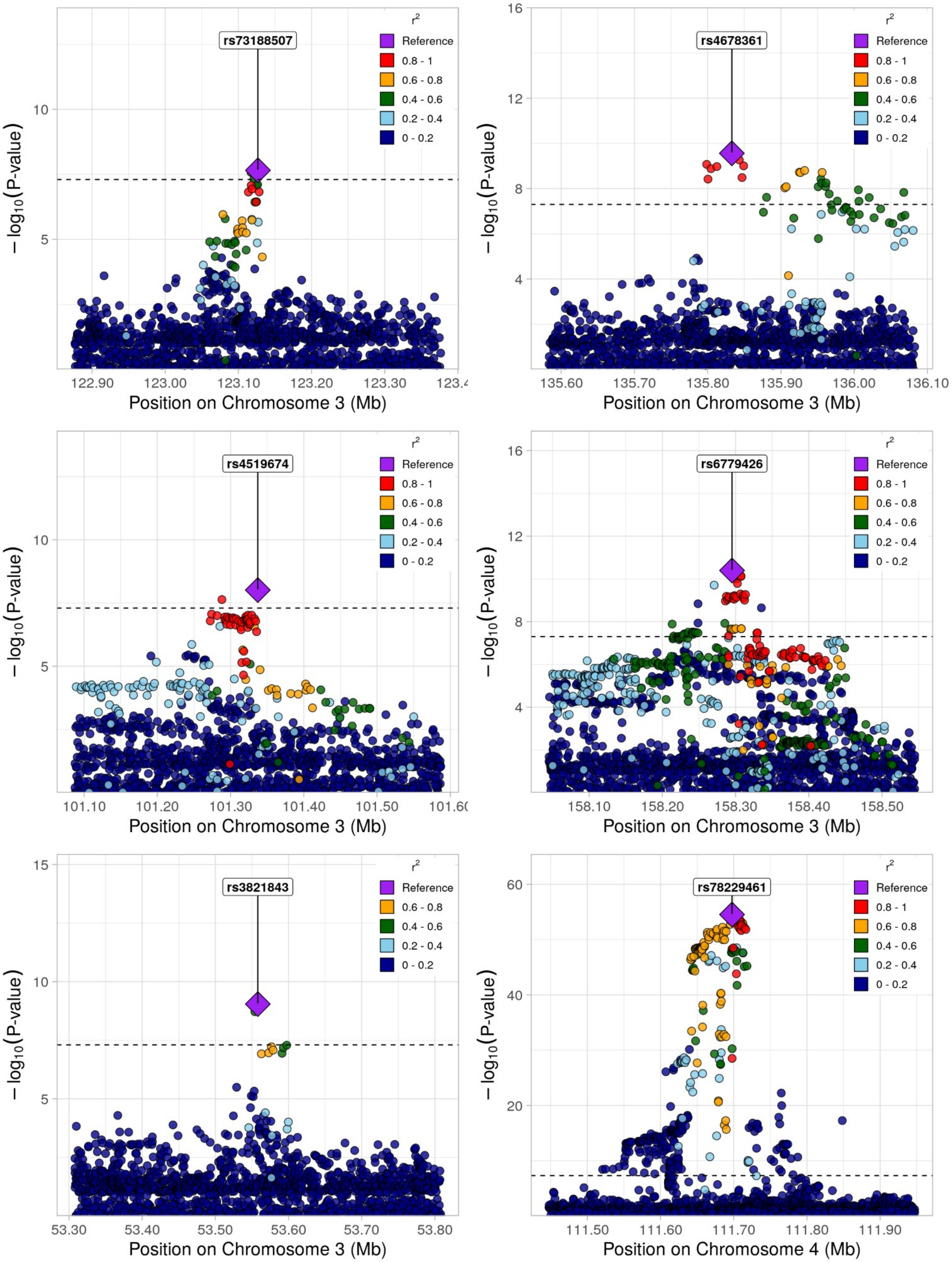
rs73188507
rs4678361
rs4519674
rs6779426
rs3821843
rs78229461
r²
Reference
0.8 - 1
0.6 - 0.8
0.4 - 0.6
0.2 - 0.4
0 - 0.2
−log10(P-value)
Position on Chromosome 3 (Mb)
Position on Chromosome 4 (Mb)
122.90
123.00
123.10
123.20
123.30
135.60
135.70
135.80
135.90
136.00
136.10
101.10
101.20
101.30
101.40
101.50
158.10
158.20
158.30
158.40
158.50
53.30
53.40
53.50
53.60
53.70
53.80
111.50
111.60
111.70
111.80
111.90

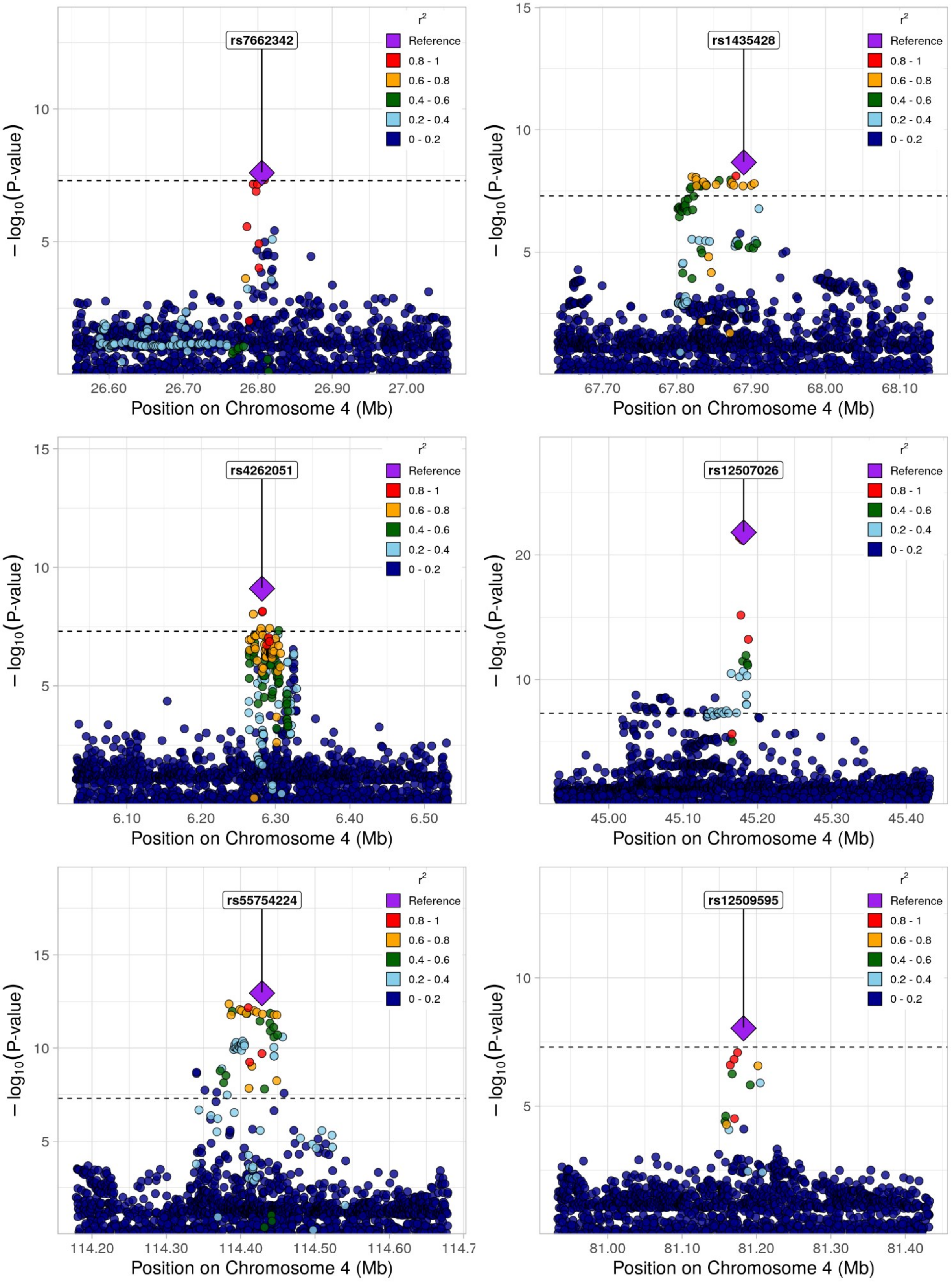

rs7662342
rs1435428
rs4262051
rs12507026
rs55754224
rs12509595
r²
Reference
0.8 - 1
0.6 - 0.8
0.4 - 0.6
0.2 - 0.4
0 - 0.2
− log10(P-value)
Position on Chromosome 4 (Mb)

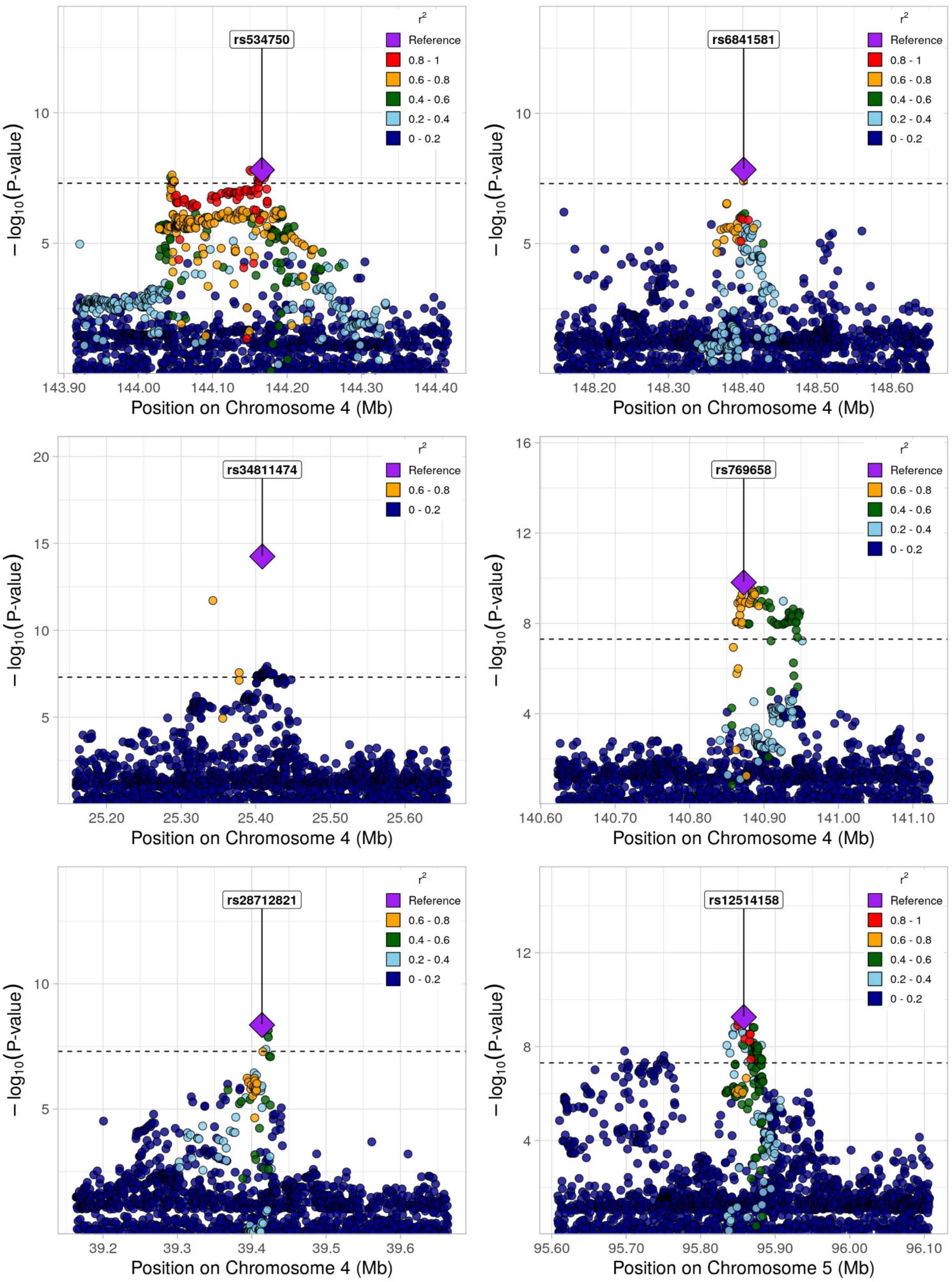
rs534750
rs6841581
rs34811474
rs769658
rs28712821
rs12514158
r²
Reference
0.8 - 1
0.6 - 0.8
0.4 - 0.6
0.2 - 0.4
0 - 0.2
− log10(P-value)
Position on Chromosome 4 (Mb)
Position on Chromosome 5 (Mb)

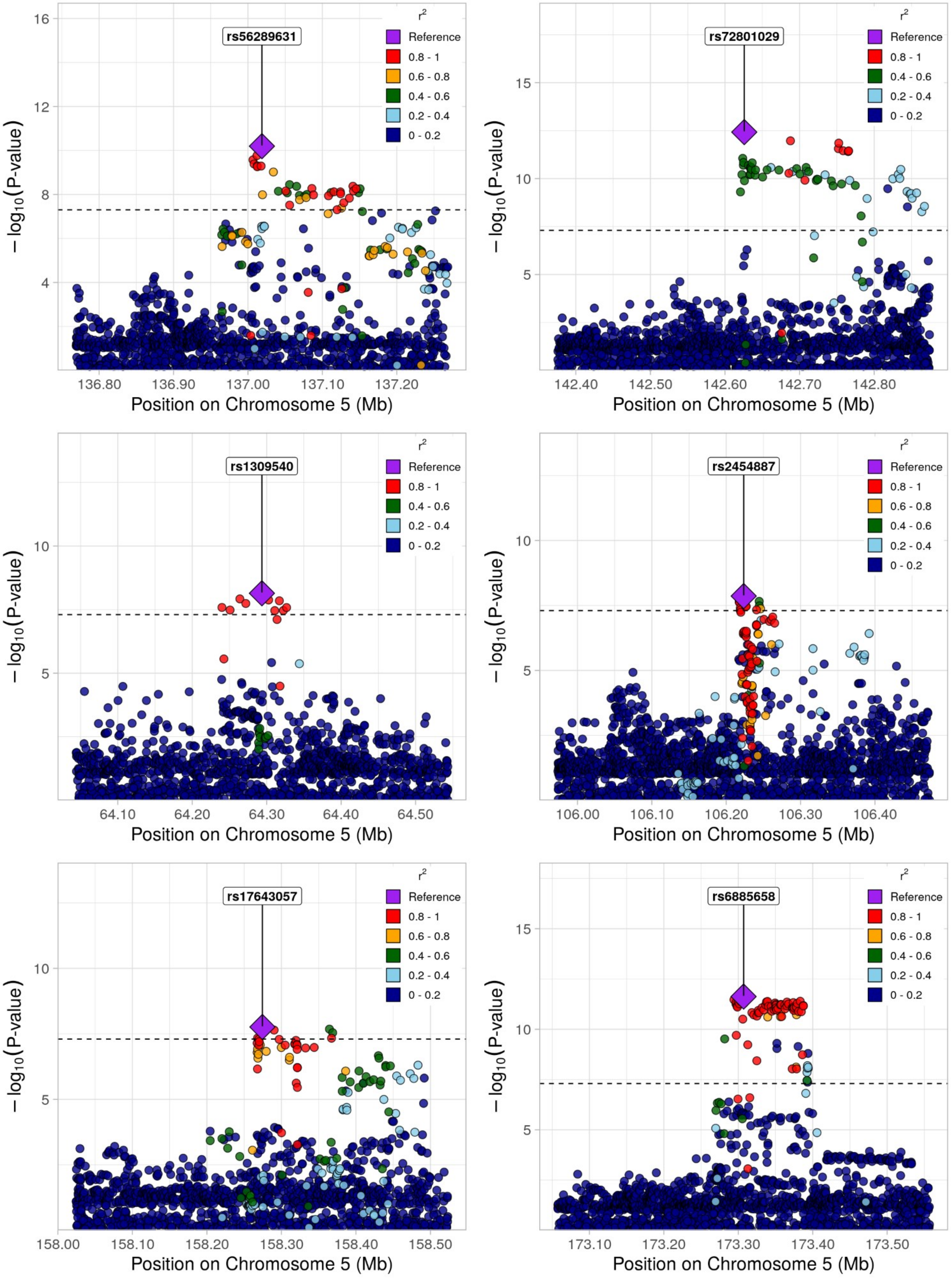
rs56289631
rs72801029
rs1309540
rs2454887
rs17643057
rs6885658
r²
Reference
0.8 - 1
0.6 - 0.8
0.4 - 0.6
0.2 - 0.4
0 - 0.2
−log₁₀(P-value)
Position on Chromosome 5 (Mb)

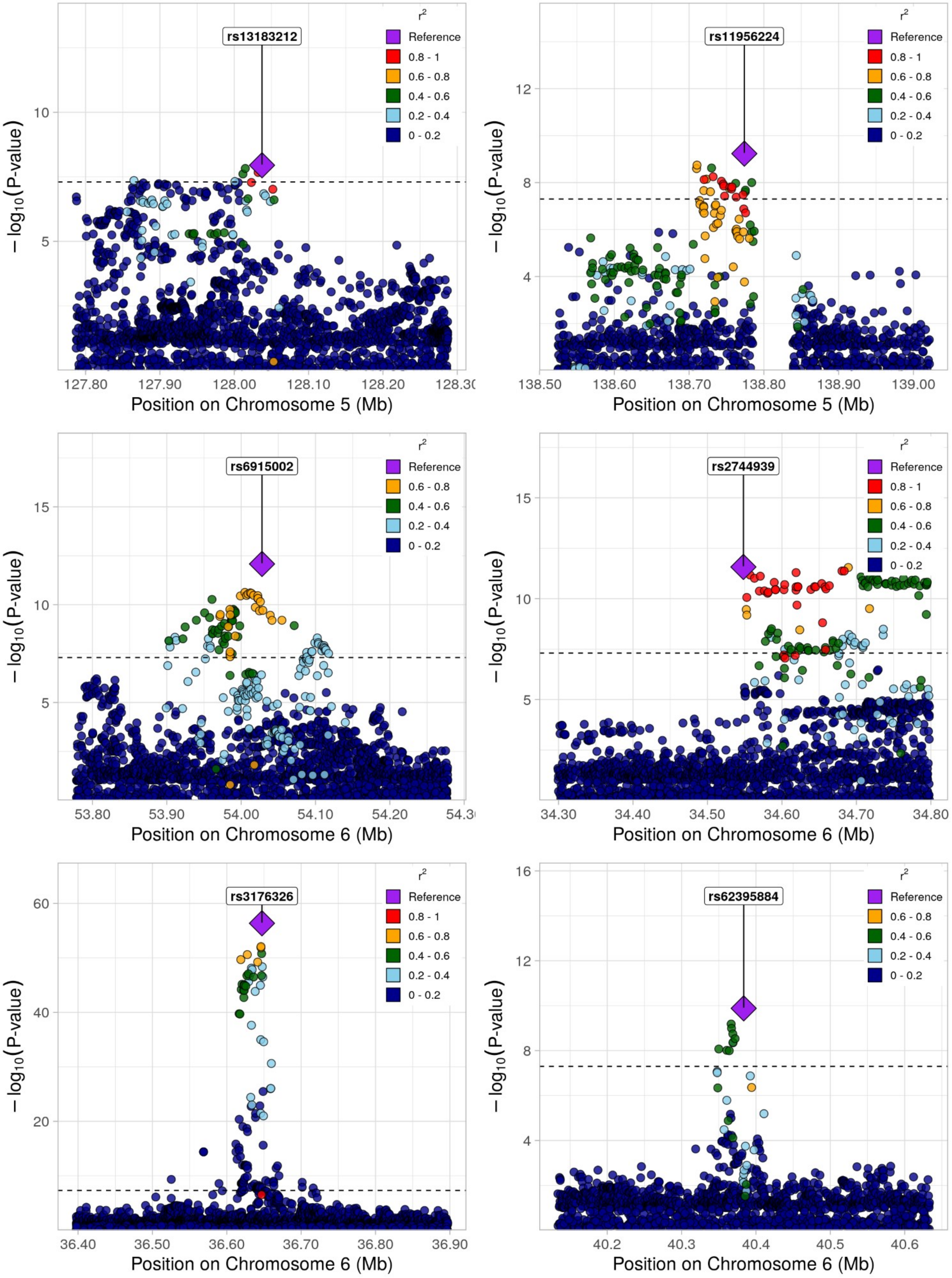
rs13183212
r²
Reference
0.8 - 1
0.6 - 0.8
0.4 - 0.6
0.2 - 0.4
0 - 0.2
−log₁₀(P-value)
Position on Chromosome 5 (Mb)
rs11956224
Position on Chromosome 5 (Mb)
rs6915002
Position on Chromosome 6 (Mb)
rs2744939
Position on Chromosome 6 (Mb)
rs3176326
Position on Chromosome 6 (Mb)
rs62395884
Position on Chromosome 6 (Mb)

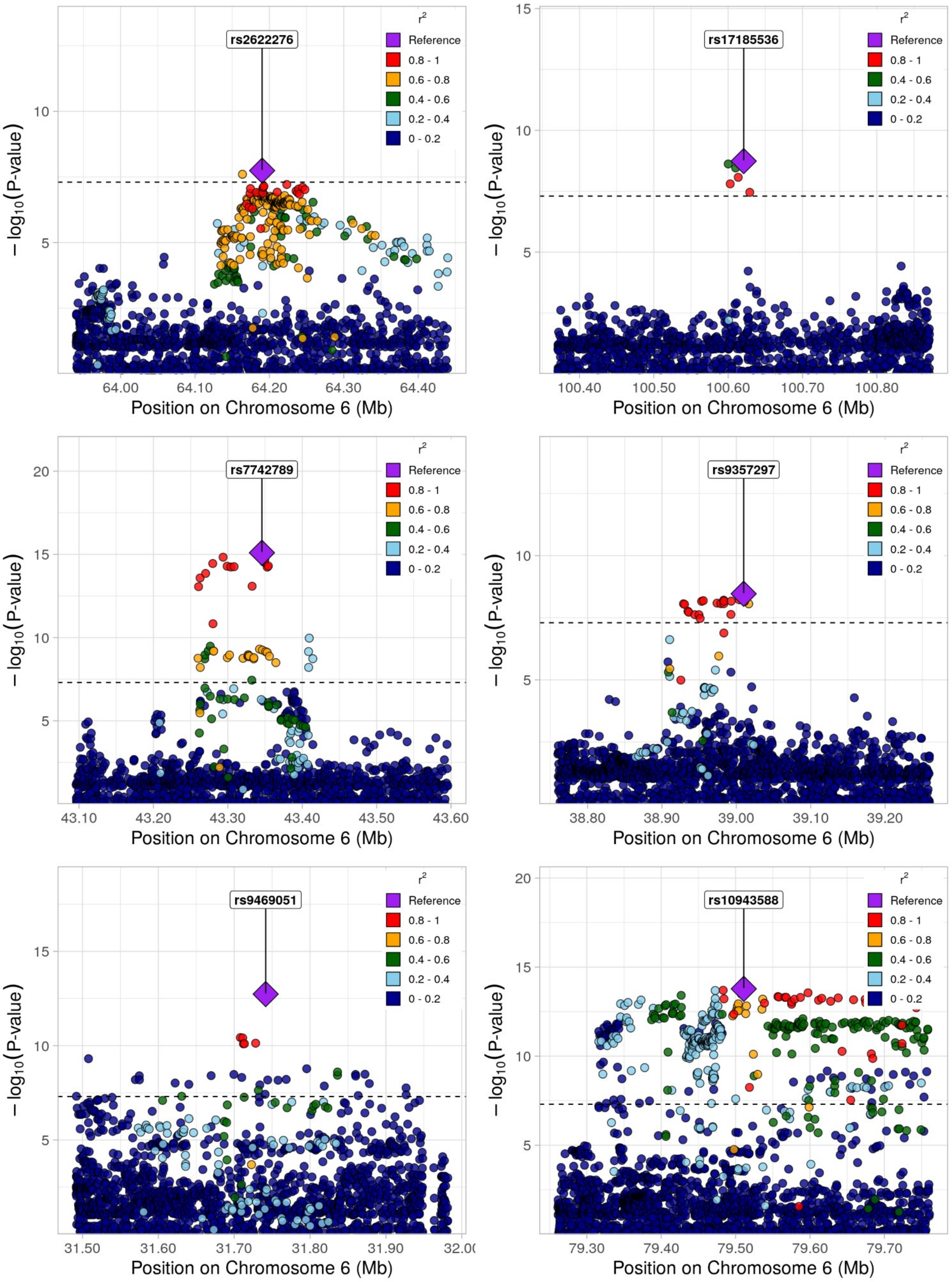
rs2622276
rs17185536
rs7742789
rs9357297
rs9469051
rs10943588
r²
Reference
0.8 - 1
0.6 - 0.8
0.4 - 0.6
0.2 - 0.4
0 - 0.2
− log10(P-value)
Position on Chromosome 6 (Mb)

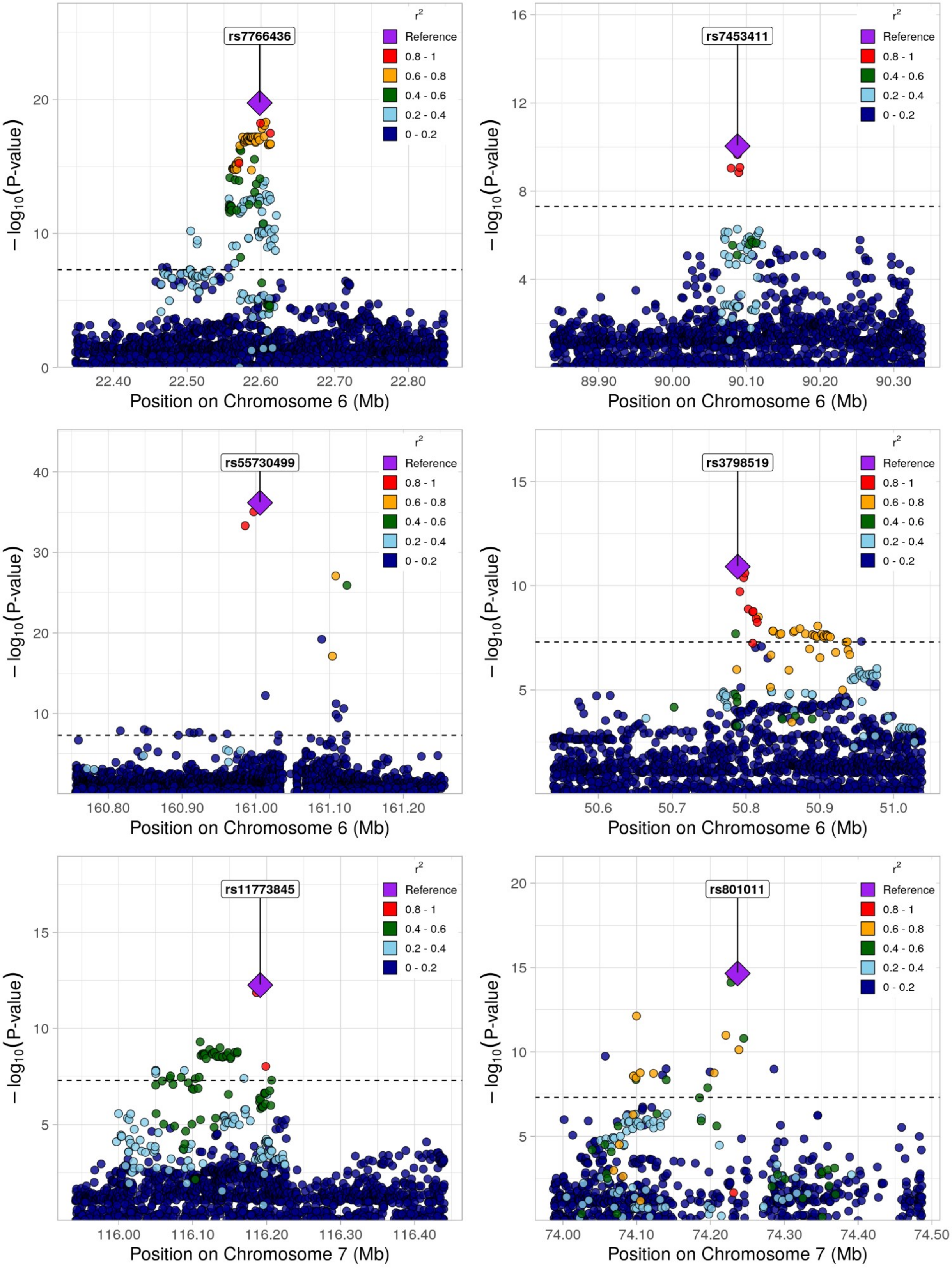
rs7766436
Reference
0.8 - 1
0.6 - 0.8
0.4 - 0.6
0.2 - 0.4
0 - 0.2
r2
−log10(P-value)
Position on Chromosome 6 (Mb)
22.40
22.50
22.60
22.70
22.80
0
10
20
rs7453411
Position on Chromosome 6 (Mb)
89.90
90.00
90.10
90.20
90.30
4
8
12
16
rs55730499
Position on Chromosome 6 (Mb)
160.80
160.90
161.00
161.10
161.20
10
20
30
40
rs3798519
Position on Chromosome 6 (Mb)
50.6
50.7
50.8
50.9
51.0
5
10
15
rs11773845
Position on Chromosome 7 (Mb)
116.00
116.10
116.20
116.30
116.40
5
10
15
rs801011
Position on Chromosome 7 (Mb)
74.00
74.10
74.20
74.30
74.40
74.50
5
10
15
20

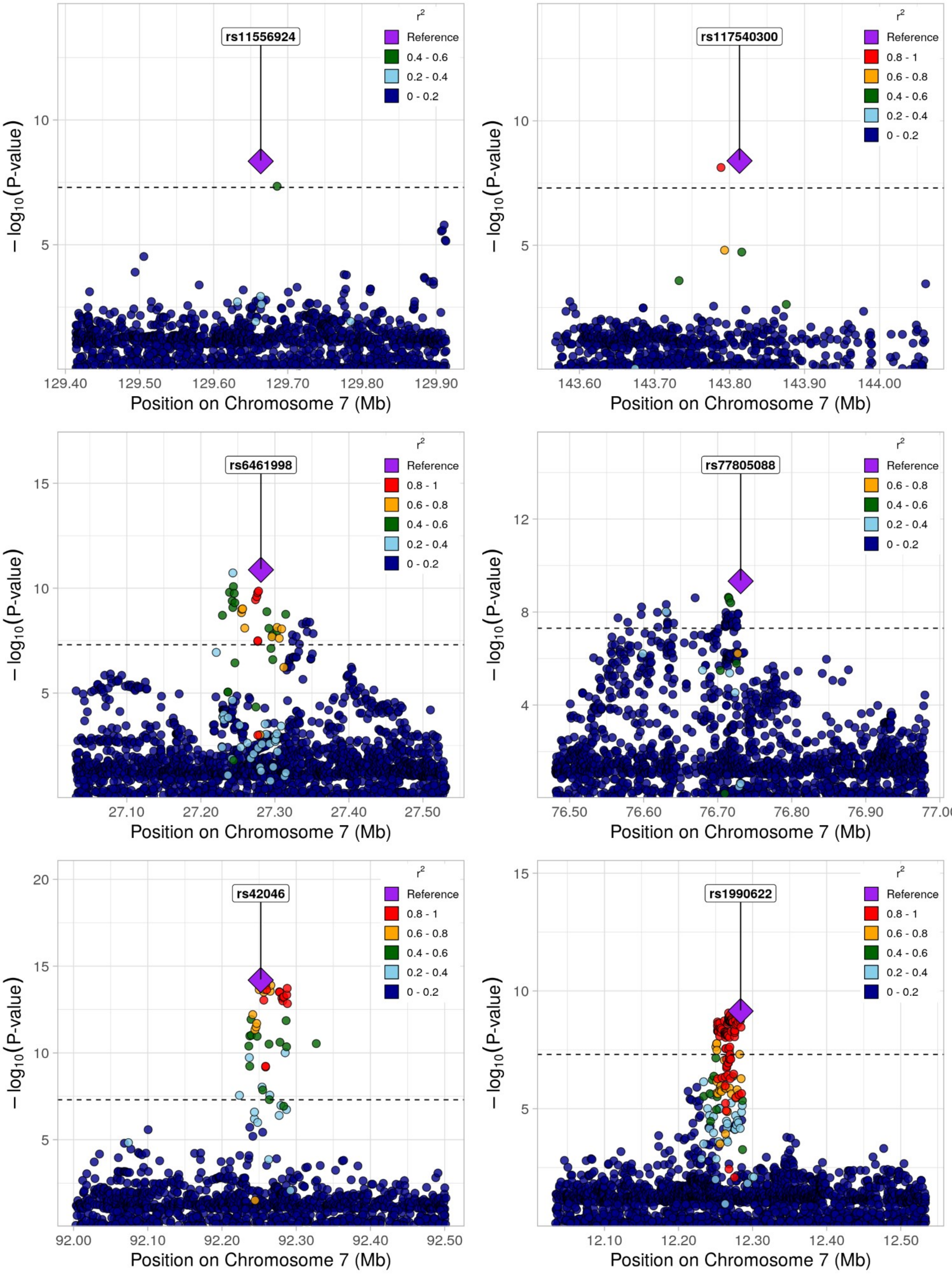
rs11556924
rs117540300
rs6461998
rs77805088
rs42046
rs1990622
r2
Reference
0.8 - 1
0.6 - 0.8
0.4 - 0.6
0.2 - 0.4
0 - 0.2
−log10(P-value)
Position on Chromosome 7 (Mb)

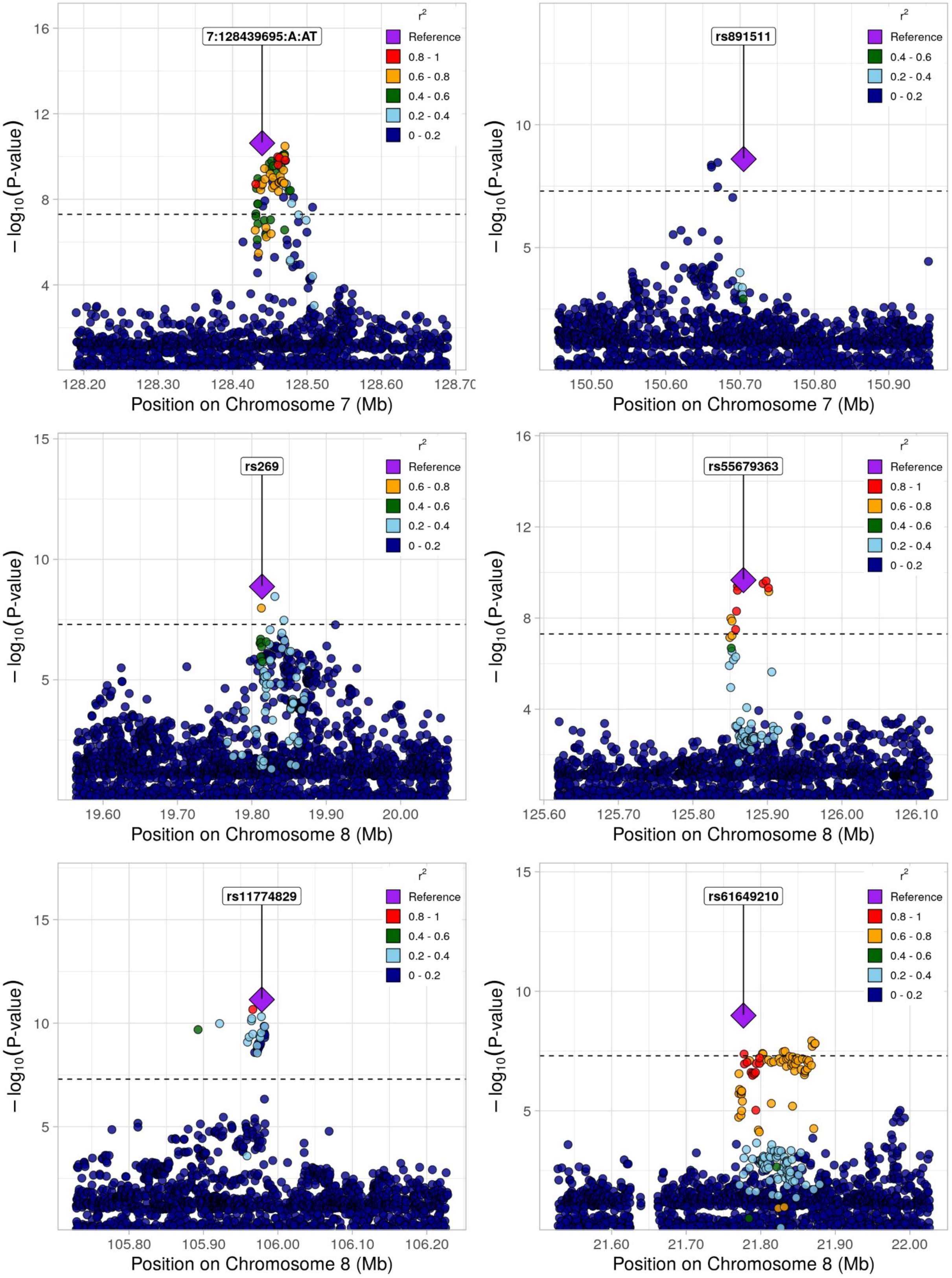
7:128439695:A:AT
rs891511
rs269
rs55679363
rs11774829
rs61649210
r²
Reference
0.8 - 1
0.6 - 0.8
0.4 - 0.6
0.2 - 0.4
0 - 0.2
− log10(P-value)
Position on Chromosome 7 (Mb)
Position on Chromosome 8 (Mb)
128.20
128.30
128.40
128.50
128.60
150.50
150.60
150.70
150.80
150.90
19.60
19.70
19.80
19.90
20.00
125.60
125.70
125.80
125.90
126.00
126.10
105.80
105.90
106.00
106.10
106.20
21.60
21.70
21.80
21.90
22.00

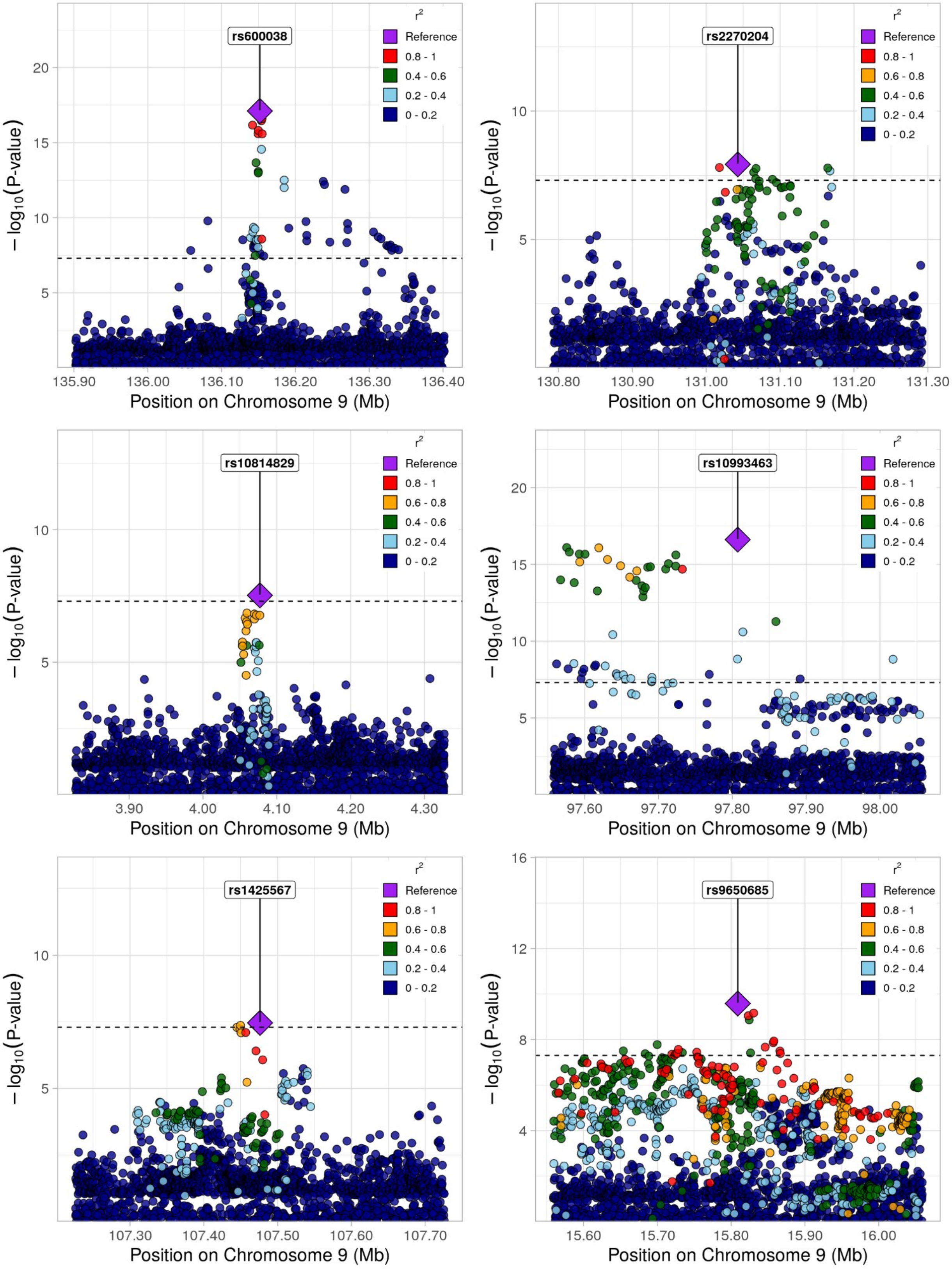
rs600038
r²
Reference
0.8 - 1
0.4 - 0.6
0.2 - 0.4
0 - 0.2
− log10(P-value)
Position on Chromosome 9 (Mb)
135.90
136.00
136.10
136.20
136.30
136.40
5
10
15
20
rs2270204
r²
Reference
0.8 - 1
0.6 - 0.8
0.4 - 0.6
0.2 - 0.4
0 - 0.2
− log10(P-value)
Position on Chromosome 9 (Mb)
130.80
130.90
131.00
131.10
131.20
131.30
5
10
rs10814829
r²
Reference
0.8 - 1
0.6 - 0.8
0.4 - 0.6
0.2 - 0.4
0 - 0.2
− log10(P-value)
Position on Chromosome 9 (Mb)
3.90
4.00
4.10
4.20
4.30
5
10
rs10993463
r²
Reference
0.8 - 1
0.6 - 0.8
0.4 - 0.6
0.2 - 0.4
0 - 0.2
− log10(P-value)
Position on Chromosome 9 (Mb)
97.60
97.70
97.80
97.90
98.00
5
10
15
20
rs1425567
r²
Reference
0.8 - 1
0.6 - 0.8
0.4 - 0.6
0.2 - 0.4
0 - 0.2
− log10(P-value)
Position on Chromosome 9 (Mb)
107.30
107.40
107.50
107.60
107.70
5
10
rs9650685
r²
Reference
0.8 - 1
0.6 - 0.8
0.4 - 0.6
0.2 - 0.4
0 - 0.2
− log10(P-value)
Position on Chromosome 9 (Mb)
15.60
15.70
15.80
15.90
16.00
4
8
12
16

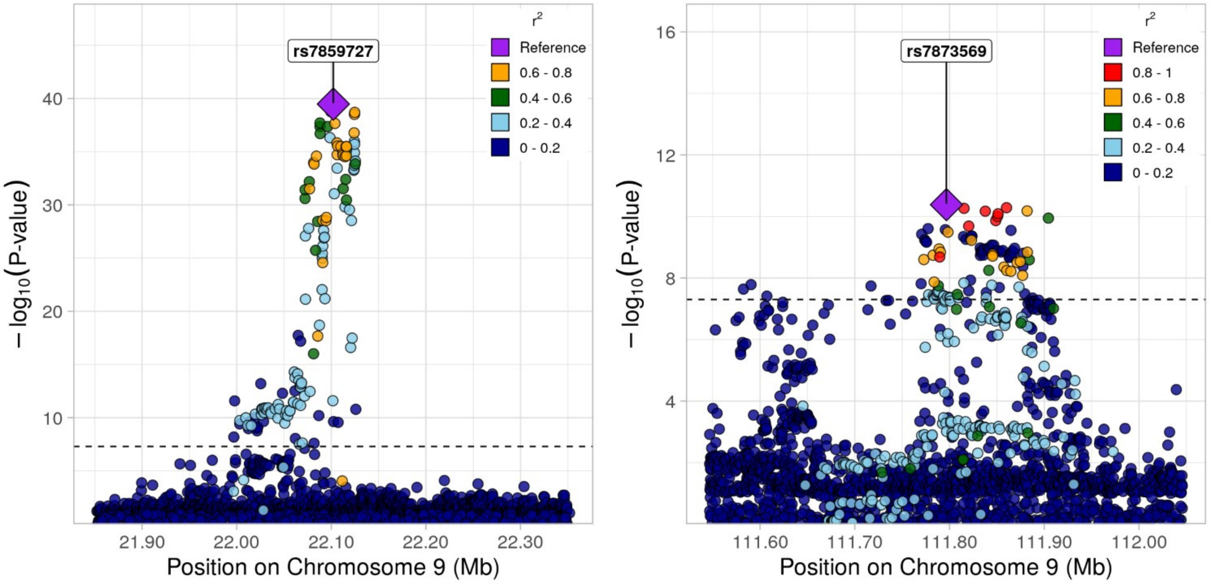
rs7859727
rs7873569
r²
Reference
0.8 - 1
0.6 - 0.8
0.4 - 0.6
0.2 - 0.4
0 - 0.2
− log10(P-value)
Position on Chromosome 9 (Mb)
21.90
22.00
22.10
22.20
22.30
111.60
111.70
111.80
111.90
112.00

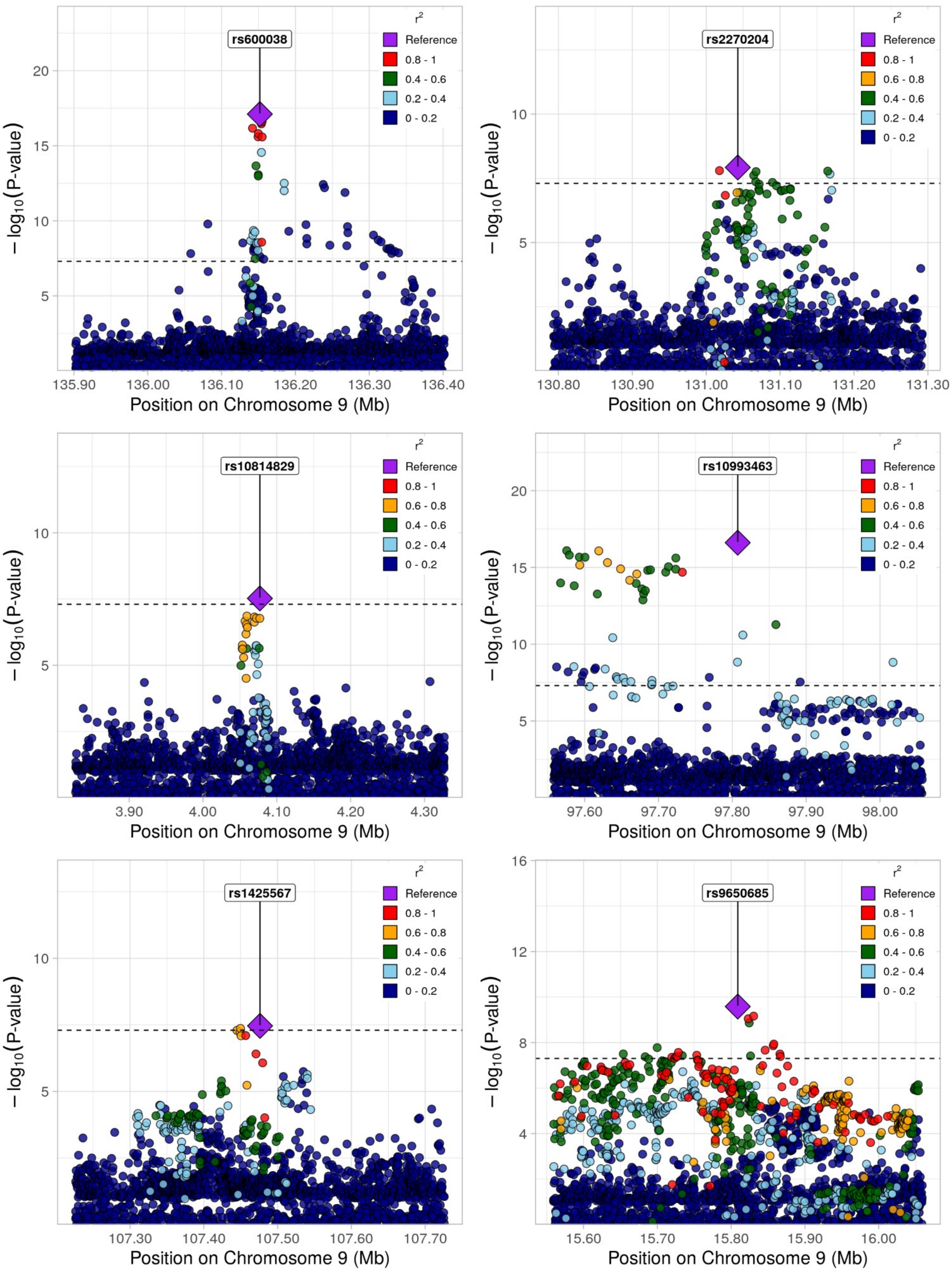
rs600038
rs2270204
rs10814829
rs10993463
rs1425567
rs9650685
r²
Reference
0.8 - 1
0.6 - 0.8
0.4 - 0.6
0.2 - 0.4
0 - 0.2
−log10(P-value)
Position on Chromosome 9 (Mb)
135.90
136.00
136.10
136.20
136.30
136.40
130.80
130.90
131.00
131.10
131.20
131.30
3.90
4.00
4.10
4.20
4.30
97.60
97.70
97.80
97.90
98.00
107.30
107.40
107.50
107.60
107.70
15.60
15.70
15.80
15.90
16.00

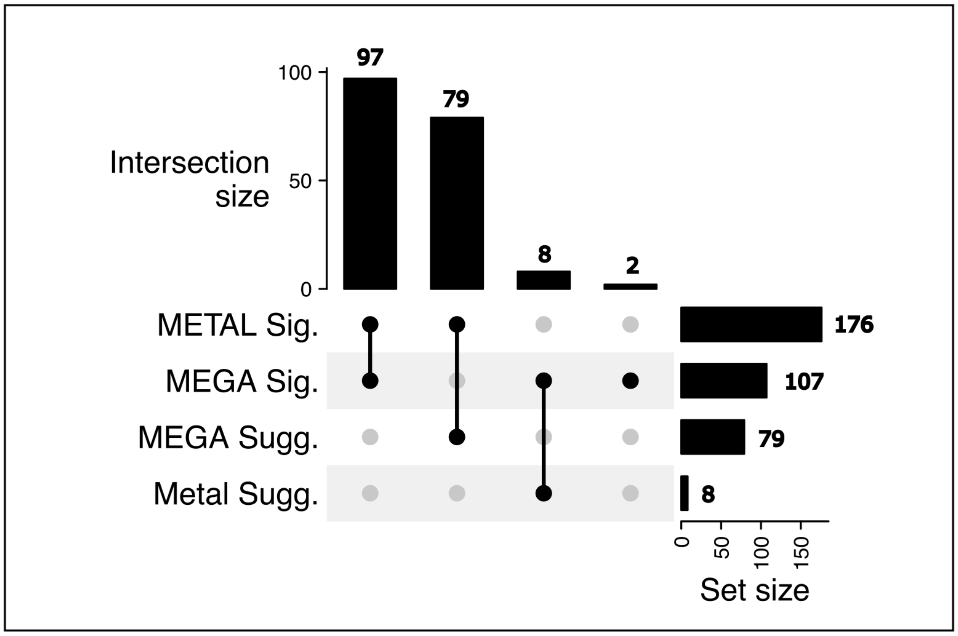


Suppl. Figure 2: Overlap between lead loci identified using METAL versus MR-MEGA. Loci passing genome-wide threshold (P-value < $5x10^{-8}$) are labeled as METAL Sig or MEGA Sig while loci only passing the Bonferroni-corrected threshold for replication (P-value < 0.05/176 for METAL loci, and P-value < 0.05/107 for MEGA loci) are labreled as METAL Sugg and MEGA Sugg. Two loci identified by MEGA were not identified by METAL as passing either the genome-wide significant or Bonferroni-corrected replication threshold.

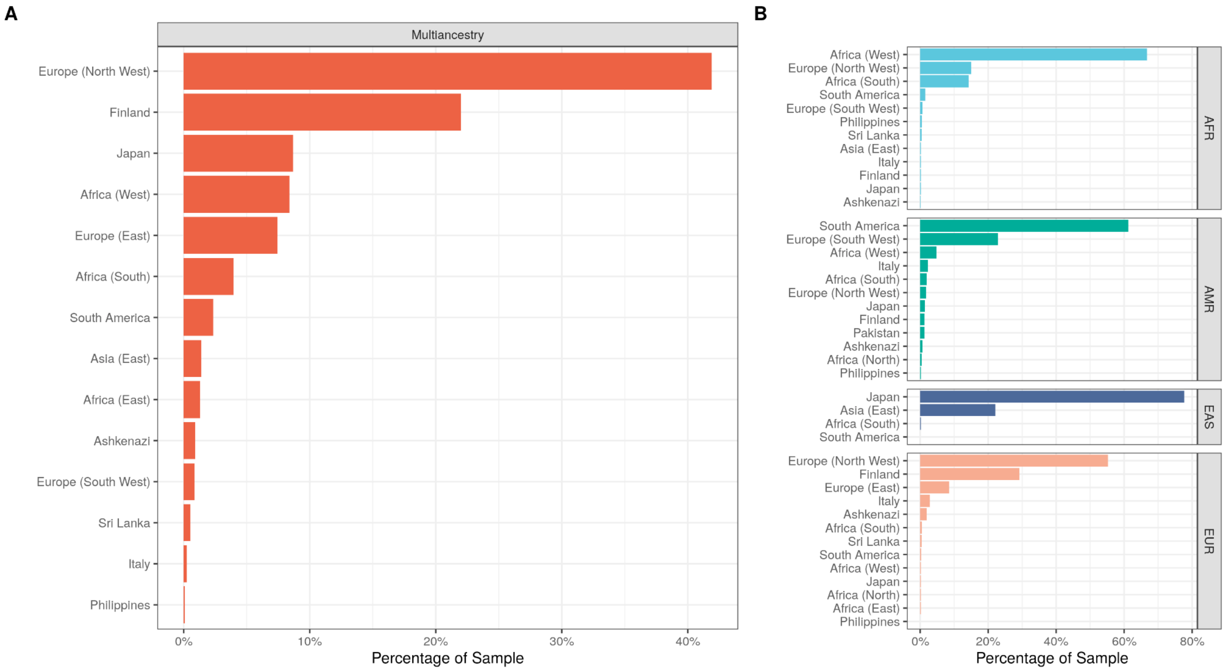


Suppl. Figure 3: Fine-scale inference of ancestry proportions using summary statistics, based on Prive et. al. 2022

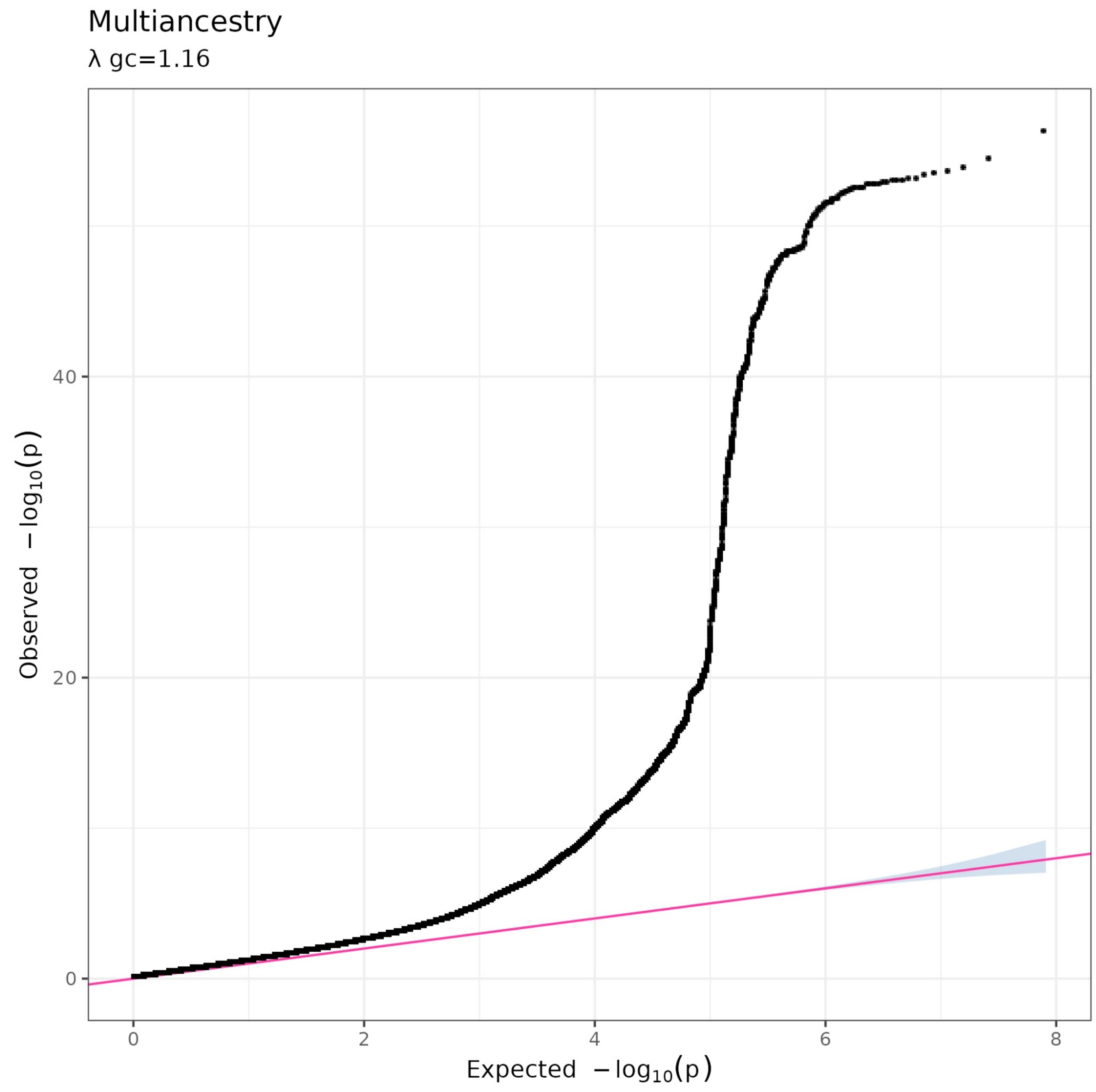


Suppl. Figure 4: Q-Q plot of p-value distribution from multi-ancestry GWAS meta analysis.

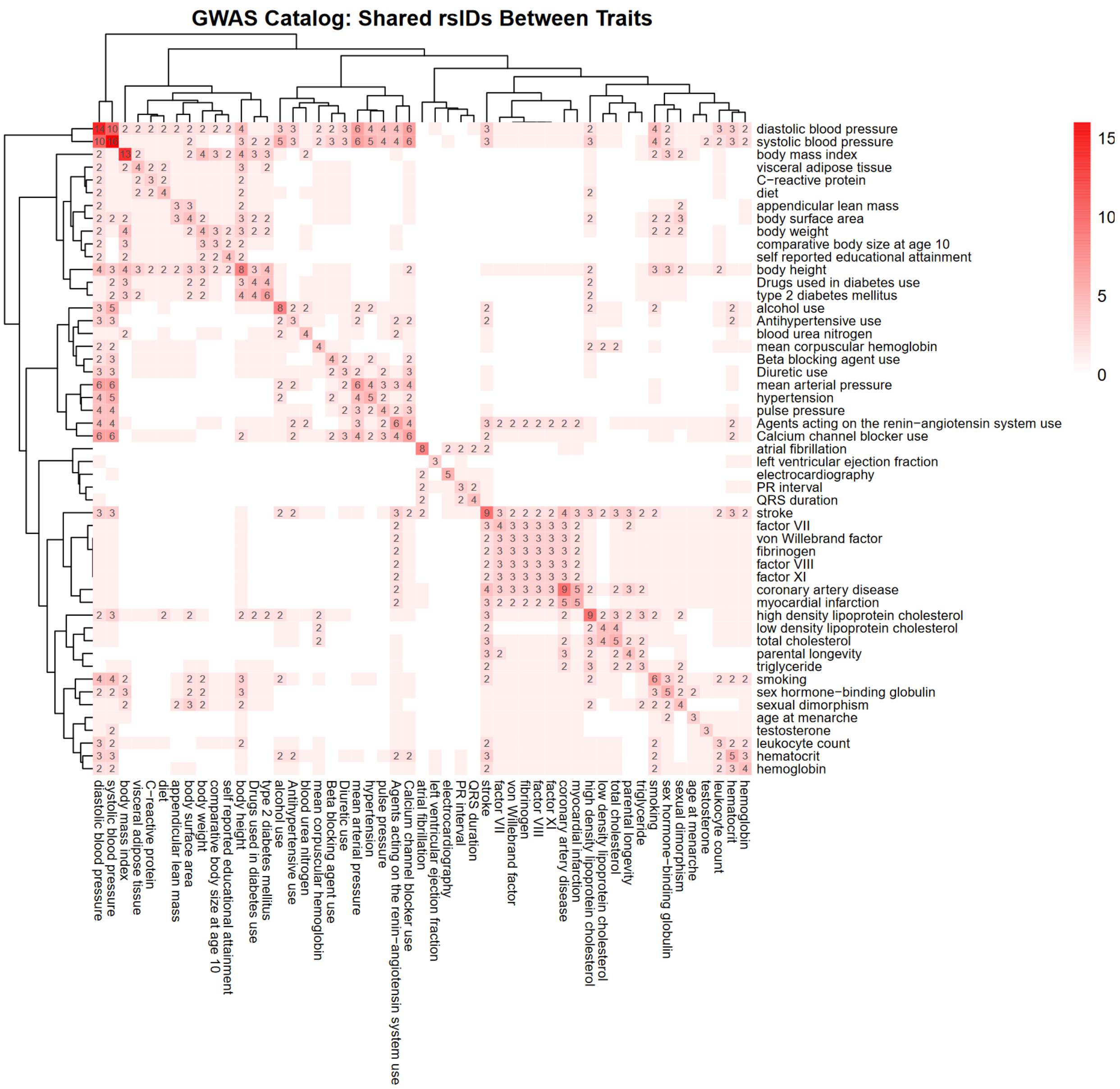


Suppl. Fig. 5: HF GWAS lead variants and shared trait associations in the NHGRI-EBI Catalog. Cells are labeled by their number of shared rsIDs with lead HF GWAS rsIDs. Cells on the diagonal display the number of shared rsIDs for each trait and HF. Hierarchical clustering was applied to rows and columns to identify groupings of shared traits.

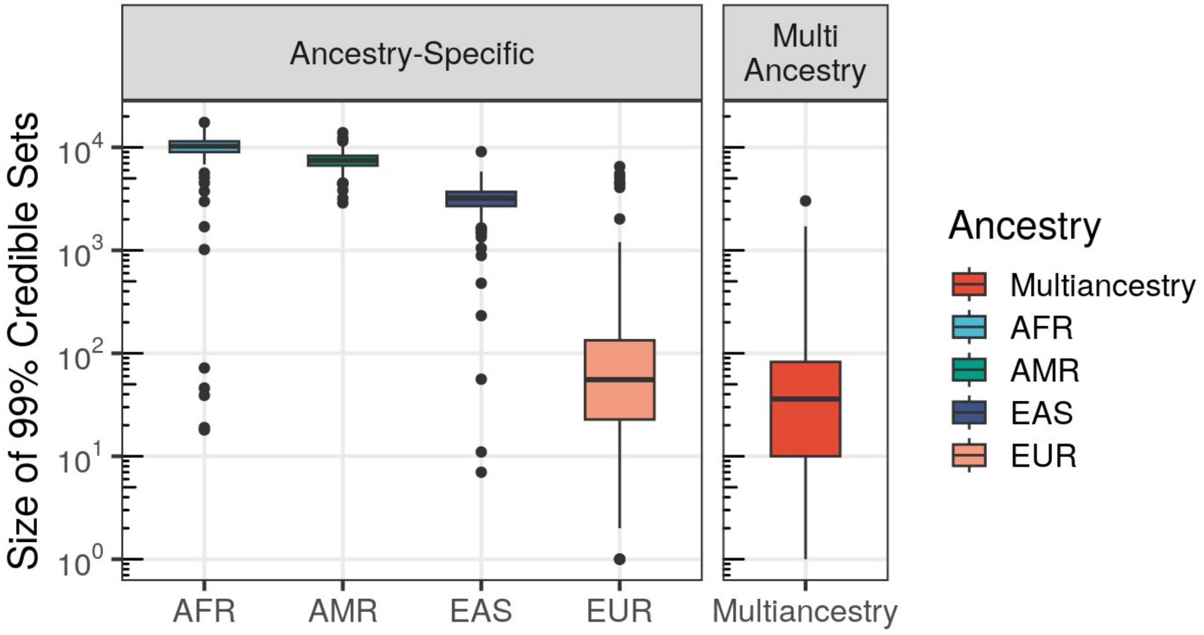


Suppl. Figure 6: Credible sets were generated surrounding the index variant at each 1MB locus identified in the multi-ancestry meta analysis, centered on the index variant (+/-500kb).

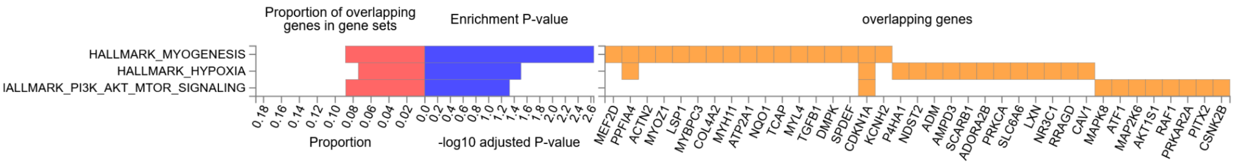


Suppl. Fig. 7: GWAS prioritized genes enrichment in hallmark pathways as determined by FUMA.

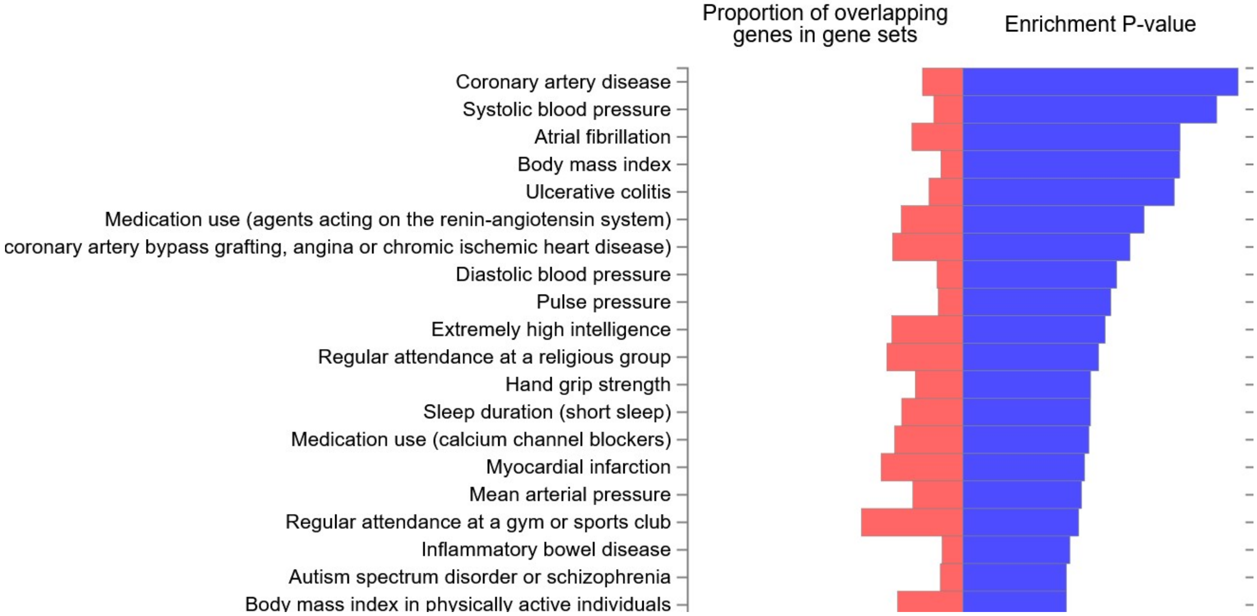


Suppl. Fig. 8: Top 20 GWAS catalog gene sets enriched for HF-prioritized genes as determined by FUMA.

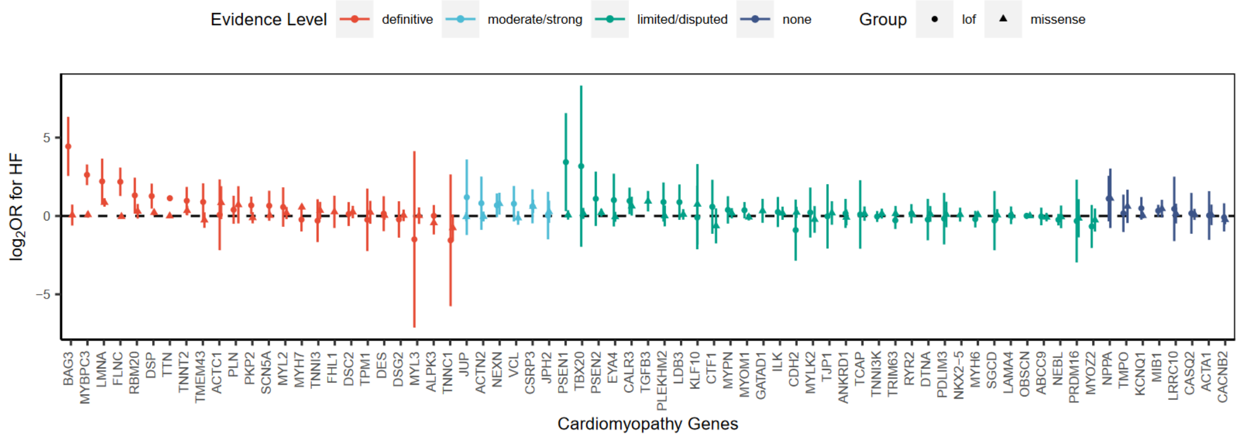


Suppl. Figure 9: Association of predicted damaging missense or high-confidence loss-of-function variants in known cardiomyopathy genes with all-cause HF in multi-population gene burden analysis. Variant carriers in the PMBB were classified based on presence or absence of a predicted damaging missense mutation (REVEL score >0.5) or a LOFTEE-classified high-confidence loss-of-function variant.

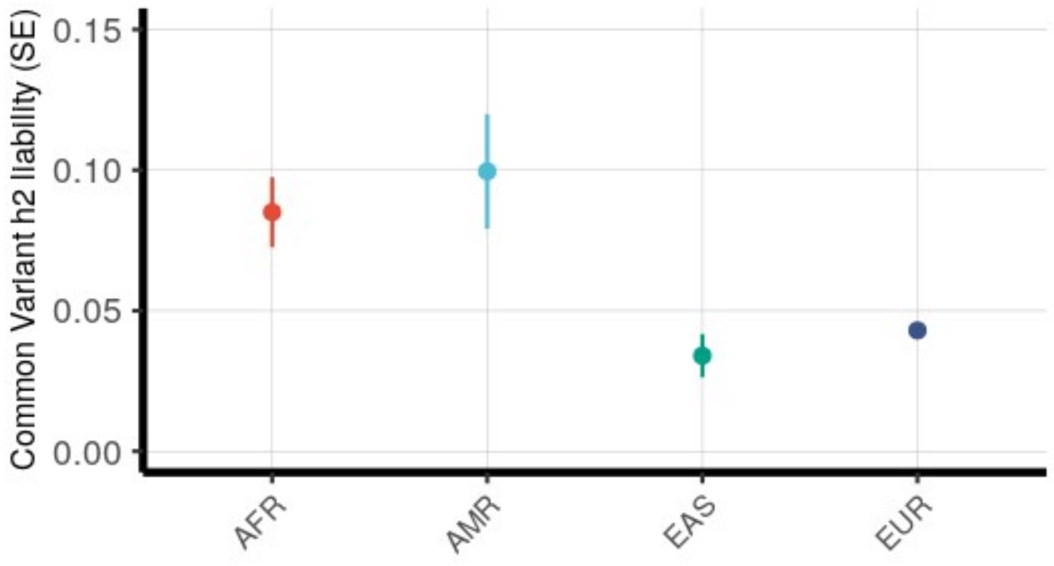

Suppl. Fig. 10: Common variant h2 estimates from HF GWAS across individuals of African, Admixed American, East Asian, and European ancestries. Heritability estimates were generated using LD score regression and LD reference panels published by the UKB.

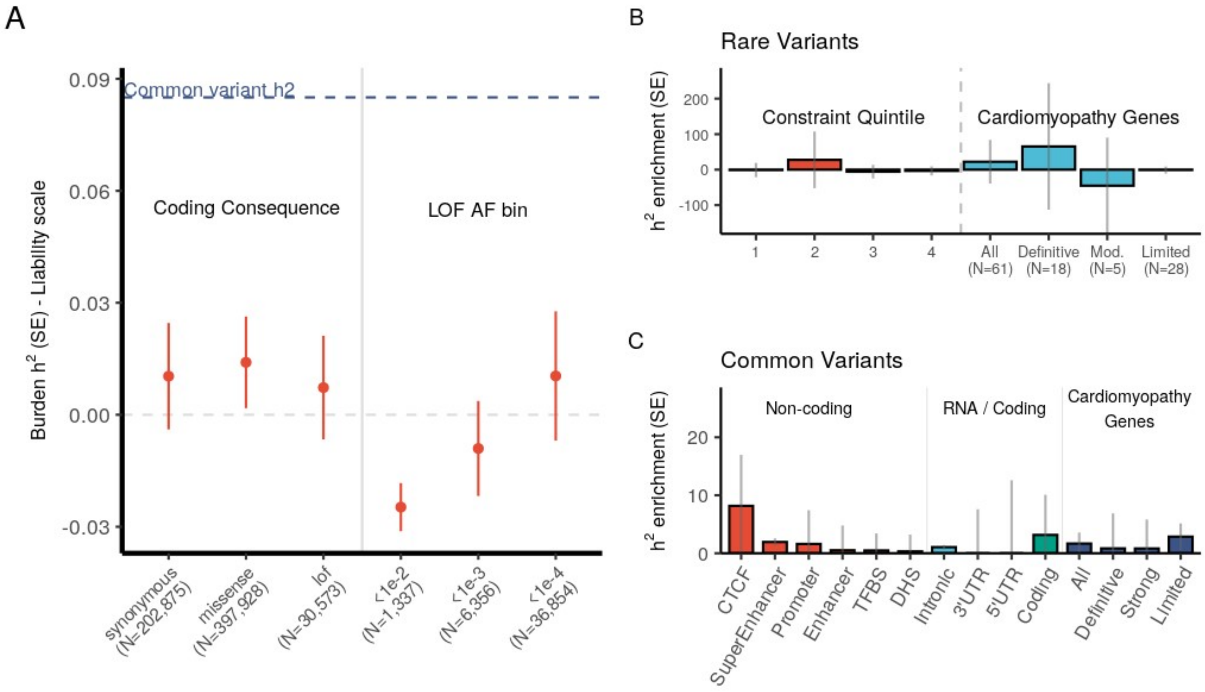


Suppl. Fig. 11: Burden h2 (liability scale) for PMBB AFR population across coding consequence and AF bin categories. Although not statistically significant, there is a trend for increased burden h2 in pLoF variants at more rare allele frequencies.

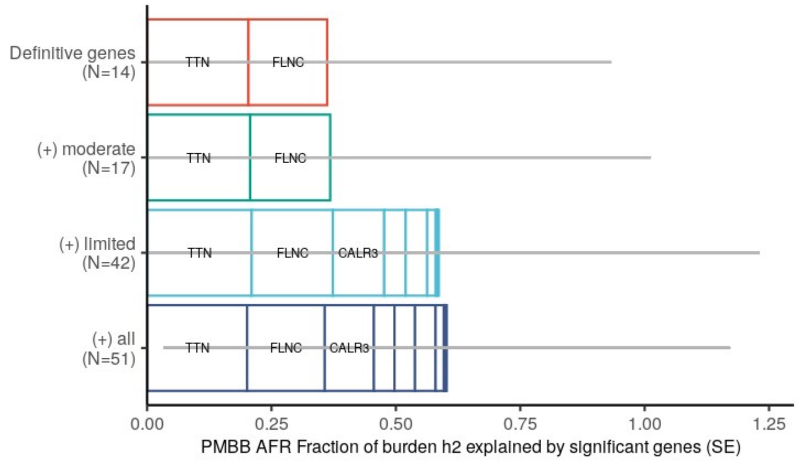


Suppl. Fig. 12: Fraction burden heritability explained by known cardiomyopathy genes in AFR HF participants.

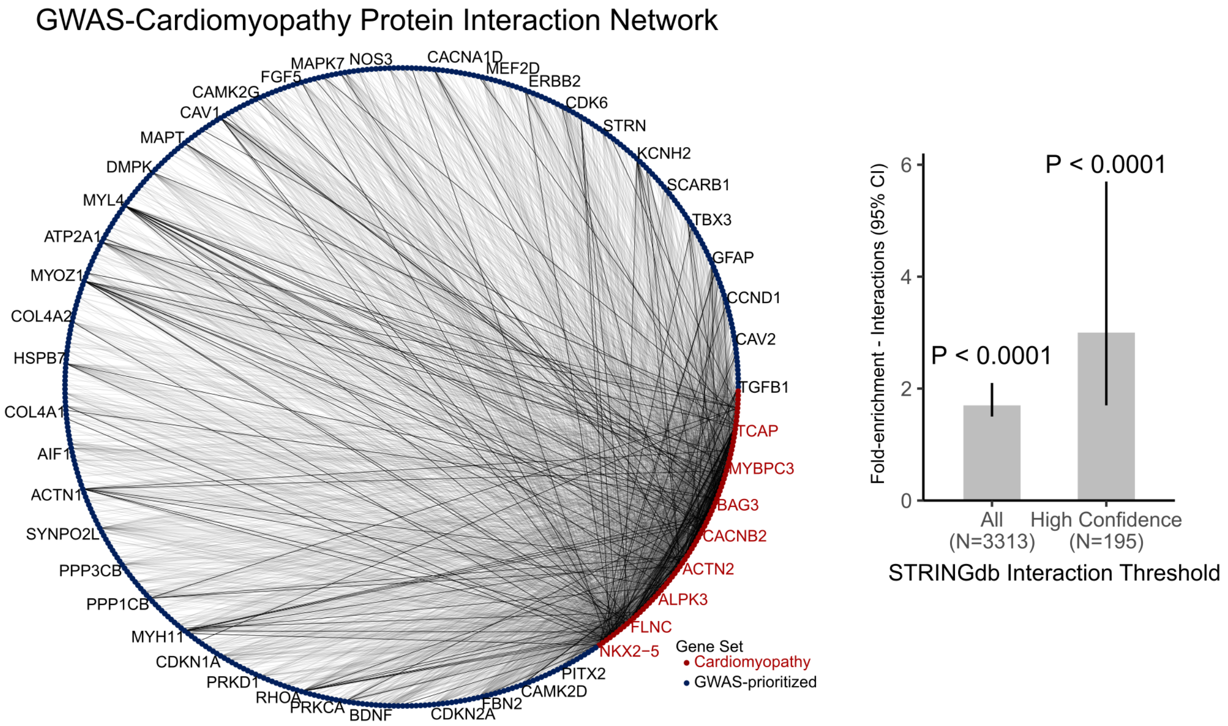


Suppl. Fig. 13: (A) StringDB interaction network between GWAS-nominated genes and known cardiomyopathy genes. Genes with >19 interactions are labeled. High-confidence interactions (combined score >/= 700) are depicted as black lines while non-high-confidence interactions are depicted as grey lines. (B) Enrichment for StringDB interactions between known cardiomyopathy genes and GWAS-nominated genes. P-values generated by comparing against a null distribution of interactions between randomly sampled sets of genes and known cardiomyopathy gene in StringDB.

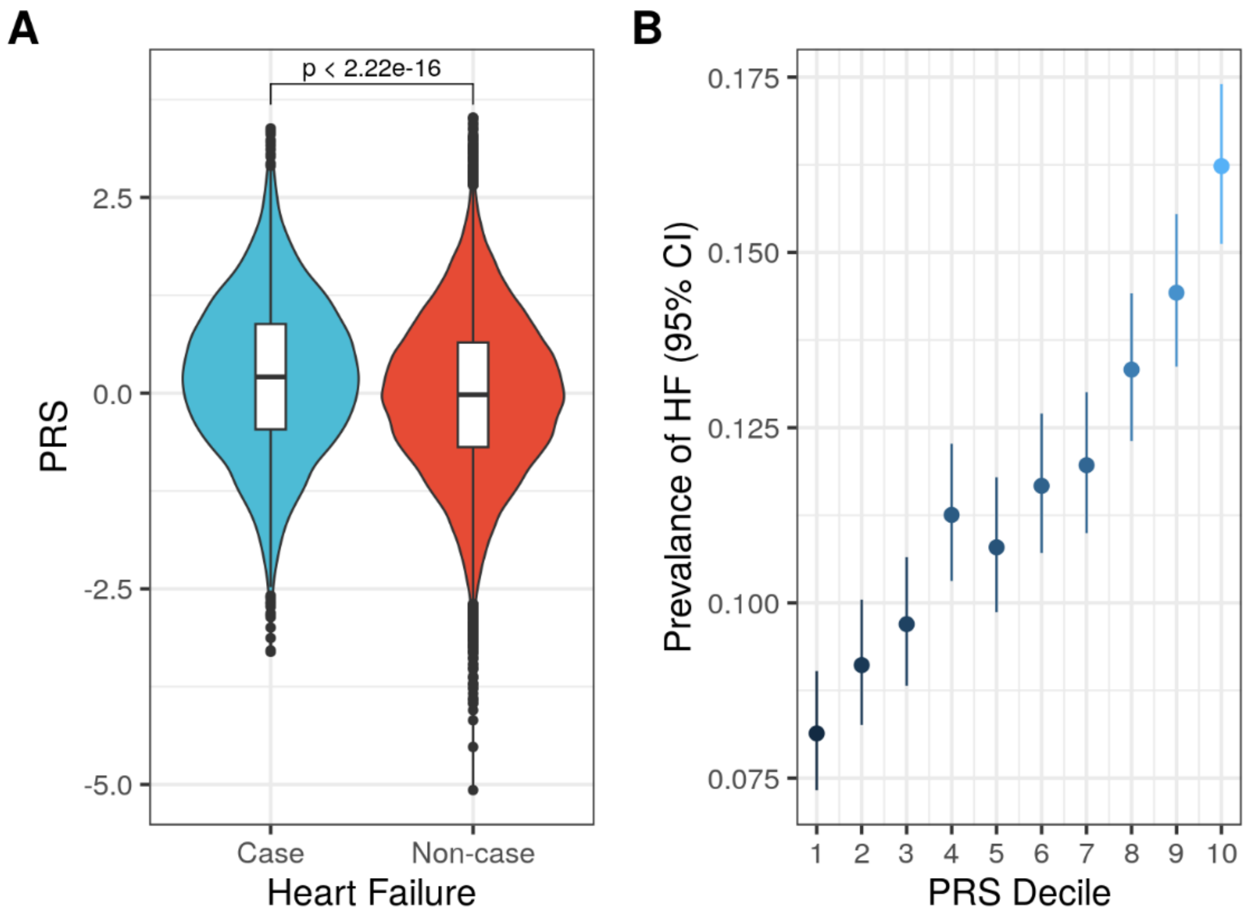


Suppl. Fig. 14: (A) HF PRS distributions among PMBB participants with and without HF. Difference in PRS was assessed using the Wilcox rank sum test. (B) Prevalence of HF among each PRS decile. 95% CI = 95% Confidence Interval.

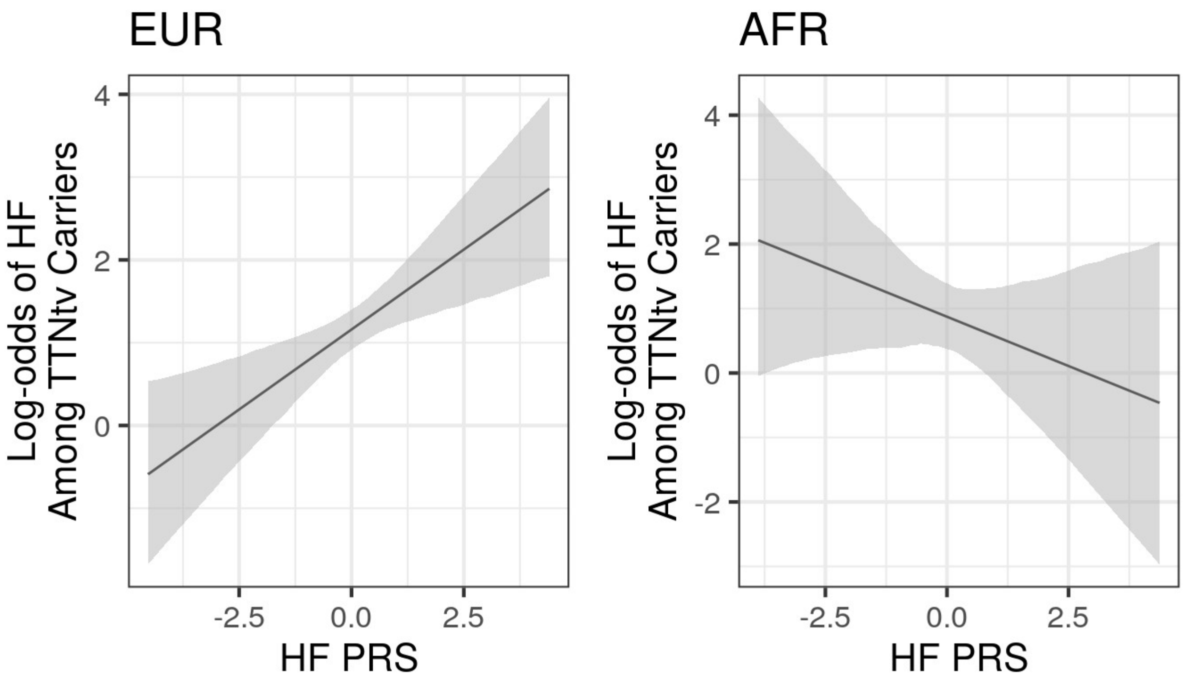


Suppl. Fig. 15: Log-odds of HF in the PMBB for TTNtv carriers as a function of multiancestry PRS Z-score scaled separately by ancestry for EUR and AFR populations. There is a significant interaction between HF PRS and TTNtv carrier status in the PMBB EUR population (OR 4.35, 95% CI: 3.42-5.52) but not for the PMBB AFR population (OR 2.10, 95% CI: 1.22-3.61).
